# Proteomic Signature of HIV-Associated Subclinical Left Atrial Remodeling and Incident Heart Failure

**DOI:** 10.1101/2024.02.13.24302797

**Authors:** Tess E Peterson, Virginia S Hahn, Ruin Moaddel, Min Zhu, Sabina A Haberlen, Frank J Palella, Michael Plankey, Joel S Bader, Joao AC Lima, Robert E Gerszten, Jerome I Rotter, Stephen S Rich, Susan R Heckbert, Gregory D Kirk, Damani A Piggott, Luigi Ferrucci, Joseph B Margolick, Todd T Brown, Katherine C Wu, Wendy S Post

## Abstract

**Background:** People living with HIV (PLWH) are at higher risk of heart failure (HF) and preceding subclinical cardiac abnormalities, including left atrial dilation, compared to people without HIV (PWOH). Hypothesized mechanisms include premature aging linked to chronic immune activation. We leveraged plasma proteomics to identify potential novel contributors to HIV-associated differences in indexed left atrial volume (LAVi) among PLWH and PWOH and externally validated identified proteomic signatures with incident HF among a cohort of older PWOH.

**Methods:** We performed proteomics (Olink Explore 3072) on plasma obtained concurrently with cardiac magnetic resonance imaging among PLWH and PWOH in the United States. Proteins were analyzed individually and as agnostically defined clusters. Cross-sectional associations with HIV and LAVi were estimated using multivariable regression with robust variance. Among an independent general population cohort, we estimated associations between identified signatures and LAVi using linear regression and incident HF using Cox regression.

**Results:** Among 352 participants (age 55±6 years; 25% female), 61% were PLWH (88% on ART; 73% with undetectable HIV RNA) and mean LAVi was 29±9 mL/m^2^. Of 2594 analyzed proteins, 439 were associated with HIV serostatus, independent of demographics, hepatitis C virus infection, renal function, and substance use (FDR<0.05). We identified 73 of these proteins as candidate contributors to the independent association between positive HIV serostatus and higher LAVi, enriched in tumor necrosis factor (TNF) signaling and immune checkpoint proteins regulating T cell, B cell, and NK cell activation. We identified one protein cluster associated with LAVi and HIV regardless of HIV viral suppression status, which comprised 42 proteins enriched in TNF signaling, ephrin signaling, and extracellular matrix (ECM) organization. This protein cluster and 30 of 73 individual proteins were associated with incident HF among 2273 older PWOH (age 68±9 years; 52% female; 8.5±1.4 years of follow-up).

**Conclusion:** Proteomic signatures that may contribute to HIV-associated LA remodeling were enriched in immune checkpoint proteins, cytokine signaling, and ECM organization. These signatures were also associated with incident HF among older PWOH, suggesting specific markers of chronic immune activation, systemic inflammation, and fibrosis may identify shared pathways in HIV and aging that contribute to risk of HF.

## INTRODUCTION

With improved access to early and effective antiretroviral therapy (**ART**), HIV has become a chronic condition marked by higher risk and earlier onset of aging-related diseases, including cardiovascular disease (**CVD**).^1^ People living with HIV (**PLWH**), even when virologically suppressed due to ART, are at higher risk of myocardial diseases,^2^ such as incident heart failure (**HF**)^3–5^ and atrial fibrillation (**AF**).^6^ Advanced HIV disease has been classically associated with left ventricular (**LV**) systolic dysfunction and dilated cardiomyopathy, but with effective ART, this phenotype has transitioned to one characterized by subclinical abnormalities in cardiac structure and function that may presage clinical HF, particularly HF with preserved ejection fraction (**EF**).^7^ We previously demonstrated PLWH have higher risk of diastolic dysfunction and related structural abnormalities compared to people without HIV (**PWOH**), including subclinical left atrial (**LA**) enlargement,^8,9^ which has been supported by other data across world regions.^10–13^ Indexed left atrial volume (**LAVi**) reflects the cumulative effect of chronically elevated LV filling pressure and is an independent predictor of diastolic dysfunction and clinical HF.^14^

Mechanisms contributing to HIV-associated myocardial disease are hypothesized to be multifactorial, mediated in large part by chronic immune activation and dysfunction that persists despite ART-induced viral suppression. Hypothesized contributors include direct immune activation and exhaustion due to HIV viral persistence and co-pathogens, metabolic dysfunction due to both HIV and use of certain antiretrovirals, higher mucosal permeability and microbial translocation, and higher prevalence of risk factors with known inflammatory effects, such as substance use.^7^ However, it remains unclear to what degree mechanisms underlying this risk differ among PLWH compared to PWOH, among whom aging-related immunosenescence and inflammaging may similarly contribute to clinical HF risk. Moreover, therapeutic and preventive strategies among PLWH, particularly for HF, are largely guided by data from clinical trials conducted among PWOH.

Proteomics provides a high-throughput discovery approach to generate novel mechanistic hypotheses that could lead to improved risk prediction, disease characterization, and novel targeted pharmacologic therapies. We performed proteomic analyses using the Olink Explore 3072 platform on stored plasma obtained concurrently with cardiovascular magnetic resonance (**CMR**) among largely ART-treated PLWH and sociodemographically similar PWOH enrolled in the Subclinical Myocardial Abnormalities in HIV (**SMASH**) study,^9^ the discovery cohort. Our objective was to identify plasma proteomic signatures that may represent novel contributors to HIV-associated atrial remodeling. We then sought to evaluate if identified proteomic signatures were associated with LAVi, incident AF, and incident adjudicated clinical HF among a large independent external validation cohort of older PWOH in the Multi-Ethnic Study of Atherosclerosis (**MESA**). Such a validation approach could provide support for a premature aging hypothesis among PLWH and identify common and potentially novel mechanistic pathways that contribute to higher risk of clinical HF among older PWOH. A full study overview is depicted in **FIGURE 1**.

**FIGURE 1.**
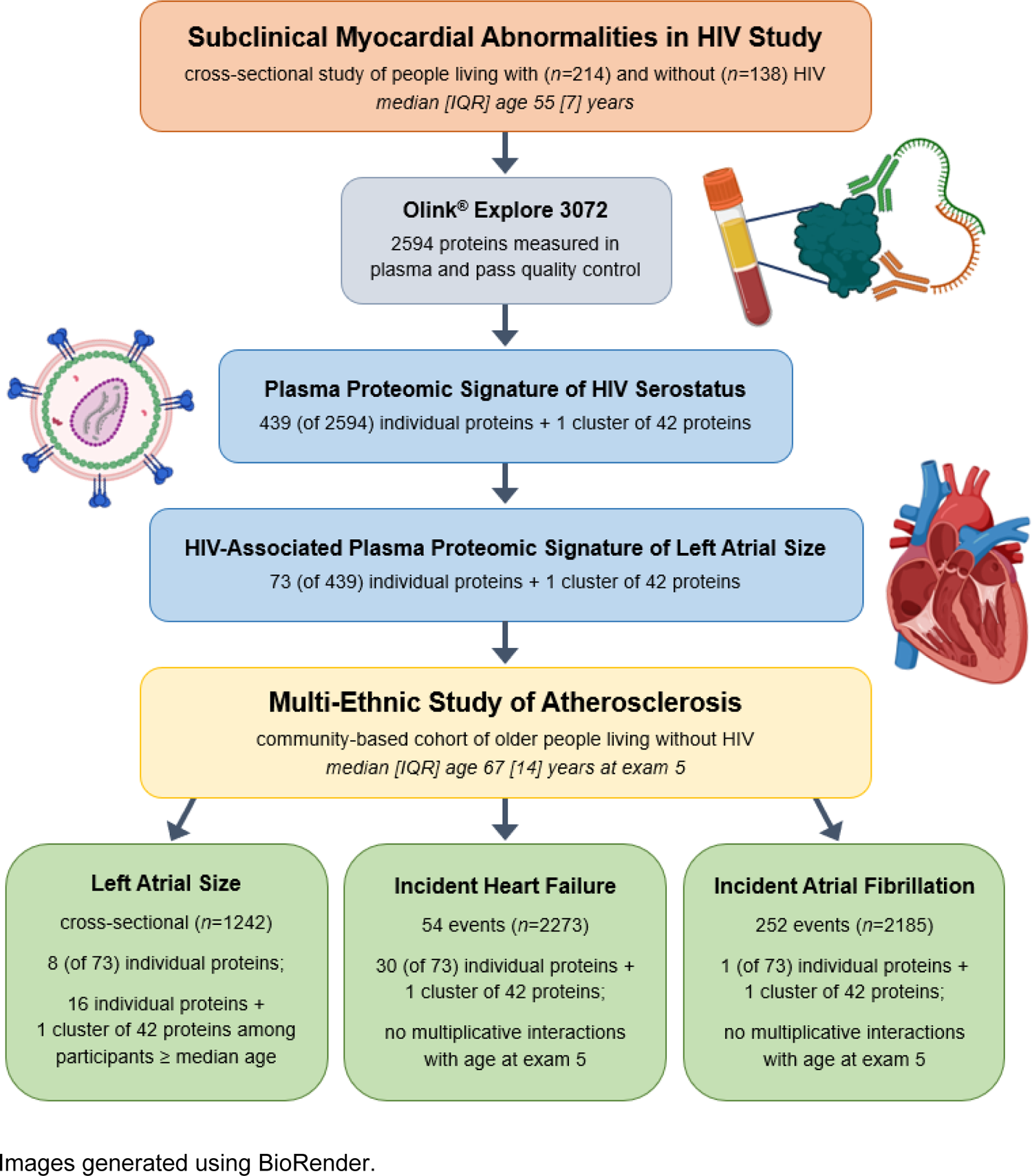
Study overview

## METHODS

### Discovery Study Population (SMASH)

Participants in the SMASH study were recruited from the Multicenter AIDS Cohort Study (**MACS**), the Women’s Interagency HIV Study (**WIHS**), and the AIDS Linked to the IntraVenous Experience (**ALIVE**) study and underwent CMR with concurrent blood collection and storage between March 2015 and February 2018. All three cohorts include PLWH and PWOH. The WIHS and MACS are multicenter observational longitudinal cohort studies composed of women or men who have sex with men, respectively.^15,16^ The ALIVE study is a community-based cohort of men and women with a history of injection drug use.^17^ See the ***Supplemental Methods*** for additional cohort details. The SMASH study enrolled active participants 40-70 years of age from the Chicago and Baltimore/Washington, DC MACS sites; the Washington, DC WIHS site; and the ALIVE study, which is based in Baltimore. Exclusion criteria included estimated glomerular filtration rate (**eGFR**, Chronic Kidney Disease Epidemiology Collaboration 2021) <45 mL/min/1.73 m^2^, weight >350 pounds, known claustrophobia or contrast allergy, and contraindications to CMR. Proteomics was performed on a subset of total SMASH participants with complete CMR and adequate stored plasma volume (***Supplemental Figure S1A***). The Institutional Review Boards of Johns Hopkins University, Georgetown University, and Northwestern University approved this study, and all participants signed informed consent.

Given the sensitive nature of data collected for this study, requests for data sharing from researchers certified in human subject confidentiality can be submitted to and will be reviewed by the parent cohorts: https://statepi.jhsph.edu/mwccs/ and https://www.jhsph.edu/research/affiliated-programs/aids-linked-to-the-intravenous-experience/.

### Sociobehavioral and Clinical Data in SMASH

Participants completed an interviewer-administered structured questionnaire and biological measures concurrent with CMR. Prescribed medications, history of CVD, and substance use during the preceding 5 years were queried. These data were supplemented with data collected through the parent cohorts, including demographics and CVD risk factors. See ***Supplemental Methods*** for a full list and definitions of covariates. Among PLWH, measures of HIV disease activity included current plasma HIV RNA concentrations (Roche ultrasensitive assay), current and nadir CD4+ T cell counts/µL, history of AIDS-defining malignancy or opportunistic infection, and use of ART, including regimen.

### Cardiovascular Magnetic Resonance in SMASH

CMR was performed at Johns Hopkins Hospital (Baltimore) or Northwestern Memorial Hospital (Chicago) on 1.5 T Siemens Avanto or Aera scanners (Erlangen, Germany) using a standardized protocol that included gadolinium enhancement, detailed previously.^9^ Briefly, short and long-axis cines (30 phases/cardiac cycle) were acquired with a steady-state free precession sequence. Multimodality Tissue Tracking software (MTT; version 6.0, Toshiba, Japan) was used to quantify LA volumes and other standard metrics,^18^ blinded to participant characteristics. Maximum LA volume was measured using the LA volume curve generated by the Simpson’s method from four-chamber and two-chamber views and was indexed to body surface area (LAVi).

### Proteomics in SMASH

We performed proteomics on EDTA plasma stored at −80°C using the Olink Explore 3072 platform at the Laboratory of Clinical Investigation at the National Institute on Aging, NIH (Baltimore, MD) using a standardized protocol. Olink technology has been described previously^19^ and is outlined in the ***Supplemental Methods*** alongside details on data quality control, normalization, and calibration. Values are reported as log_2_-transformed relative plasma abundances, which were winsorized to 5 standard deviations (**SD**) to minimize the effect of outliers and standardized for comparability. Following quality control, no samples were excluded, and 2594 proteins were analyzed with final mean intra- and inter-assay coefficients of variation of 9% and 15%, respectively.

### Weighted Gene Co-expression Network Analysis

High-dimensional proteomic data often contain informative patterns lost when analyzing proteins individually. To address this, we derived unique clusters of highly correlated proteins using weighted gene co-expression network analysis (**WGCNA**) implementing the WGCNA package in R (version 4.2). See ***Supplemental Methods*** for details.

### Statistical Modeling

Complete case analyses were performed to estimate the cross-sectional associations between plasma protein abundances and both HIV serostatus and continuous LAVi. Proteins were analyzed individually (single-analyte approach) and as clusters agnostically defined using WGCNA (multi-analyte approach). Inference was made using multivariable linear regression with Huber-White robust variance estimators to account for likely heteroscedasticity. Analyses were corrected for multiple comparisons using a step-down false discovery rate (**FDR**) procedure defined by Benjamini and Hochberg and a threshold for significance of FDR<0.05.

Mean standardized differences in plasma protein abundances by HIV serostatus were first estimated adjusting for age, sex, race/ethnicity, educational attainment (dichotomized at high school diploma), current hazardous alcohol use, pack-years of smoking in prior 5 years, stimulant use in prior 5 years, opioid use in prior 5 years, hepatitis C virus infection (**HCV**), and eGFR. Mean differences in LAVi were then estimated per SD increment in individual plasma protein abundances, restricting to those proteins with HIV associations (at FDR<0.05) and adjusting for covariates above, as well as HIV serostatus, body mass index (**BMI**), systolic blood pressure (**SBP**), hypertension medication use, dyslipidemia, and diabetes. Proteins of interest were defined as those with significant independent associations with both HIV serostatus and LAVi. We further analyzed these proteins in subgroup analyses excluding PLWH with detectable plasma HIV RNA (>50 copies/mL). We assessed the degree to which they may reflect the independent association between positive HIV serostatus and higher LAVi. We evaluated differences in their associations with LAVi by HIV serostatus and age using multiplicative interaction terms. We estimated protein associations with basic clinical and HIV-related characteristics. All analyses were conducted using R (version 4.2).

### Functional Annotation Analysis

Annotation-based overrepresentation analyses were performed using the Gene Ontology (GO): Biological Processes annotation database and the clusterProfiler package in R (version 4.2).^20^ This employs a Fisher’s exact test comparing the proportion of total significantly associated proteins mapping to a given functional annotation to the proportion of total analyzed proteins mapping to the same annotation. The background used in these analyses was proteins measured on Olink Explore 3072 that passed quality control. Relationships between proteins of interest were further explored using interaction networks generated using a database of known and predicted protein-protein interactions, STRING.^21^

### External Validation Cohort (Older Persons without HIV in MESA)

External validation was performed in MESA, a prospective community-based cohort initiated in 2000 to study the characteristics and progression of subclinical CVD among a diverse population in the United States, age 45-84 years without clinical CVD at baseline.^22^ We used data from exam 5 (years 2010-2012) when participants were age 53-94 years due to use of the same CMR protocol^23^ and proteomics platform (Olink Explore 3072) as SMASH at that time point. MESA participants were also, on average, over 10 years older than SMASH participants at exam 5, allowing more extensive evaluation of the impact of older age on the identified proteomic signature among PWOH. See ***Supplemental Methods*** for study population and data acquisition details.

We first performed cross-sectional analyses of LAVi among MESA participants with proteomics and CMR using the same approach to statistical modeling as described above, with slight variation in covariates due to differences in data collection or prevalences among study populations—specifically, we adjusted for field center, age, sex, race/ethnicity, education level, smoking, BMI, SBP, blood pressure-lowering therapy, dyslipidemia, diabetes, and eGFR.

We additionally assessed associations of identified proteomic signatures with time to incident AF and incident clinical HF. AF was defined by the presence of ≥1 inpatient or outpatient International Classification of Diseases, Ninth Revision codes for AF. HF criteria included symptomatic, physician-diagnosed HF as well as ≥1 of the following: (i) pulmonary edema/congestion by chest x-ray; (ii) dilated LV or poor LV function by echocardiography or ventriculography; and/or (iii) evidence of LV diastolic dysfunction. Inpatient and/or outpatient records were independently reviewed by two physicians for HF classification and event date assignment. See ***Supplemental Methods*** for detailed ascertainment and adjudication procedures. Hazard ratios (**HR**) were estimated per SD increment in plasma protein level using Cox proportional hazards models adjusting for the same covariates as cross-sectional analyses. Participants with prevalent disease (AF or HF) at the start of follow-up were excluded from those analyses, and time at risk was calculated as the time from the date of MESA exam 5 to an incident event, death, loss to follow-up, or the end of follow-up, whichever occurred first. Lastly, we estimated associations between proteins validated with at least one outcome and basic clinical characteristics independent of field center.

## RESULTS

### SMASH Participant Characteristics

Of 468 persons initially enrolled in SMASH, 352 were selected for plasma proteomics based on availability of complete CMR data, including gadolinium enhancement, and adequate stored plasma volume (***Figure S1A***). Selected participants had lower representation of females and lower prevalence of hypertension and HCV compared to unselected participants (***Table S1***).

Demographic, clinical, and CMR characteristics of selected participants are summarized in **TABLE 1** by HIV serostatus and by parent cohort in ***Table S2***. Participants were mean (SD) age 55 (6) years, 25% female, 61% PLWH, and had predominantly normal LVEF. LAVi was higher among PLWH compared to PWOH—29.7 (11.3) and 27.7 (8.3) mL/m^2^, respectively (*p*=0.02 for difference, adjusting for demographics, substance use, and traditional CVD risk factors).

**TABLE 1.** SMASH participant characteristics by HIV serostatus (*n*=352)

| CHARACTERISTIC | Median [IQR] or % (n) |  |
| --- | --- | --- |
|  | PLWH (n=214) | PWOH (n=138) |
| Study site | --- | --- |
| MACS Baltimore | 75 (35%) | 46 (33%) |
| MACS Chicago | 28 (13%) | 17 (12%) |
| WIHS | 39 (18%) | 21 (15%) |
| ALIVE | 72 (34%) | 54 (39%) |
| *Demographics* |  |  |
| Age, years | 55 [51, 58] | 55 [52, 58] |
| Female sex at birth | 55 (26%) | 34 (25%) |
| Race and ethnicity | --- | --- |
| Black, non-Hispanic | 151 (71%) | 94 (68%) |
| White, non-Hispanic | 52 (24%) | 34 (25%) |
| Hispanic | 11 (5%) | 10 (7%) |
| Education level ≥ high school | 156 (73%) | 108 (78%) |
| *Substance Use* |  |  |
| Smoking status | --- | --- |
| Current | 114 (53%) | 57 (41%) |
| Former | 56 (26%) | 57 (41%) |
| Never | 44 (21%) | 24 (17%) |
| Pack-years of smoking <sup>a</sup> | 0.99 [0.00, 2.82] | 0.21 [0.00, 2.16] |
| Hazardous alcohol use (AUDIT score >8) | 23 (11%) | 20 (14%) |
| Opioid use <sup>a</sup> | 61 (29%) | 47 (34%) |
| Stimulant use <sup>a</sup> | 87 (41%) | 50 (36%) |
| *Clinical Factors* |  |  |
| History of cardiovascular disease | 14 (7%) | 8 (6%) |
| Body mass index, kg/m<sup>2</sup> | 25.7 [23.0, 29.3] | 26.9 [24.1, 31.0] |
| Hypertension <sup>b</sup> | 108 (51%) | 74 (54%) |
| Systolic blood pressure, mmHg | 124 [118, 134] | 128 [121, 135] |
| Blood pressure-lowering medication use | 74 (35%) | 49 (36%) |
| Dyslipidemia <sup>c</sup> | 130 (61%) | 77 (56%) |
| Total cholesterol, mg/dL | 172 [149, 195] | 177 [151, 208] |
| High density lipoprotein cholesterol, mg/dL | 51 [41, 63] | 58 [46, 69] |
| Lipid-lowering medication use | 54 (25%) | 31 (23%) |
| Diabetes <sup>d</sup> | 27 (13%) | 16 (12%) |
| Diabetes medication use | 20 (9%) | 12 (9%) |
| eGFR, CKD-EPI, mL/min/1.73m<sup>2</sup> | 89 [74, 103] | 93 [78, 106] |
| Hepatitis C diagnosis | 50 (23%) | 28 (20%) |
| *Cardiovascular Magnetic Resonance* |  |  |
| LV ejection fraction, % | 72 [67, 76] | 72 [68, 76] |
| LV ejection fraction <50% | 4 (2%) | 0 (0%) |
| LV mass indexed by BSA, mg/m<sup>2</sup> | 61.8 [56.8, 68.6] | 61.7 [55.8, 67.0] |
| LV end-diastolic volume indexed by BSA, mL/m<sup>2</sup> | 67.6 [58.0, 77.0] | 64.4 [54.3, 76.2] |
| LA volume indexed by BSA, mL/m<sup>2</sup> | 28.1 [22.1, 35.8] | 26.7 [22.6, 32.9] |
| LA volume indexed by BSA ≥40 mL/m<sup>2</sup> | 32 (15%) | 8 (6%) |
| LA volume indexed by BSA ≥53 mL/m<sup>2</sup> | 6 (3%) | 2 (1%) |
| Late gadolinium enhancement | 81 (38%) | 46 (33%) |
| Extracellular volume fraction, % | 28.9 [26.5, 31.2] | 28.2 [26.1, 29.9] |
| *HIV-Related Factors* |  |  |
| HIV viral load detectable (>50 RNA copies/mL) | 55 (26%) | --- |
| CD4+ T cell count, cells/μL | 605 [400, 815] | --- |
| CD4+ nadir T cell count, cells/μL | 271 [149, 386] | --- |
| History of AIDS diagnosis | 29 (20%) | --- |
| ART use | 188 (88%) | --- |
| Duration of ART, years | 12.8 [5.4, 16.0] | --- |
| Protease inhibitor base | 75 (35%) | --- |
| Non-nucleoside reverse transcriptase inhibitor base | 57 (27%) | --- |
| Integrase strand inhibitor base | 53 (25%) | --- |
| Other ART base | 3 (1%) | --- |

<sup>a</sup> Reported in the five years preceding cardiovascular magnetic resonance imaging (CMR).
<sup>b</sup> Hypertension is defined as use of antihypertensive medication or systolic blood pressure ≥140 mmHg or diastolic blood pressure ≥90 mmHg averaged over the preceding 5 years when available or at the time of CMR study visit.
<sup>c</sup> Dyslipidemia is defined as use of lipid-lowering medication or fasting total cholesterol level ≥200 mg/dL or low-density lipoprotein cholesterol level ≥130 mg/dL or high-density lipoprotein cholesterol level <40 mg/dL or serum triglyceride level ≥150 mg/dL closest to the time of CMR study visit.
<sup>d</sup> Diabetes is defined as use of hypoglycemic medication or fasting serum glucose levels ≥126 mg/dL closest to the time of CMR study visit. Hemoglobin A1C level <6.5% was used to exclude diabetes if fasting glucose levels were not available.
PLWH=people living with HIV; PWOH=people without HIV; IQR=interquartile range, reported as (25<sup>th</sup>, 75<sup>th</sup>) percentiles; AUDIT=Alcohol Use Disorders Identification Test; eGFR=estimated glomerular filtration rate; LV=left ventricular; LA=left atrial; BSA=body surface area; ART=antiretroviral therapy.

### Proteomic Signature of HIV Serostatus

Of 2596 proteins analyzed, plasma abundances of 439 proteins were significantly associated with HIV serostatus following multivariable adjustment (415 positively associated with positive serostatus and 24 inversely associated, FDR<0.05) (**FIGURE 2**). These proteins were statistically enriched for many biological processes, including T cell activation, differentiation, and migration; B cell activation; natural killer (**NK**) cell mediated cytotoxicity; regulation of the mitogen-activated protein kinase (**MAPK**) and extracellular signal-regulated kinase 1/2 (**ERK1/ERK2**) cascades; tumor necrosis factor (**TNF**) signaling; response to interleukin (**IL**)-1; IL-10 production; and interferon-γ (**IFN-γ**) production. The strongest observed association with HIV was that of cytotoxic and regulatory T cell molecule (**CRTAM**). On average, the adjusted plasma abundance of CRTAM was 1.01 SD higher among PLWH compared to PWOH (95% confidence interval [**CI**]: 0.84−1.17; FDR<1.0×10^-18^). Upon exclusion of PLWH with detectable plasma HIV RNA, 414 of these 439 proteins remained significantly associated with positive HIV serostatus (390 positively, 24 inversely); one of the strongest of which was CRTAM (FDR<1.0×10^-18^). See ***Table S3*** for results for all evaluated protein associations with HIV serostatus and ***Table S4*** for enrichment details.

**FIGURE 2.**
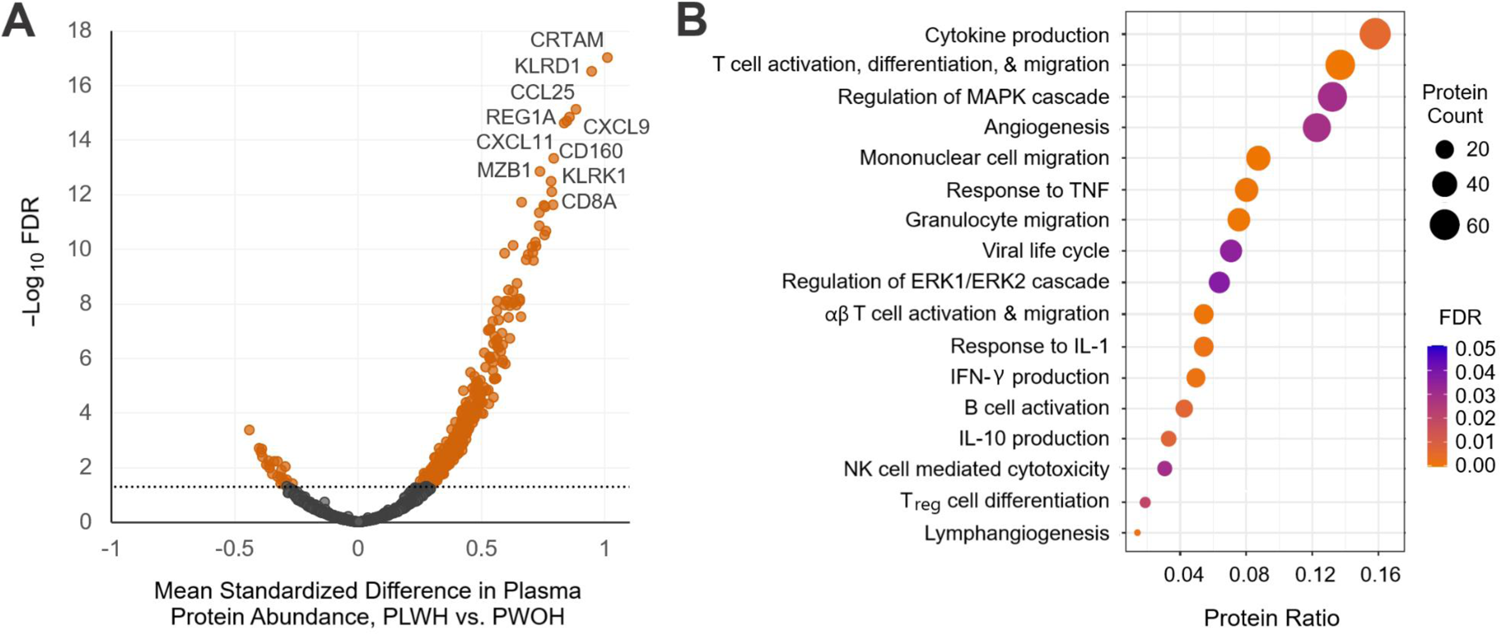
Proteomic signature of HIV seropositivity among people living with and without HIV in the United States (*n*=352). (A) Volcano plot of mean differences in standardized plasma protein abundance comparing PLWH to PWOH vs. -log_10_ false discovery rate (FDR; Benjamini-Hochberg) estimated using linear regression with robust variance, adjusting for age, sex, race/ethnicity, education, pack-years of smoking, hazardous alcohol use, stimulant use in the prior 5 years, opioid use in the prior 5 years, hepatitis C infection, and estimated glomerular filtration rate. Threshold for significance is indicated by gray dotted line, FDR<0.05; 415 of 2596 proteins positively associated with positive HIV serostatus and 24 proteins inversely associated; comparing suppressed PLWH vs. PWOH, 414 of these 439 total proteins were associated with suppressed positive HIV serostatus (not depicted). Complete modeling results can be found in *Supplemental Table S3*. (B) Enriched biological processes among proteins associated with HIV serostatus, estimated using Fisher’s exact test, Gene Ontology: Biological Processes reference database, and a threshold for significance of FDR<0.05. Protein ratio=proportion of proteins significantly associated with HIV serostatus (*n*=439) that map to given annotation. Proteins mapping to each annotation can be found in *Supplemental Table S4*. PLWH=persons living with HIV; PWOH=persons without HIV; suppressed HIV viral load=HIV RNA<50 copies/µL

Many classical markers reflecting chronic immune activation and systemic inflammation among PLWH were significantly elevated among PLWH compared to PWOH, including the monocyte activation markers soluble CD14 (FDR=2.60×10^-10^) and CD163 (FDR=0.009), as well as CXCL10 (FDR=1.73×10^-8^) and TNFR1 (FDR=0.014). Notably, IL-6 levels did not differ by HIV serostatus (*p*=0.19, FDR=0.42), consistent with previously reported immunoassay results among this study population.^24^ These inferences were unchanged following exclusion of PLWH with detectable plasma HIV RNA.

Of six protein clusters agnostically identified using WGCNA (details in ***Figure S2*** and ***Tables S5-S6***), higher plasma abundance of one cluster (Brown) was independently associated with positive HIV serostatus (*p*=3.99×10^-6^), and this association remained following exclusion of PLWH with detectable plasma HIV RNA (*p*=1.02×10^-4^) (*Table S7A-B*). This cluster comprised 42 proteins enriched for regulation of T cell proliferation, TNF signaling, ephrin signaling, cell-cell adhesion, and extracellular matrix (**ECM**) organization (***Table S8***).

### HIV-Associated Proteomic Signature of Left Atrial Remodeling

Next, we identified proteome features significantly associated with both HIV serostatus and LAVi that may reflect potential contributors to HIV-associated LA remodeling. Of 439 proteins associated with HIV serostatus, 78 were also associated with LAVi (74 positively, 4 inversely), adjusting for demographics, substance use, HIV, HCV, and renal function. Upon further adjustment for traditional CVD risk factors, 73 remained associated (69 positively, 4 inversely) (**FIGURE 3A, *Table S9***), all of which were associated with concordant directionality compared to positive HIV serostatus (**FIGURE 3B**). In subgroup analyses, 72 (99%) of these proteins differed in mean plasma level between PLWH with undetectable plasma HIV RNA and PWOH. These proteins were statistically enriched for involvement in T cell activation, differentiation, and proliferation; B cell activation; NK cell-mediated cytotoxicity; cell-cell adhesion; and TNF signaling, amongst others (**FIGURE 3C**, ***Table S10***).

**FIGURE 3.**
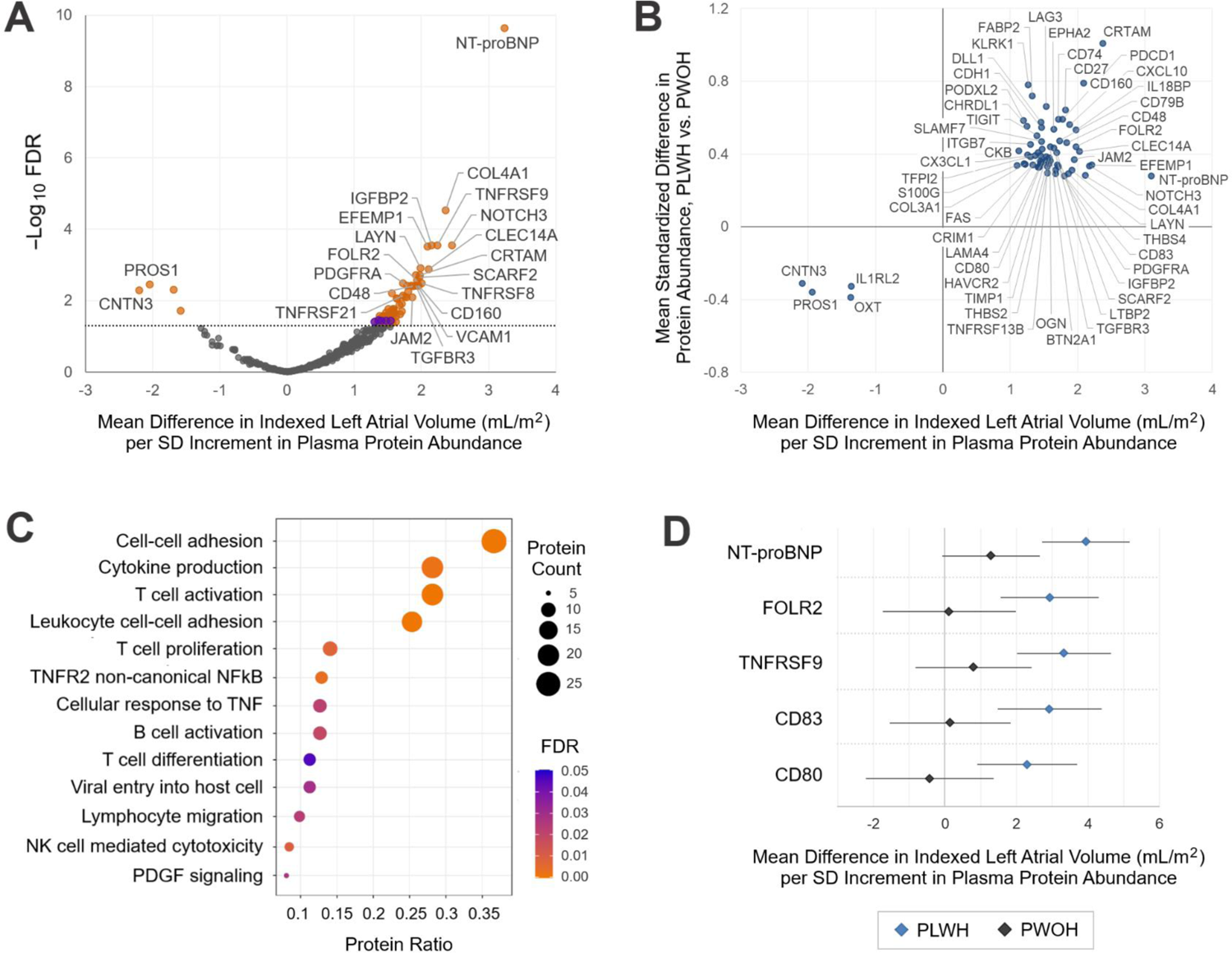
HIV-associated proteomic signature of left atrial size among people living with and without HIV in the United States (*n*=352). (A) Volcano plot of mean differences in left atrial volume indexed for body surface area (LAVi) per standard deviation (SD) increment in plasma abundance of 2594 individual proteins vs. -log_10_ false discovery rate (FDR; Benjamini-Hochberg) estimated using linear regression with robust variance. Purple indicates proteins only associated when adjusting for age, sex, race/ethnicity, HIV, hepatitis C (HCV) infection, and estimated glomerular filtration rate (eGFR). Orange indicates proteins associated with further adjustment for education, body mass index (BMI), systolic blood pressure (SBP), anti-hypertensive medication, diabetes, dyslipidemia, current hazardous alcohol use, pack-years of smoking in prior 5 years, stimulant use in prior 5 years, and opioid use in prior 5 years. The threshold for significance is indicated by gray dotted line, FDR<0.05. Most saturated model (orange) yielded 69 proteins positively associated with LAVi and 4 proteins inversely associated. Complete modeling results can be found in *Supplemental Table S9*. (B) Enriched biological processes among proteins associated with both positive HIV serostatus and higher LAVi, same directionality, estimated using Fisher’s exact test, Gene Ontology: Biological Processes reference database, and a threshold for significance of FDR<0.05. Protein ratio=proportion of total HIV- and LAVi-associated proteins (*n*=73) that map to given annotation. Proteins mapping to each annotation can be found in *Supplemental Table S10*. TNF=tumor necrosis factor; NK=natural killer; PDGF=platelet derived growth factor. (C) Beta-beta plot of mean differences in LAVi per SD increment in individual plasma protein abundances vs. mean differences in standardized protein abundances comparing persons living with vs. without HIV (PLWH, PWOH). All associations were estimated using linear regression with robust variance, and proteins depicted (*n*=73) are restricted to those significantly associated with both parameters with a FDR<0.05. HIV point estimates (y-axis) were adjusted for age, sex, race/ethnicity, education, current hazardous alcohol use, pack-years of smoking in prior 5 years, stimulant use in prior 5 years, opioid use in prior 5 years, HCV, and eGFR. LAVi point estimates (x-axis) were adjusted for the same covariates, as well as BMI, SBP, anti-hypertensive medication, dyslipidemia, and diabetes. (D) Mean differences in LAVi per SD increment in individual plasma protein abundances among strata of PLWH and PWOH, estimated using linear regression with robust variance adjusting for age, sex, race/ethnicity, education, BMI, SBP, anti-hypertensive medication, dyslipidemia, diabetes, current hazardous alcohol use, pack-years of smoking in prior 5 years, stimulant use in prior 5 years, opioid use in prior 5 years, HCV, and eGFR. Proteins depicted are limited to those with HIV×protein multiplicative interactions with a FDR<0.10 (*n*=73 proteins tested).

The strongest observed protein association with LAVi was that of known HF biomarker, N-terminal pro-B type natriuretic peptide (**NT-proBNP**). On average, LAVi was 3.09 mL/m^2^ higher (11% of mean LAVi) per SD increment in plasma NT-proBNP (95% CI: 2.16−4.03; FDR=7.75×10^-9^). Notably, among the top 5 strongest associations with LAVi was CRTAM, which also had the strongest association with HIV serostatus; LAVi was 2.37 mL/m^2^ higher on average per SD (95% CI: 1.27−3.47; FDR=3.40×10^-4^). The 73 proteins associated with LAVi exhibited pairwise correlations in plasma abundances that ranged widely from weak to strong (***Figure S3***) with several moderate confidence protein-protein interactions detected (***Figure S4***).

Next, we determined what proportion of the multivariable adjusted association between HIV and LAVi was accounted for by further adjustment for each identified proteome feature. When further adjusting the HIV-LAVi analysis for each of these 73 proteins separately (in addition to demographics, substance use, HCV, eGFR, and CVD risk factors), 38 proteins reduced the independent HIV association by >30%, and 8 reduced it by >50%—those being CRTAM, CD27, CD79B, CD160, TNF receptor superfamily (**TNFRSF**) 8, TNFRSF9, TNFRSF17, and IL18BP (***Table S11***). Notably, adjustment for CRTAM reduced the association between positive HIV serostatus and higher LAVi by 82%, suggesting it may substantially account for that observed association.

We evaluated these 73 proteins for differences in their adjusted LAVi associations by HIV serostatus and found no significant protein × HIV interactions (FDR<0.05). Given type II error is likely high for these multiplicative tests, we present interactions with higher significance thresholds to broaden hypothesis-generating potential. Five proteins exhibited associations with LAVi that differed by HIV serostatus at FDR<0.10 and were present only among PLWH: NT-proBNP; CD80, CD83, and TNFRSF9 (CD137), all important immune checkpoint co-stimulatory molecules;^25–27^ and folate receptor β (**FOLR2**), an important folate binding protein expressed by myeloid cells, including infiltrating M2-like macrophages^28^ (**FIGURE 3D**). See ***Table S12*** for results of all 73 individual protein interaction tests alongside modeling results stratified by HIV serostatus. We detected no protein × age interactions (***Table S13***).

In our multi-analyte approach, we found higher plasma level of the one agnostically defined HIV-associated protein cluster (Brown) was also associated with higher LAVi, adjusting for age, sex, race/ethnicity, eGFR, HCV, and HIV serostatus (2.03 mL/m^2^ higher per SD increment in Brown cluster plasma level, 95% CI: 0.85−3.21, *p*=0.001, **FIGURE 4**). Following further adjustment for substance use and traditional CVD risk factors, the estimate of association was only mildly attenuated (1.96 mL/m^2^ higher per SD, 95% CI: 0.75−3.16, *p*=0.002). We found adjustment for this protein cluster reduced the independent association between positive HIV serostatus and higher LAVi by 36%. Moreover, this association differed by HIV serostatus (*p*=0.006) and was only present among PLWH. We did not detect an interaction with age (*p*=0.18).

**FIGURE 4.**
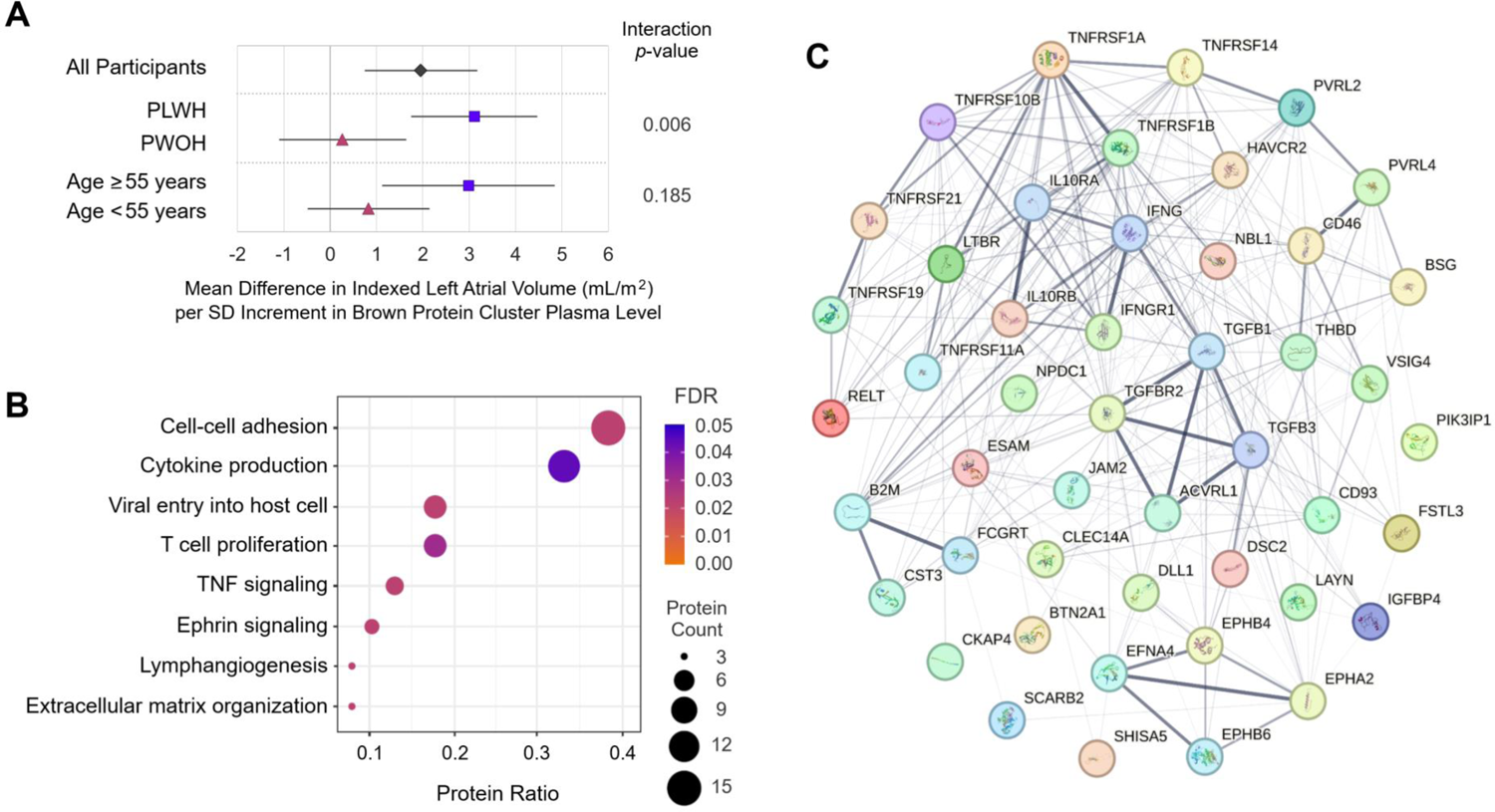
Relationship between an HIV-associated, agnostically defined cluster of plasma proteins and left atrial size among people living with and without HIV in the United States (*n*=352). (A) Mean difference in indexed left atrial volume (LAVi) per standard deviation (SD) increment in ‘Brown’ protein cluster plasma abundance among all participants, among strata of participants living with and without HIV, and among strata of participants above and below median age in SMASH (55 years), estimated using linear regression with robust variance adjusting for age, sex, race/ethnicity, education, body mass index, systolic blood pressure, anti-hypertensive medication, dyslipidemia, diabetes, current hazardous alcohol use, pack-years of smoking in prior 5 years, stimulant use in prior 5 years, opioid use in prior 5 years, hepatitis C infection, and estimated glomerular filtration rate. (B) Enriched biological processes among 42 proteins comprising the ‘Brown’ protein cluster, estimated using Fisher’s exact test, Gene Ontology: Biological Processes reference database, and a threshold for significance of Benjamini-Hochberg false discovery rate (FDR)<0.05. Protein ratio=proportion of total ‘Brown’ cluster proteins (*n*=42) that map to given annotation. Proteins mapping to each annotation can be found in *Supplemental Table S8*. (C) Interaction network of 42 proteins comprising the ‘Brown’ protein cluster generated using STRING, a public database of known and predicted protein-protein interactions. Interactions include direct (physical) and indirect (functional) associations derived from computational prediction, knowledge transfer between organisms, and interactions aggregated from other databases. Line thickness indicates strength of data support. Szklarczyk D, et al. The STRING database in 2023: protein–protein association networks and functional enrichment analyses for any sequenced genome of interest. Nucleic Acids Res. 2023;51(D1):D638-64

Eleven proteins were identified in both single- and multi-analyte approaches—specifically butyrophilin subfamily 2 member A1 (**BTN2A1**), C-type lectin domain containing 14A (**CLEC14A**), delta-like canonical notch ligand 1 (**DLL1**), ephrin A2 (**EPHA2**), endothelial cell adhesion molecule (**ESAM**), T cell immunoglobulin and mucin-domain containing-3 (**TIM-3** or **HAVCR2**), junctional adhesion molecule 2 (**JAM2**), layilin (**LAYN**), TNFRSF1B, TNFRSF14, and TNFRSF21.

### Associations Between Proteomic Signature and Clinical Variables in SMASH

Many components of the identified protein signature were associated with age (*p*<0.05), including the Brown cluster and 26 of 73 individual proteins. Notably, higher plasma CRTAM was not associated with traditional CVD risk factors (age, BMI, hypertension, dyslipidemia, diabetes) but was associated with substance use (smoking and stimulant use) and many HIV-related factors (HCV, detectable plasma HIV RNA, lower current and nadir CD4+ T cell counts, not receiving ART, and shorter ART duration). Similarly, the Brown protein cluster was largely only associated with substance use and HIV-related clinical factors—smoking, opioid use, HCV, detectable plasma HIV RNA, and lower current and nadir CD4+ T cell counts. See ***Figure S5*** for results of all evaluated protein associations with demographic and clinical characteristics.

### External Validation of Protein Signatures with Indexed Left Atrial Volume in MESA

A total of 1242 MESA participants had both CMR imaging and Olink Explore 3072 proteomic data (***Figure S1B***), demographic and clinical characteristics of whom are summarized in **TABLE 2**. Participants were mean (SD) age 67 (9) years, 52% female, and had predominantly normal LVEF.

**TABLE 2.**
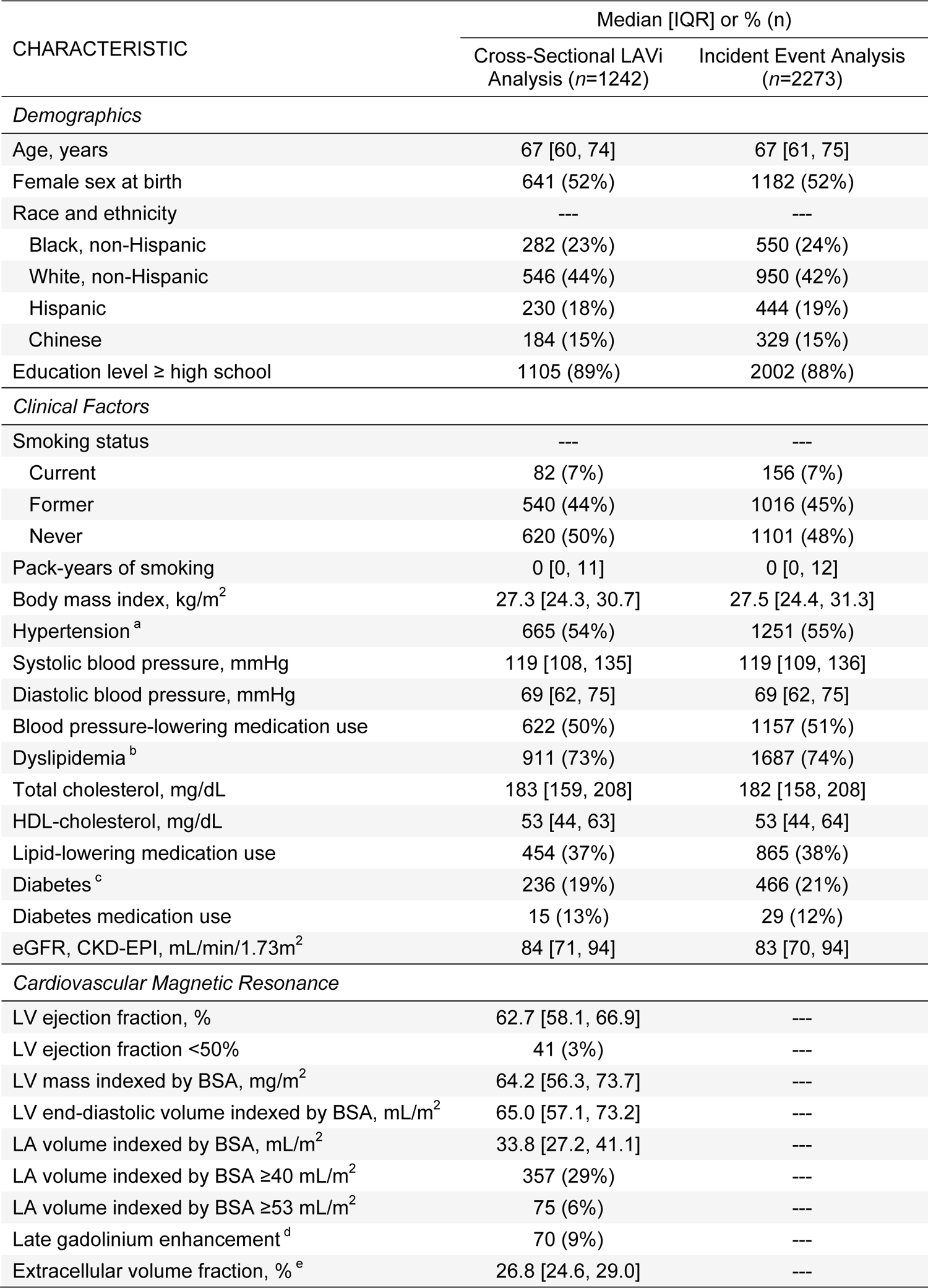
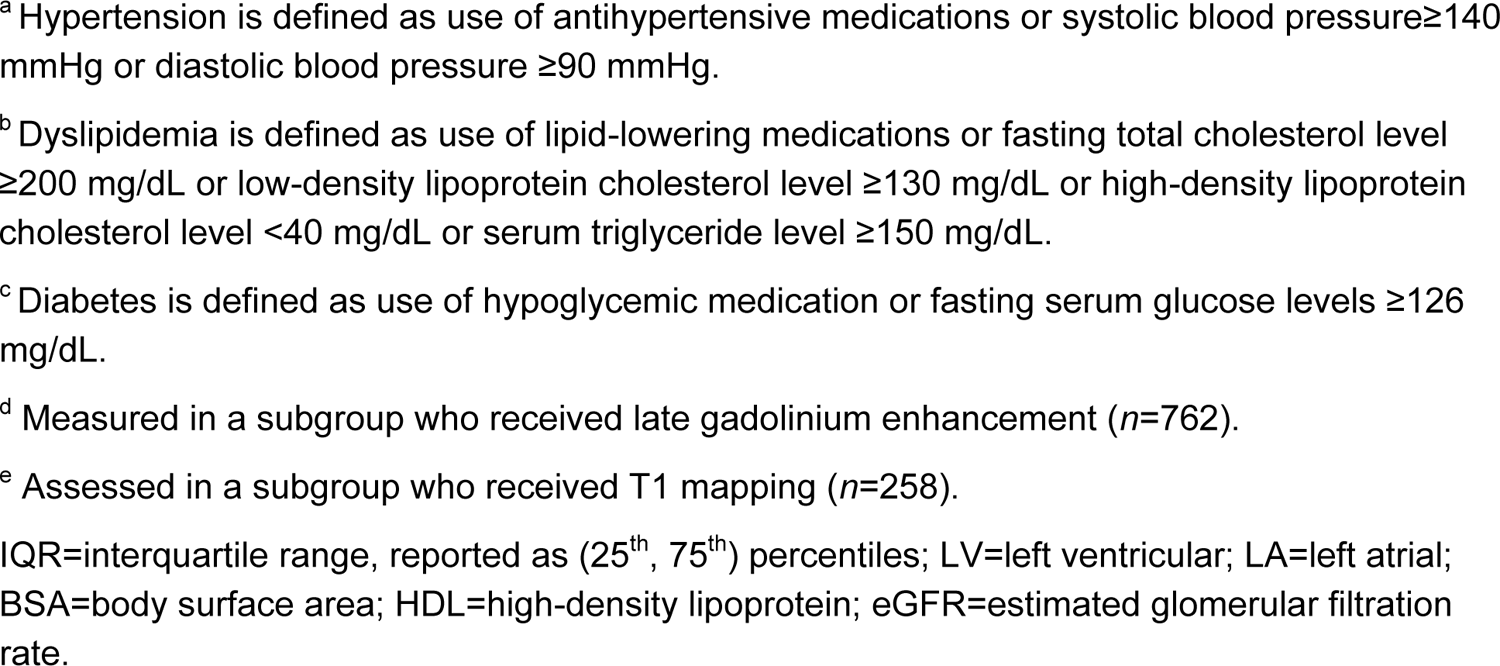
MESA external validation participant characteristics (2010-2012)

| CHARACTERISTIC | Median [IQR] or % (n) |  |
| --- | --- | --- |
|  | Cross-Sectional LAVi Analysis (n=1242) | Incident Event Analysis (n=2273) |
| *Demographics* |  |  |
| Age, years | 67 [60, 74] | 67 [61, 75] |
| Female sex at birth | 641 (52%) | 1182 (52%) |
| Race and ethnicity | --- | --- |
| Black, non-Hispanic | 282 (23%) | 550 (24%) |
| White, non-Hispanic | 546 (44%) | 950 (42%) |
| Hispanic | 230 (18%) | 444 (19%) |
| Chinese | 184 (15%) | 329 (15%) |
| Education level ≥ high school | 1105 (89%) | 2002 (88%) |
| *Clinical Factors* |  |  |
| Smoking status | --- | --- |
| Current | 82 (7%) | 156 (7%) |
| Former | 540 (44%) | 1016 (45%) |
| Never | 620 (50%) | 1101 (48%) |
| Pack-years of smoking | 0 [0, 11] | 0 [0, 12] |
| Body mass index, kg/m<sup>2</sup> | 27.3 [24.3, 30.7] | 27.5 [24.4, 31.3] |
| Hypertension<sup>a</sup> | 665 (54%) | 1251 (55%) |
| Systolic blood pressure, mmHg | 119 [108, 135] | 119 [109, 136] |
| Diastolic blood pressure, mmHg | 69 [62, 75] | 69 [62, 75] |
| Blood pressure-lowering medication use | 622 (50%) | 1157 (51%) |
| Dyslipidemia<sup>b</sup> | 911 (73%) | 1687 (74%) |
| Total cholesterol, mg/dL | 183 [159, 208] | 182 [158, 208] |
| HDL-cholesterol, mg/dL | 53 [44, 63] | 53 [44, 64] |
| Lipid-lowering medication use | 454 (37%) | 865 (38%) |
| Diabetes<sup>c</sup> | 236 (19%) | 466 (21%) |
| Diabetes medication use | 15 (13%) | 29 (12%) |
| eGFR, CKD-EPI, mL/min/1.73m<sup>2</sup> | 84 [71, 94] | 83 [70, 94] |
| *Cardiovascular Magnetic Resonance* |  |  |
| LV ejection fraction, % | 62.7 [58.1, 66.9] | --- |
| LV ejection fraction <50% | 41 (3%) | --- |
| LV mass indexed by BSA, mg/m<sup>2</sup> | 64.2 [56.3, 73.7] | --- |
| LV end-diastolic volume indexed by BSA, mL/m<sup>2</sup> | 65.0 [57.1, 73.2] | --- |
| LA volume indexed by BSA, mL/m<sup>2</sup> | 33.8 [27.2, 41.1] | --- |
| LA volume indexed by BSA ≥40 mL/m<sup>2</sup> | 357 (29%) | --- |
| LA volume indexed by BSA ≥53 mL/m<sup>2</sup> | 75 (6%) | --- |
| Late gadolinium enhancement<sup>d</sup> | 70 (9%) | --- |
| Extracellular volume fraction, %<sup>e</sup> | 26.8 [24.6, 29.0] | --- |

<sup>a</sup> Hypertension is defined as use of antihypertensive medications or systolic blood pressure $\geq 140$ mmHg or diastolic blood pressure $\geq 90$ mmHg.
<sup>b</sup> Dyslipidemia is defined as use of lipid-lowering medications or fasting total cholesterol level $\geq 200$ mg/dL or low-density lipoprotein cholesterol level $\geq 130$ mg/dL or high-density lipoprotein cholesterol level $< 40$ mg/dL or serum triglyceride level $\geq 150$ mg/dL.
<sup>c</sup> Diabetes is defined as use of hypoglycemic medication or fasting serum glucose levels $\geq 126$ mg/dL.
<sup>d</sup> Measured in a subgroup who received late gadolinium enhancement ( $n=762$ ).
<sup>e</sup> Assessed in a subgroup who received T1 mapping ( $n=258$ ).
IQR=interquartile range, reported as (25<sup>th</sup>, 75<sup>th</sup>) percentiles; LV=left ventricular; LA=left atrial; BSA=body surface area; HDL=high-density lipoprotein; eGFR=estimated glomerular filtration rate.

We tested each feature of the HIV-associated proteomic signature of LA remodeling that we identified in SMASH for associations with LAVi in MESA. Among this cohort of older PWOH, 8 of 73 plasma proteins were cross-sectionally associated with LAVi following multivariable adjustment (**TABLE 3, *Table S14***). Following NT-proBNP, the strongest validated association was that of CLEC14A (mean difference: 1.36 mL/m^2^ higher per SD, 95% CI: 0.65−2.07, FDR=0.006), followed by collagen type IV alpha 1 chain (**COL4A1**) (1.20 mL/m^2^ higher per SD, 95% CI: 0.56−1.85, FDR=0.006). Moreover, 28 (of 73 proteins) had LAVi associations that differed by age (interaction FDR<0.05, 41 proteins FDR<0.10) (***Table S15***). Sixteen of these proteins that differed by age had significant associations with LAVi only among a subgroup of MESA participants older than median age at exam 5 (≥67 years), and only one (NT-proBNP) was associated among participants age <67 years (**TABLE 3**). Furthermore, plasma level of the Brown protein cluster was only associated with LAVi among older MESA participants (age ≥67 years, mean difference in LAVi per SD: 1.23 mL/m^2^, 95% CI: 0.03−2.43, *p*=0.04; age <67 years: -0.72, 95% CI: −1.75−0.32, *p*=0.17; interaction *p*=0.005).

**TABLE 3.** Summary of cross-sectional associations between identified HIV-associated proteomic signature of left atrial size and indexed left atrial volume by age among people without HIV in MESA (*n*=1242)

| Analysis Sample | Mean (SD) Age | Proteins FDR<0.05 |
| --- | --- | --- |
| All participants | 68 (9) | CKB, **CLEC14A**, CNTN3, **COL4A1**, IGFBP2, **NOTCH3**, **NT-proBNP**, **PDGFRA** |
| Participants ≥ median age | 75 (5) | **B3GNT7**, **CLEC14A**, **COL4A1**, **CRIM1**, **CX3CL1**, **DCTPP1**, **EPHA2**, **LTBP2**, **NOTCH3**, **NT-proBNP**, **PDGFRA**, **SCARF2**, **THBS2**, **THBS4**, **TIMP1**, **VCAM1**,<br>Brown protein cluster |
| Participants < median age | 60 (4) | **NT-proBNP** |

### External Validation of Protein Signatures with Incident Clinical Events in MESA

Incident clinical event analyses did not require CMR data and were performed in a larger subgroup of the MESA cohort to maximize statistical power (***Figure S1C***). Distributions of demographic and clinical characteristics were similar to those in cross-sectional analyses presented above (**TABLE 2**).

Among 2185 MESA participants without known AF at exam 5, there were 252 incident AF events over a median [IQR] follow-up of 7.8 [0.9] years. Only NT-proBNP (of 73 proteins) was associated with incident AF (***Table S16*** *and **Figure S6**)* following multivariable adjustment (HR: 1.54 per SD increment in NT-proBNP, 95% CI: 1.33−1.77, FDR=2.08×10^-7^). Plasma abundance of the Brown protein cluster was also associated with incident AF (HR: 1.18 per SD increment, 95%CI: 1.01 to 1.37, *p*=0.036). No interactions with baseline age were detected (all FDR>0.10).

Among 2273 MESA participants without known HF at exam 5, there were 54 incident adjudicated clinical HF events over a median [IQR] follow-up of 8.9 [0.8] years. Thirty of 73 proteins were associated with incident HF (**FIGURE 5**) following adjustment, the strongest of which was TNFRSF1B (HR: 1.81 per SD increment, 95% CI: 1.41−2.33, FDR=2.58×10^-4^). Five of these proteins were also associated with LAVi in MESA when restricting to participants above the median age—CLEC14A, cysteine rich transmembrane BMP regulator 1 (**CRIM1**), NT-proBNP, thrombospondin-2 (**THBS2**), and tissue inhibitor matrix metalloproteinase-1 (**TIMP1**).

**FIGURE 5.**
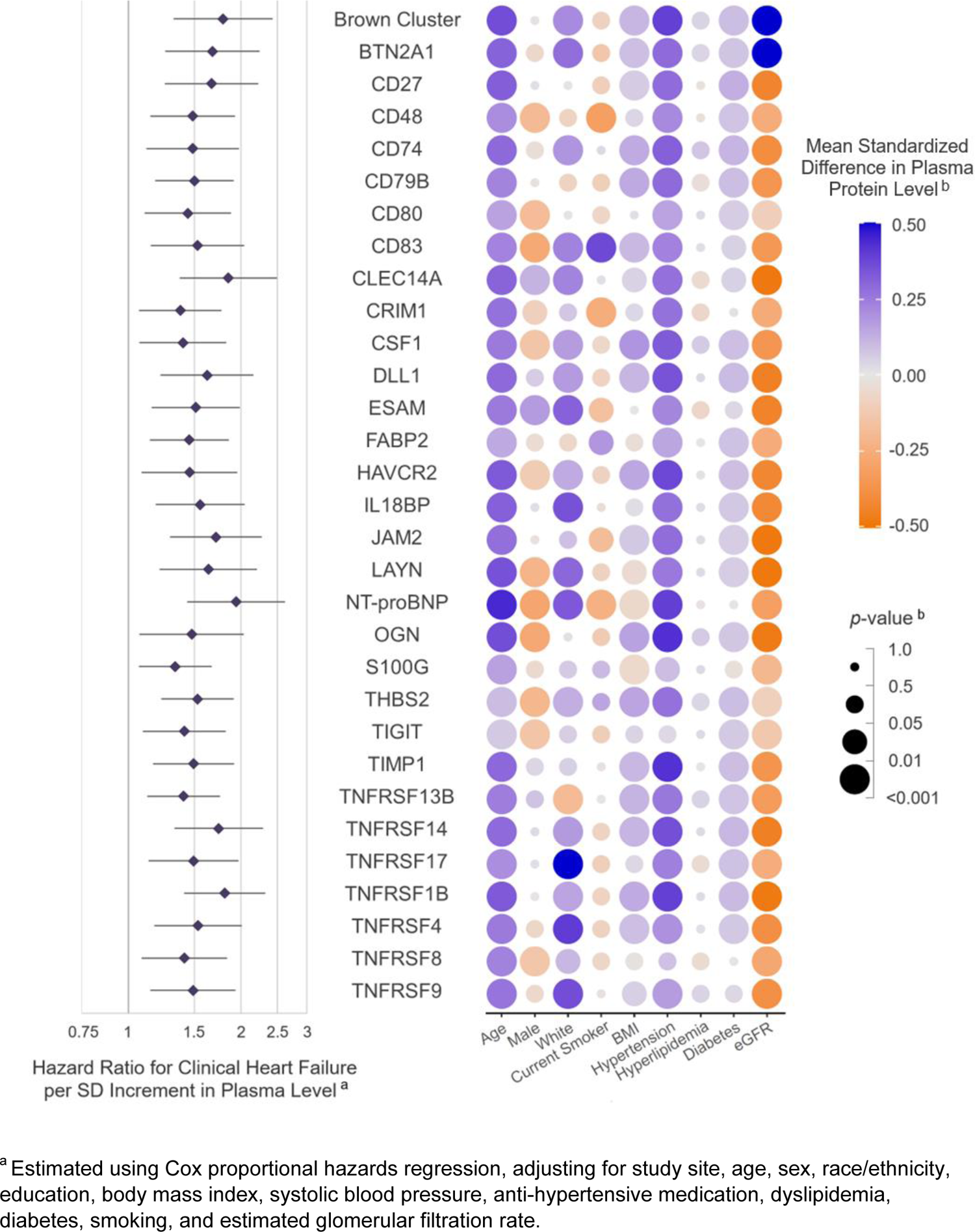
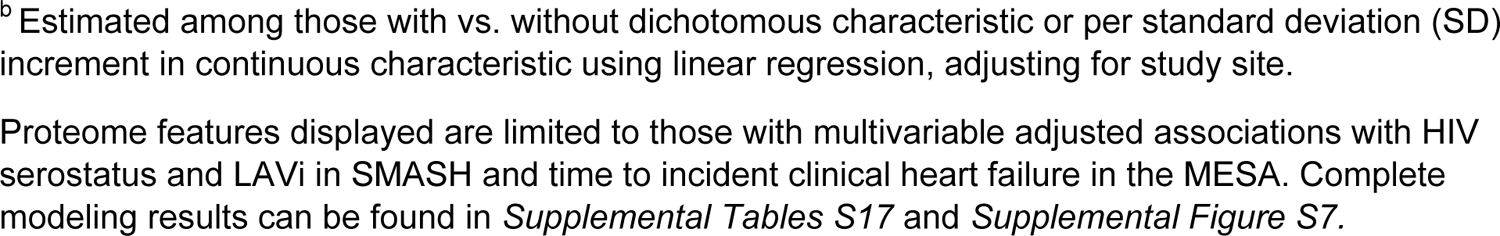
Association between identified HIV-associated proteomic signature of left atrial size and time to incident adjudicated clinical heart failure among people without HIV in MESA (*n*=2273).

See ***Table S17*** and ***Figure S7*** *for* results of all evaluated protein associations with time to incident HF. The hazard of HF was also significantly greater with higher plasma level of the Brown protein cluster (HR: 1.79 per SD, 95% CI: 1.32 to 2.43, *p*=1.82×10^-4^). No interactions with age were detected (all FDR>0.10). Notably, 9 of the 11 proteins identified in both single- and multi-analyte discovery analyses in SMASH were associated with incident HF in MESA— BTN2A1, CLEC14A, DLL1, ESAM, HAVCR2, JAM2, LAYN, TNFRSF1B, and TNFRSF14.

### Associations Between Validated Proteomic Signature Features and Clinical Variables

Every component of the protein signature validated with clinical HF among MESA participants was positively and strongly associated with age (all *p*<0.001) (**FIGURE 5**). Many components were also strongly associated with hypertension (28 proteins and Brown cluster), diabetes (25 proteins and Brown cluster), and higher BMI (22 proteins and Brown cluster)—which remained associated when adjusted for eGFR.

## DISCUSSION

In this discovery study, we identified a plasma proteomic signature representing potential novel biological contributors to the association between HIV infection and left atrial remodeling—a marker of subclinical heart disease. Importantly, this signature was strongly associated with age and higher risk for incident clinical HF among a large independent cohort of PWOH who were over 10 years older. This proteomic signature comprised 73 individual proteins and one agnostically-defined cluster of 42 proteins, largely comprising immune checkpoint proteins (**ICPs**), cytokine signaling, ephrin signaling, and ECM organization pathways. Our findings support the premise that persistent immune activation, higher systemic inflammation, and fibrosis contribute to HIV-associated atrial remodeling, even among PLWH on ART with viral suppression. Furthermore, these same mechanisms may also contribute to aging-related HF among PWOH.

### Aging, HIV and Heart Failure Converge on Inflammation and Immune Activation

It is well-known that PLWH exhibit elevated systemic inflammation and immune activation compared to PWOH, characterized by T cell exhaustion, macrophage activation, and elevated levels of circulating pro-inflammatory cytokines that persist even among virally suppressed PLWH on ART. Immunosenescence is a well-described consequence of aging, and maladaptively activated innate and adaptive immune responses have also been implicated in acute and chronic HF.^29^ This is the prevailing hypothesized mechanism through which PLWH experience excess risk of aging-related diseases, including HF, and our findings support and expand upon those previously put forward.

### Proteins Associated with HIV, Left Atrial Remodeling, and Incident Clinical HF

We identified 30 proteins associated with HIV serostatus and LAVi in SMASH and incident clinical HF among older PWOH in MESA, many of which are novel. Five of these proteins were associated with LAVi among PWOH in MESA when restricting to participants above median age. Interestingly, this list also included several proteins exhibiting especially high potential for mediating and/or moderating effects on the association between HIV and atrial remodeling as well as 9 proteins identified in both single- and multi-analyte discovery analyses. This overlap in identification is important, because these approaches are both agnostic and independent of each other; identification by both approaches therefore strengthens the plausibility of the identified proteins as potential contributors to both HIV- and aging-related LA remodeling and HF.

Many of these proteins of especially high interest are important ICPs—specifically CD27, TNFRSF8 (CD30), CD79B, CD80, TNFRSF9 (CD137), HAVCR2 (TIM-3), TNFRSF14 (HVEM), TIGIT, TNFRSF1B (TNFR2), and less extensively studied BTN2A1, CD83, IL18BP, and TNFRSF17. Many of the remaining proteins—CLEC14A, CRIM1, DLL1, ESAM, JAM2, LAYN, THBS2, and TIMP1—are involved broadly in cell migration and tissue homeostasis.

### Immune Checkpoint Proteins

Activation of T cells, B cells, and NK cells is regulated by several costimulatory and inhibitory ICPs. Elevated expression of ICPs is a known consequence of HIV infection that persists despite ART and contributes to immune dysfunction, HIV disease progression, and viral persistence, detailed recently.^30^ Many of the ICPs we identified belong to the TNF receptor superfamily, upregulation of which is both a hallmark of HIV-associated inflammation and immune activation^31^ and has been extensively observed in HF.^32^ Higher soluble CD27 and CD80 have also recently been identified as correlates of risk for non-AIDS adverse events among PLWH receiving ART.^33^ Therapeutic blockade of ICPs has, however, proven controversial and dependent on whether target molecules serve co-stimulatory or co-inhibitory functions. Taken together with the unique features of HIV-associated immune dysfunction, this highlights the need for focused future investigation of ICPs in HIV-associated cardiac remodeling and HF.

### Cell Migration and Tissue Homeostasis

In this study, the one agnostically defined protein cluster of interest included four proteins involved in ephrin signaling, one of which was also identified in single-analyte discovery analyses (EPHA2). Ephrin signaling mediates a wide range of cellular processes involved in tissue homeostasis and response to injury, including angiogenesis and activation/migration of T cells, B cells, and macrophages.^34^ Though ephrin signaling has not been well-studied among PLWH, it has been linked to other viral infections^35^ as well as cardiomyocyte development, systolic and diastolic function, and TGF-β mediated cardiac fibrosis.^36–38^

Several of these higher interest proteins are related to tissue growth and development, angiogenesis, and ECM remodeling—including CRIM1, THBS2, and TIMP1. CRIM1 modulates activity of bone morphogenic proteins, members of the TGFβ superfamily that are known to play roles in cardiac development and disease pathophysiology in HF, pulmonary arterial hypertension, and atherosclerosis.^39^ Both THBS2 and TIMP1 are critical regulators of tissue homeostasis with roles in ECM remodeling, angiogenesis, and cell growth/differentiation, and have been extensively implicated in HF pathogenesis,^40,41^ including in recent plasma proteome studies.^42–44^

Other proteome features of particularly high interest have been linked to migration of various immune cell subsets in tissue homeostasis—specifically DLL1 (a ligand of Notch receptors), ESAM (endothelial cell adhesion molecule), JAM2 (involved in cell migration), and LAYN (involved in focal adhesion). Immune cell recruitment plays an important role in myocardial response to injury;^29^ however, further investigation is needed to clarify the role of these proteins in shared mechanisms of HIV- and aging-related cardiac remodeling and HF.

### CRTAM

We identified potential biological contributors to the association between HIV and LA remodeling. Abundance of one protein—CRTAM—reduced that association by 82%, markedly more than any other protein, and was strongly associated with several clinical factors related to HIV disease severity. CRTAM is an immune activation marker expressed by NK cells, CD8+ T cells and a subset of CD4+ T cells with enhanced cytotoxic activity.^45^ Measures of CRTAM expression have previously been shown to associate with positive HIV serostatus, HIV viral reservoir size, and viral rebound kinetics.^46,47^ It is also believed critical to shaping the gut microbiome and Th17 responses at the mucosa,^48^ increased permeability of which is a consequence of Th17 HIV-susceptibility and a contributor to chronic inflammation among PLWH.^49^ CRTAM was not associated with LAVi or incident HF among older PWOH in MESA, consistent with our hypothesis that it is an HIV-specific contributor to CVD. Taken together, our results and prior work suggest CRTAM may represent a novel HIV-specific marker of cardiovascular risk worthy of further investigation into its prognostic and therapeutic utility.

### Limitations

There are limitations to this study. Protein levels measured with Olink are semi-quantitative, limiting comparison to other assays. Previously published data demonstrated many proteins assessed by Olink Explore 3072 were associated with *cis*-protein quantitative trait loci (pQTLs), suggesting specificity of protein identification.^50^ Nonetheless, all proteins of interest require validation using other methods.

Type II error may be inflated due to the method used for multiple testing correction, which assumes independence of tests (i.e., biological independence of proteins) and is therefore conservative. For example, given macrophage activation is a hallmark of chronic immune activation among PLWH, we hypothesized it would be detected in our enrichment analyses. It was indeed positively associated with HIV and LAVi in SMASH but not following multiple testing correction (*p*=0.03; FDR=0.36).

Discovery analyses are cross-sectional, resulting in temporal ambiguity. There are also limitations of incident event ascertainment in MESA, particularly for AF which relied on hospitalizations and claims data alone, possibly leading to misclassification bias. Finally, though the study populations are sociodemographically diverse, results may not be translatable to unrepresented or underrepresented subpopulations.

## Conclusion

In conclusion, we discovered a novel plasma proteomic signature that may in part reflect or contribute to HIV-associated left atrial remodeling and also predicted incident clinical HF among a large independent cohort of older PWOH. This signature was enriched in pathways of immune activation, cytokine signaling, and extracellular matrix organization. Our findings suggest pathways underlying risk of HF among PLWH may also contribute to HF pathogenesis among older PWOH. If successfully validated in other external populations, these proteins may help refine current HF risk prediction models and could represent novel therapeutic targets for HF among PLWH and PWOH.

## ACKNOWLEDGMENTS

The authors gratefully acknowledge the contributions of the study participants and the dedication of the MWCCS, ALIVE, SMASH, and MESA-TOPMed investigators and staff. Full MWCCS acknowledgement may be found at https://statepi.jhsph.edu/mwccs/acknowledgements/. A full list of participating MESA investigators and institutes can be found at http://www.mesa-nhlbi.org. We would also like to thank Jinshui Fan and Giovanna Fantoni for their help in the Olink proteomic analysis.

## Supplemental Materials

### SUPPLEMENTAL METHODS

#### SMASH (Discovery) Parent Cohort Details

The Multicenter AIDS Cohort Study (MACS) enrollment began in 1984 and occurred at 4 U.S. sites (Baltimore, MD/Washington, DC; Chicago, IL; Pittsburgh, PA; and Los Angeles, CA) over 4 enrollment waves: 1984-1985, 1987-1991, 2001-2003 and 2010-2018.

The Women’s Interagency HIV Study (WIHS) enrollment began in 1994 and recruited women with and without HIV in 4 waves: 1994-1995, 2001-2002, and 2011-2012 from 10 cities (6 original cohort sites were in Brooklyn, NY; the Bronx/Manhattan, NY; Washington, DC; Chicago, IL; San Francisco, CA; and Los Angeles, CA; 4 southern sites were added in 2013: Chapel Hill, NC; Atlanta, GA; Birmingham, AL/Jackson, MS; and Miami, FL. The Los Angeles site discontinued active follow-up in 2013).

MACS and WIHS have since become the MACS/WIHS Combined Cohort Study (https://statepi.jhsph.edu/mwccs/).

AIDS Linked to the Intravenous Experience (ALIVE) enrollment began in 1988. Additional cohort recruitment occurred during 1994–1995, 1998, 2000, and 2005–2008.

#### SMASH (Discovery) Covariate Definitions

*Body mass index*: calculated as weight in kilograms divided by the square of height in meters.

*Chronic active hepatitis C virus infection*: positive antibody and detectable hepatitis C virus ribonucleic acid (RNA).

*Diabetes*: use of hypoglycemic medications or fasting serum glucose levels ≥126 mg/dL closest to the time of CMR study visit. Hemoglobin A1C level <6.5% was used to exclude diabetes if fasting glucose levels were not available.

*Dyslipidemia*: use of lipid-lowering medications or fasting total cholesterol level ≥200 mg/dL or lowdensity lipoprotein (LDL) cholesterol level ≥130 mg/dL or high-density lipoprotein (HDL) cholesterol level <40 mg/dL or serum triglyceride level ≥150 mg/dL closest to the time of CMR study visit.

*Educational level*: did or did not attain a high school diploma.

Estimated glomerular filtration rate (eGFR): calculated using the CKD Epi (2021) equation.

*Hazardous alcohol use*: a score of >8 on the Alcohol Use Disorders Identification Test (AUDIT) assessed at the time of CMR.

*History of cardiovascular disease*: prior myocardial infarction, angioplasty, stent, coronary artery bypass surgery, other heart surgery, or heart failure confirmed by medical records.

*Hypertension*: use of antihypertensive medications or systolic blood pressure≥140 mmHg or diastolic blood pressure ≥90 mmHg averaged over the preceding 5 years when available or at time of cardiac magnetic resonance imaging (CMR) study visit.

*Opioid use*: included any of the following by any administration route over the 5 years preceding

CMR: methadone (including both prescribed and non-prescribed), heroin, speedball, opioid-based pain medications, prescribed and non-prescribed.

#### *Pack-years of smoking*: assessed over the 5 years preceding CMR

*Stimulant use*: included any of the following by any administration route over the 5 years preceding

CMR: cocaine including crack, speedball, crystal methamphetamine, Phenmetrazine (Preludin), benzedrine, methadrine, uppers, speed, methylphenidate (Ritalin), (dextroamphetamine) Dexedrine, amphetamine/dextroamphetamine (Adderall).

#### MESA (External Validation) Cohort Details and Methods

Elements of the primary analysis were externally validated in the Multi-Ethnic Study of Atherosclerosis (MESA) (https://www.mesa-nhlbi.org/). The MESA is a prospective population based cohort initiated in 2000 to study the characteristics and progression of subclinical cardiovascular disease (CVD) among a diverse population in the United States. MESA recruited a total of 6814 participants from six geographic areas across the U.S.—Baltimore City and Baltimore County, Maryland; Chicago, Illinois; Forsyth County, North Carolina; Los Angeles County, California; Northern Manhattan and the Bronx, New York; and St. Paul, Minnesota. At the time of enrollment, participants were 45-84 years of age and had no history of clinical CVD—including coronary artery disease, peripheral vascular disease, cerebrovascular disease, and heart failure.1 The study protocol was approved by Institutional Review Boards of Columbia University, Johns Hopkins University, Northwestern University, University of California Los Angeles, University of Minnesota, and Wake Forest University; and all participants signed informed consent. The present study utilized data on a subset of MESA participants from cohort exam 5 (2010-2012) with follow-up observed through December 31, 2019.

CMR imaging was performed at exam 5 using 1.5-T scanners (Magnetom Avanto and Magnetom Espree, Siemens Medical Systems, Erlangen, Germany) with six-channel anterior and posterior phased-array torso coil elements. The protocol has been described in detail previously (doi:10.1016/j.jcmg.2021.02.014) and included acquisition of one cine horizontal long-axis section (four-chamber view), at least 12 cine short-axis sections from the atria to the cardiac apex, and one cine vertical long-axis section (two-chamber view) using a steady-state free precession pulse sequence.

Proteomics was performed using the Olink Explore 3072 assay on EDTA plasma stored at −80°C using the same standardized laboratory protocol and data quality control, normalization, and calibration methods described in the primary methods and in more detail below. This data was generated through the Trans-Omics for Precision Medicine (TOPMed) program in the laboratory of Dr. Robert Gerzsten.

Information on hospital admissions, outpatient diagnoses, and deaths were ascertained at follow-up telephone interviews conducted every 9-12 months with the participant or a proxy. Medical records and International Classification of Disease (ICD) diagnosis codes were obtained for inpatient and outpatient events, and death certificates were obtained for all deaths.

Medical records for reported heart failure (HF) events were independently reviewed by two physicians. The probable diagnosis of HF required a physician diagnosis, signs or symptoms of HF, and medical treatment for HF. The definite diagnosis of HF also required ≥1 criterion such as pulmonary congestion or edema by chest X-ray; reduced left ventricular (LV) function or dilated LV by echocardiography or ventriculography; or evidence of LV diastolic dysfunction. We included both probable and definite diagnosis of HF. End of follow-up for HF event ascertainment was December 31, 2019.

Incident cases of atrial fibrillation (AF) during the follow-up period were identified through MESA event surveillance and, for participants enrolled in fee-for-service Medicare, from inpatient and outpatient Medicare claims data. AF was documented as present if an International Classification of Diseases, Ninth Revision diagnosis code 427.31 (AF) or 427.32 (atrial flutter) was present. End of follow-up for AF event ascertainment was December 31, 2018.

#### Olink Proximity Extension Assay and Data Quality Control

The Olink Explore 3072 assay was used in both SMASH and MESA. Olink technology has been described in detail previously (doi:10.1371/journal.pone.0095192). Briefly, DNA oligonucleotide-labeled antibody pairs bind target antigen in solution and, when bound pairwise, hybridize and are extended by a DNA polymerase. This forms a new DNA barcode, which is then amplified and quantified by microfluidic quantitative PCR. This technology differs from standard immunoassays in that it does not rely on spectrometry for quantification, and only matched DNA reporter pairs will be amplified, leading to high specificity by mitigating cross-reactive binding.

Data quality control, normalization, and calibration were carried out using the Normalized Protein eXpression (NPX) software at probe, sample, and plate levels. Specifically, data was normalized using multiple internal assay controls used to monitor immunoreaction, extension, and readout steps, and inter-plate sample controls were used to further normalize for inter-plate variation.

The Olink Explore 3072 platform assays 2924 total proteins. Proteins were excluded from analysis if they did not meet Olink batch release standards (*n*=54), did not pass internal or external protein quality control standards (*n*=37), or were detected in <50% of samples (*n*=239).

#### Weighted Gene Co-expression Network Analysis (WGCNA)

We derived unique clusters of highly correlated proteins using the WGCNA package in R (version 4.2) (doi:10.1002/0471142727.mb2010s109, doi:10.1186/1471-2105-9-559). Briefly, unsigned weighted networks of proteins were constructed using pairwise bi-weight midcorrelations of plasma abundance values and a soft threshold transformation that produced an approximately scale-free topology. Next, the topological overlap matrix was calculated and used to hierarchically cluster proteins. A summary plasma abundance measure for each cluster, the eigenprotein, was then derived from the within-network expression correlation matrix using principal component analysis and was used in downstream modeling.

---

**SUPPLEMENTAL TABLE S1.** SMASH participant characteristics by inclusion vs. exclusion from proteomics study.

| CHARACTERISTIC | Median [IQR] or % (n) |  | *p* |
| --- | --- | --- | --- |
|  | OLINK (*n*=352) | No OLINK (*n*=116) |  |
| *Demographics* |  |  |  |
| Age, years | 55 [51, 58] | 55 [50, 59] | 0.671 |
| Female sex at birth | 25.3% (89) | 43.1% (50) | 0.0004 |
| Race and ethnicity | --- | --- |  |
| Black, non-Hispanic | 69.6% (245) | 77.6% (90) | 0.190 |
| White, non-Hispanic | 24.4% (86) | 16.4% (19) |  |
| Hispanic | 6% (21) | 6% (7) |  |
| Education level ≥ high school | 75% (264) | 57% (66) | 0.0003 |
| Annual income ≥\$10,000 | 60.8% (205) | 48.2% (53) | 0.026 |
| *Substance Use* |  |  |  |
| Current smoker | 48.6% (171) | 47.4% (55) | 0.912 |
| Pack-years of smoking <sup>a</sup> | 0.77 [0.00, 2.67] | 0.81 [0.00, 2.72] | 0.849 |
| Hazardous alcohol use (AUDIT score >8) | 12.2% (43) | 9.5% (11) | 0.528 |
| Opioid use <sup>a</sup> | 30.7% (108) | 37.9% (44) | 0.183 |
| Stimulant use <sup>a</sup> | 38.9% (137) | 42.2% (49) | 0.600 |
| *Clinical Factors* |  |  |  |
| History of CVD | 6.2% (22) | 6.0% (7) | 1.000 |
| Body mass index, kg/m<sup>2</sup> | 26.1 [23.5, 30.6] | 28.2 [24.1, 31.8] | 0.014 |
| Hypertension | 51.7% (182) | 72.4% (84) | 0.0001 |
| Systolic blood pressure, mmHg | 126 [119, 135] | 126 [120, 137] | 0.311 |
| BP-lowering medication use | 34.9% (123) | 44.8% (52) | 0.072 |
| Dyslipidemia | 58.8% (207) | 58.6% (68) | 1.000 |
| Total cholesterol, mg/dL | 174 [150, 198] | 173 [152, 198] | 0.975 |
| HDL-cholesterol, mg/dL | 53 [43, 66] | 52 [42, 65] | 0.598 |
| Lipid-lowering medication use | 24.1% (85) | 21.6% (25) | 0.656 |
| Diabetes | 12.2% (43) | 15.5% (18) | 0.449 |
| Diabetes medication use | 9.1% (32) | 14.7% (17) | 0.128 |
| eGFR, CKD-EPI, mL/min/1.73m<sup>2</sup> | 93 [76, 105] | 85 [68, 100] | 0.003 |
| Hepatitis C diagnosis | 22.4% (78) | 39.3% (44) | 0.0007 |
| *Cardiovascular Magnetic Resonance* |  |  |  |
| LV ejection fraction, % | 72.3 [67.6, 75.7] | 73.1 [69.0, 76.3] | 0.194 |
| LV mass indexed by BSA, mg/m<sup>2</sup> | 61.8 [56.4, 68.3] | 59.8 [54.3, 65.2] | 0.077 |
| LV end-diastolic volume indexed by BSA, mL/m<sup>2</sup> | 65.8 [56.9, 76.6] | 66.6 [54.4, 75.1] | 0.323 |
| LA volume indexed by BSA, mL/m<sup>2</sup> | 27.3 [22.3, 35.1] | 29.5 [23.1, 37.6] | 0.200 |
| LA volume indexed by BSA ≥40 mL/m<sup>2</sup> | 11.4% (40) | 18.2% (16) | 0.127 |
| Late gadolinium enhancement | 36.1% (127) | 16% (8) | 0.008 |
| Extracellular volume fraction, % | 28.6 [26.1, 30.4] | 29.2 [27.4, 31.5] | 0.570 |

| *HIV-Related Factors* |  |  |  |
| --- | --- | --- | --- |
| HIV diagnosis | 60.8% (214) | 68.1% (79) | 0.194 |
| HIV viral load detectable (>50 RNA copies/mL) | 25.7% (55) | 35.4% (28) | 0.135 |
| CD4+ T cell count, cells/μL | 605 [400, 815] | 613 [443, 813] | 0.644 |
| CD4+ nadir T cell count, cells/μL | 271 [149, 386] | 261 [141, 483] | 0.929 |
| History of AIDS diagnosis | 20.4% (29) | 23.9% (11) | 0.768 |
| ART use | 88.3% (188) | 87.3% (69) | 0.990 |
| Duration of ART, years | 12.8 [5.4, 16.0] | 9.7 [3.6, 15.0] | 0.134 |
| Protease inhibitor base | 35.2% (75) | 36.7% (29) | 0.921 |
| NNRTI base | 26.8% (57) | 25.3% (20) | 0.921 |
| Integrase strand inhibitor base | 24.9% (53) | 25.3% (20) | 1.000 |
| Other ART base | 1.4% (3) | 0% (0) | 0.684 |

<sup>a</sup> Reported in the five years preceding cardiovascular magnetic resonance imaging.
PLWH=persons living with HIV; PWOH=persons without HIV; IQR=interquartile range, reported as (25<sup>th</sup>, 75<sup>th</sup>) percentiles; AUDIT=Alcohol Use Disorders Identification Test; BSA=body surface area; eGFR=estimated glomerular filtration rate; ART=antiretroviral therapy.

**SUPPLEMENTAL TABLE S2.**
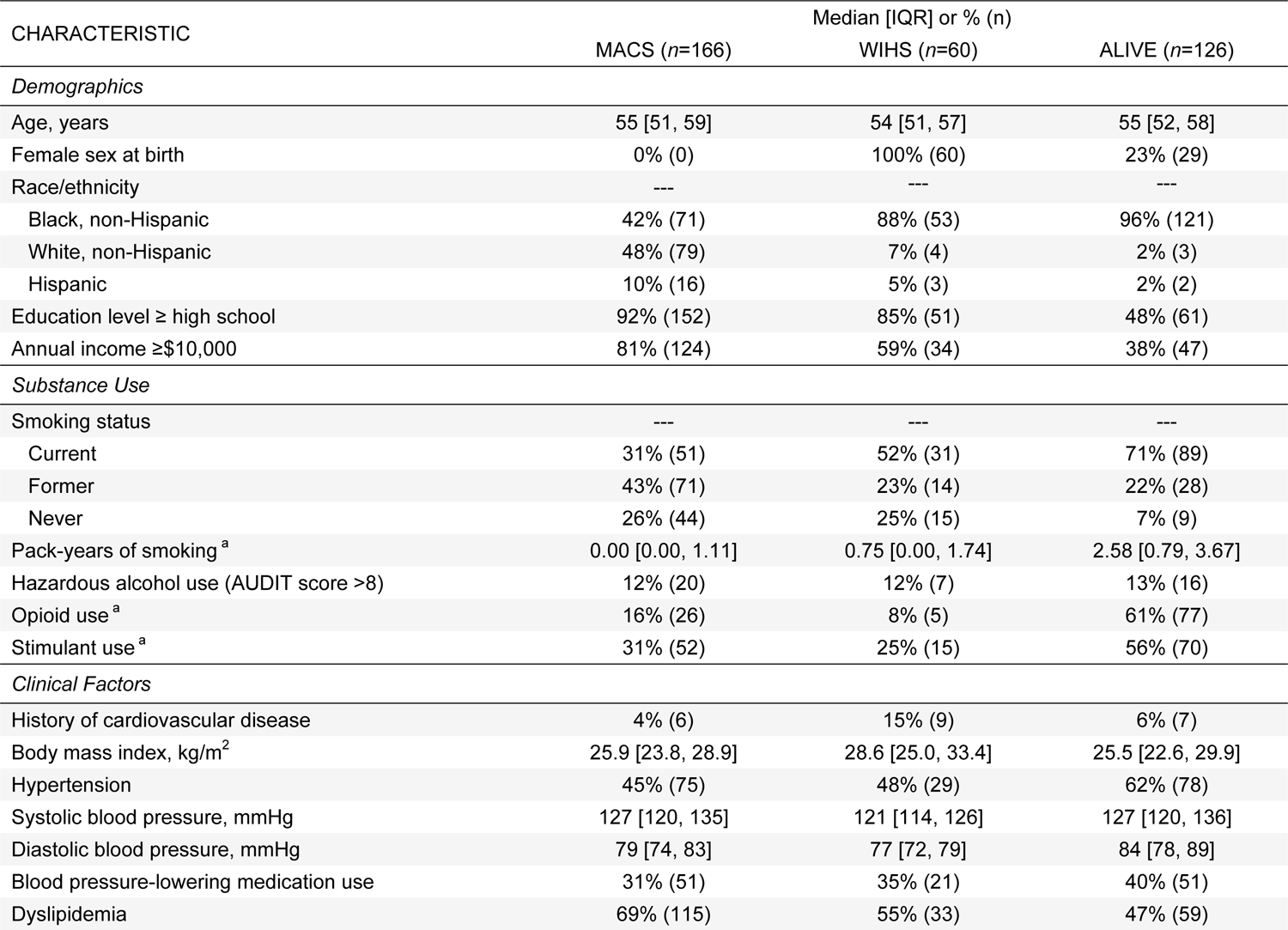
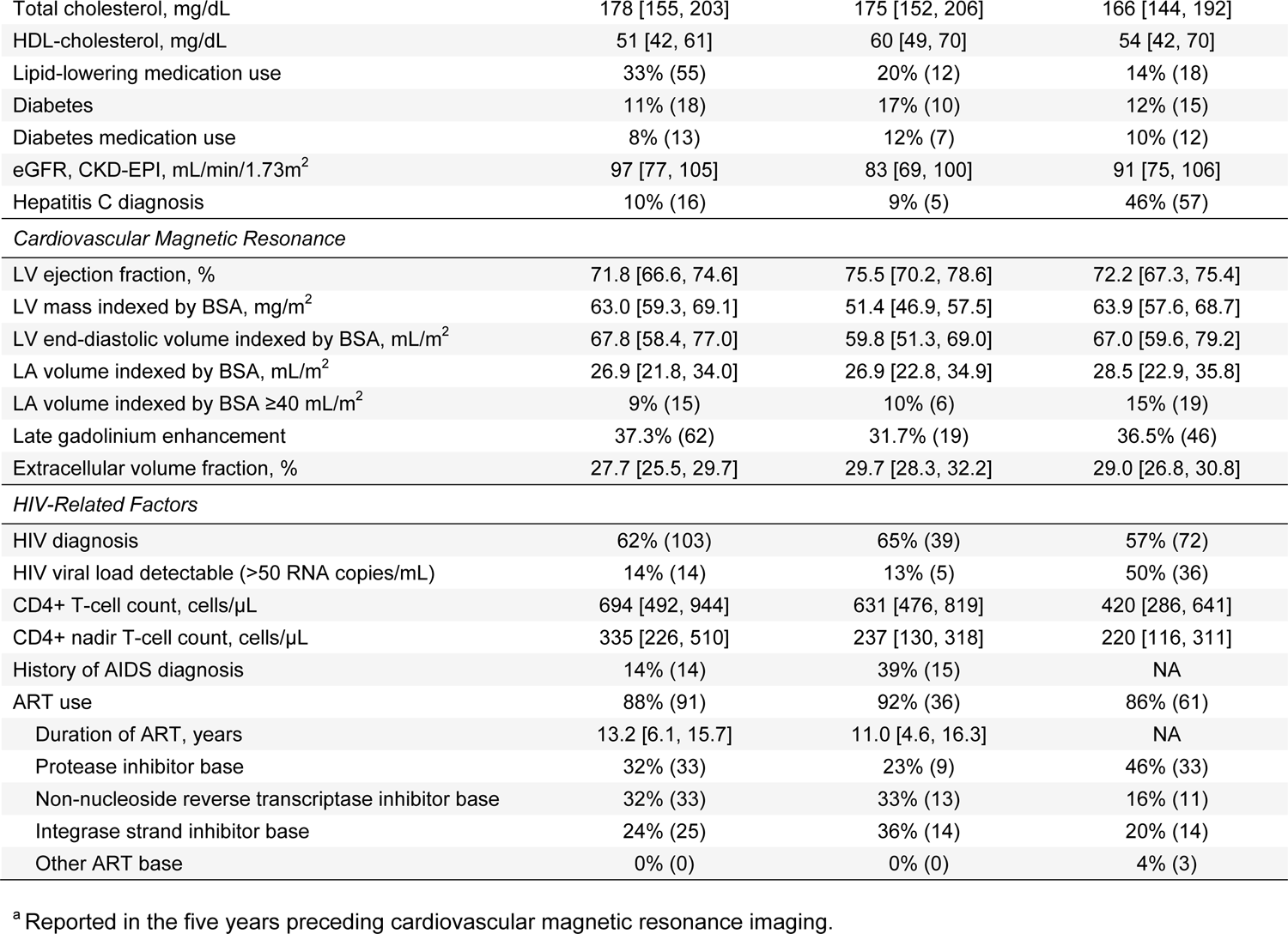
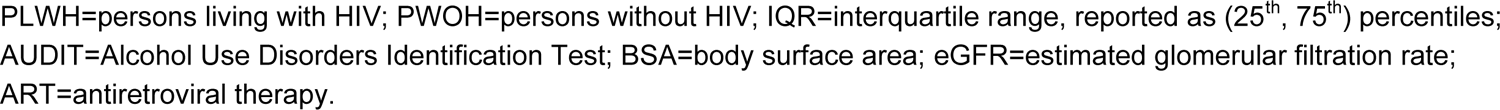
SMASH participant characteristics by parent cohort (*n*=352).

**SUPPLEMENTAL TABLE S3.** Cross-sectional associations between plasma protein abundances and <u>HIV serostatus</u> among (A) PLWH and PWOH and (B) PLWH with undetectable plasma HIV RNA and PWOH in the United States.

See Supplemental Excel file ‘Supplemental Table S3.xlsx’

Mean differences in standardized protein abundance (A) comparing PLWH to PWOH; and (B) comparing PLWH with plasma HIV RNA <50 copies/mL to PWOH, estimated using linear regression with robust variance adjusting for age, sex, race/ethnicity, education, pack-years of smoking, hazardous alcohol use, stimulant use in the prior 5 years, opioid use in the prior 5 years, hepatitis C, and estimated glomerular filtration rate. FDR=Benjamini-Hochberg false discovery rate.

**SUPPLEMENTAL TABLE S4.** Proteins cross-sectionally associated with HIV serostatus corresponding to statistically over-represented biological processes.

| Gene Ontology:<br>Biological Process | Enrichment<br>FDR<sup>a</sup> | Protein<br>Count | Proteins |
| --- | --- | --- | --- |
| Cytokine production | 0.005 | 67 | ACE2, ADGRG1, AXL, B2M, BST2, BTN2A1, BTN3A2, CCN4, CD14, CD160, CD244, CD274, CD28, CD4, CD40, CD46, CD6, CD74, CD80, CD83, CLEC4A, COL3A1, CRTAM, CX3CL1, CXCL6, DLL1, EPHA2, F11R, F2R, FCGR2B, FLT4, HAVCR2, HGF, IFNL1, IL12RB1, IL18, IL1R1, IL1RL2, IL6ST, IL7, INHBB, KLRK1, LAG3, LGALS9, LILRA2, LILRB4, LY9, MDK, NOS2, PDCD1LG2, PGLYRP2, PLA2G10, PLA2G1B, PTPRC, SEMA7A, SLAMF1, SLAMF6, SPON2, TIGIT, TNFRSF14, TNFRSF1B, TNFRSF21, TNFRSF8, TRIM21, VSIG4, VSIR, XCL1 |
| T cell activation, differentiation, and migration | 6.23E-05 | 58 | ADA, B2M, CCL2, CD160, CD27, CD274, CD28, CD300A, CD4, CD46, CD48, CD6, CD7, CD74, CD80, CD83, CD8A, CLEC4A, CRTAM, CTSL, FCGR2B, FGL1, HAVCR2, HLA-DRA, ICAM1, IFNL1, IGFBP2, IL12RB1, IL18, IL1RL2, IL2RA, IL6ST, IL7, ITGAL, KLRK1, LAG3, LGALS9, LILRB4, LY9, MDK, PDCD1LG2, PTPRC, SIRPB1, SLAMF1, SLAMF6, SLAMF7, TGFB2, TIGIT, TNFRSF14, TNFRSF1B, TNFRSF21, TNFRSF4, TNFRSF9, TNFSF13B, VCAM1, VSIG4, XCL1, VSIR |
| Regulation of MAPK cascade | 0.029 | 56 | ACE2, ADAM9, AIDA, ARHGEF5, BMPER, CCL11, CCL14, CCL15, CCL17, CCL2, CCL20, CCL23, CCL25, CCL4, CCL8, CD27, CD300A, CD4, CD40, CD74, CDH2, CX3CL1, EDA2R, EPHA2, F2R, FAS, FCGR2B, FCRL3, FLT4, GDF15, GH1, GPR37, HAVCR2, HGF, ICAM1, IGFBP3, IGFBP4, LGALS9, LILRB4, LTBR, MARCO, MYDGF, NOTCH1, PDGFRA, PLA2G1B, PTPRC, REN, RNF149, ROBO1, SEMA7A, SLAMF1, SMPD1, TNFRSF11A, TNFRSF19, VEGFA, XCL1 |
| Angiogenesis | 0.029 | 52 | ACVRL1, ADGRG1, AMOT, ANGPT2, B4GALT1, BMPER, BSG, CCL11, CCL2, CCN3, CD160, CD40, CDH5, CLEC14A, COL18A1, COL4A1, CX3CL1, CXCL10, CXCL13, CXCL8, DLL1, EPHA2, EPHB4, FLT4, GRN, HGF, HS6ST1, HSPG2, HYAL1, IL18, MDK, MYDGF, NOS3, NOTCH1, NOTCH3, NRCAM, NRP2, PDGFRA, PGF, PTPRM, ROBO1, RSPO3, SCG2, SMOC2, TGFB2, THBS2, THBS4, TIE1, TNFRSF12A, TYMP, VEGFA, VEGFC |
| Mononuclear cell migration | 1.69E-05 | 37 | ARHGEF5, CCL11, CCL14, CCL15, CCL17, CCL2, CCL20, CCL23, CCL25, CCL27, CCL4, CCL8, CCN3, CRTAM, CSF1, CX3CL1, CXCL10, CXCL11, CXCL12, CXCL13, CXCL16, F11R, ICAM1, ITGAL, ITGB7, JAM2, KLRK1, LGALS9, LGMN, MDK, MSTN, NBL1, SLAMF1, SLAMF8, TNFRSF11A, TNFRSF14, XCL1 |
| Response to tumor necrosis factor | 4.34E-04 | 34 | ADAM9, ASAH1, CCL11, CCL14, CCL15, CCL17, CCL2, CCL20, CCL23, CCL25, CCL4, CCL8, CD14, CD40, CX3CL1, CXCL16, CXCL8, EDA2R, FAS, HYAL1, IL18BP, KRT18, SMPD1, TNFRSF11A, TNFRSF14, TNFRSF17, TNFRSF19, TNFRSF1A, TNFRSF1B, TNFRSF21, TNFRSF4, TNFSF13B, VCAM1, XCL1 |
| Granulocyte migration | 1.16E-04 | 32 | ADGRE2, BSG, CCL11, CCL14, CCL15, CCL17, CCL2, CCL20, CCL23, CCL25, CCL4, CCL8, CD300A, CD74, CSF1, CX3CL1, CXCL10, CXCL11, CXCL13, CXCL6, CXCL8, CXCL9, IL1R1, ITGB2, MDK, MSTN, PLA2G1B, SCG2, SLAMF1, SLAMF8, THBS4, XCL1 |
| Viral life cycle | 0.035 | 30 | ACE2, ATG16L1, AXL, BSG, BST2, CCL2, CCL8, CD28, CD4, CD46, CD74, CD80, CTSL, CXCL8, EPHA2, F11R, HAVCR1, ICAM1, ITGB7, LGALS9, NECTIN2, NOTCH1, P4HB, SCARB2, SIGLEC1, SLAMF1, SMPD1, TNFRSF14, TNFRSF4, TRIM21 |
| Regulation of ERK1/ERK2 cascade | 0.037 | 27 | BMPER, CCL11, CCL14, CCL15, CCL17, CCL2, CCL20, CCL23, CCL25, CCL4, CCL8, CD4, CD74, CX3CL1, F2R, FLT4, HAVCR2, ICAM1, LGALS9, MARCO, NOTCH1, PDGFRA, PTPRC, SEMA7A, SLAMF1, TNFRSF11A, XCL1 |
| Response to IL-1 | 4.03E-04 | 23 | CCL11, CCL14, CCL15, CCL17, CCL2, CCL20, CCL23, CCL25, CCL4, CCL8, CD38, CD40, CX3CL1, CXCL8, HYAL1, IL1R1, IL1RL2, INHBB, LGALS9, SMPD1, TNFRSF11A, XCL1, ZBP1 |
| $\alpha\beta$ T cell activation and migration | 0.001 | 23 | ADA, CD160, CD274, CD28, CD300A, CD80, CD83, CLEC4A, CRTAM, CTSL, HLA-DRA, IL12RB1, IL18, IL2RA, LGALS9, LILRB4, LY9, PTPRC, SLAMF6, TGFB2, TNFRSF14, VSIR, XCL1 |
| Interferon-γ production | 0.001 | 21 | AXL, BTN3A2, CD14, CD160, CD244, CD274, CRTAM, HAVCR2, IFNL1, IL12RB1, IL18, IL1R1, KLRK1, LGALS9, LILRB4, PDCD1LG2, PGLYRP2, SLAMF1, SLAMF6, VSIR, XCL1 |
| B cell activation | 0.007 | 18 | ADA, CD27, CD28, CD300A, CD38, CD40, CD74, FCGR2B, FCRL3, IGLC2, IL7, MZB1, PTPRC, SLAMF8, TNFRSF13B, TNFRSF21, TNFRSF4, TNFSF13B |
| IL-10 production | 0.007 | 14 | CD274, CD28, CD46, CD83, DLL1, FCGR2B, HGF, LGALS9, LILRB4, PDCD1LG2, TIGIT, TNFRSF21, XCL1, VSIR |
| Natural killer cell mediated cytotoxicity | 0.030 | 13 | CD160, CRTAM, HAVCR2, IL18, KLRD1, KLRK1, LAG3, LGALS9, NCR1, NECTIN2, SH2D1A, SLAMF6, SLAMF7 |
| T<sub>reg</sub> cell differentiation | 0.020 | 8 | CD28, CD46, HLA-DRA, LAG3, LGALS9, LILRB4, MDK, VSIR |

<sup>a</sup> Estimated using Fisher's exact test, Gene Ontology Biological Processes reference database, and a threshold for significance of Benjamini-Hochberg false discovery rate (FDR)<0.05.
MAPK=mitogen-activated protein kinases; ERK=extracellular signal-regulated kinases.

**SUPPLEMENTAL TABLE S5.** Individual proteins within each of six clusters agnostically defined using weighted gene co-expression network analysis.

See Supplemental Excel file ‘Supplemental Table S5.xlsx’

**SUPPLEMENTAL TABLE S6.**
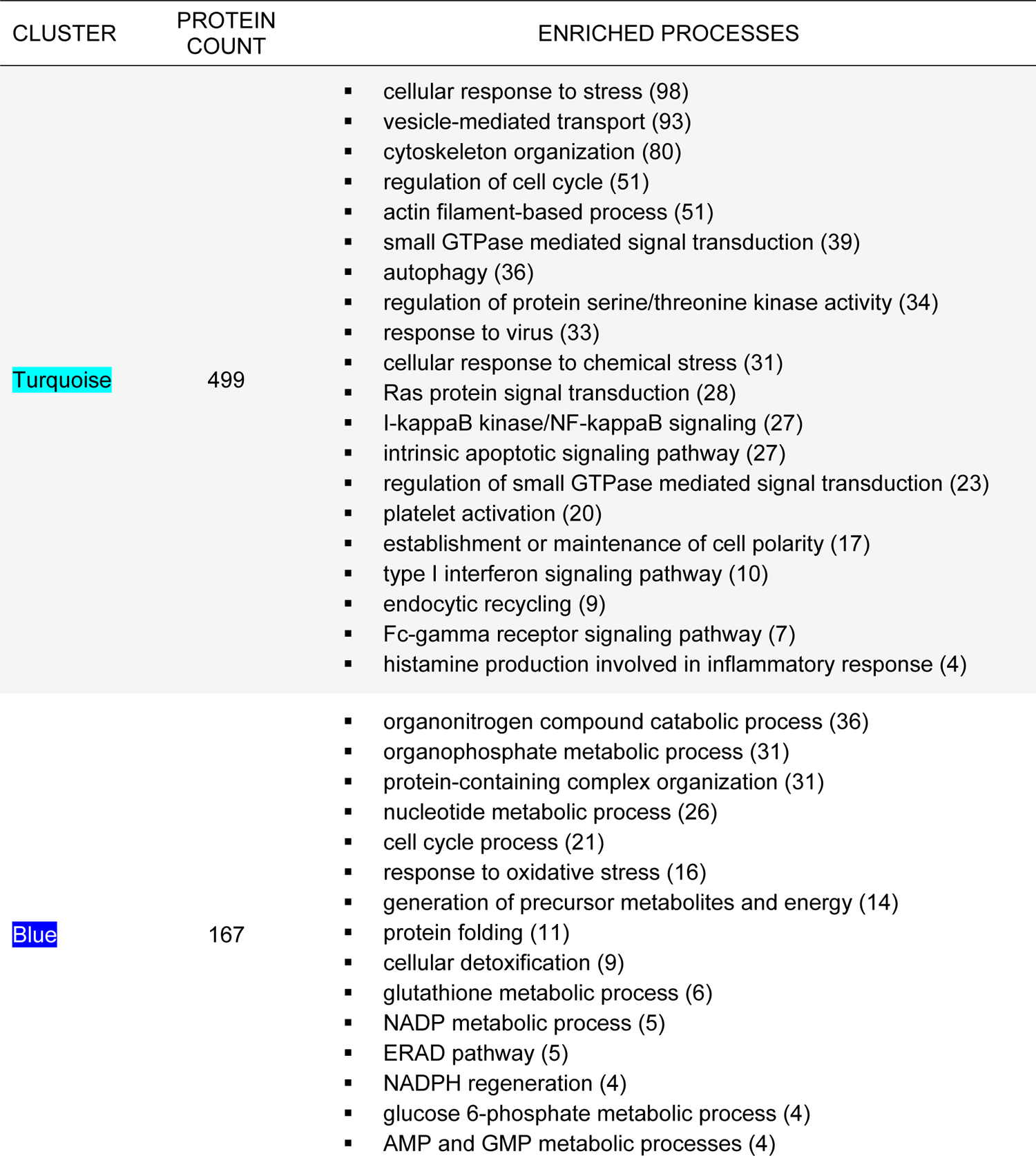
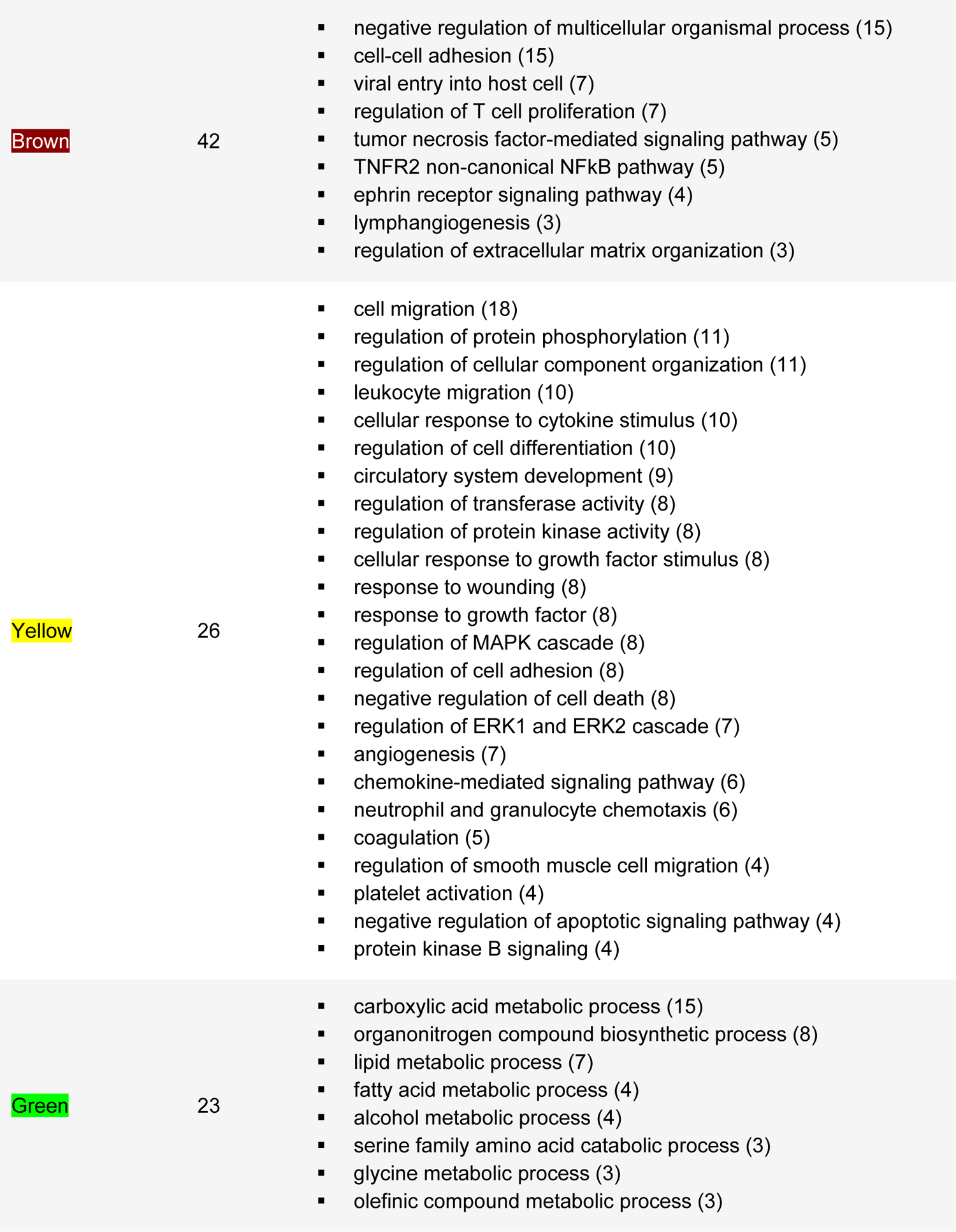
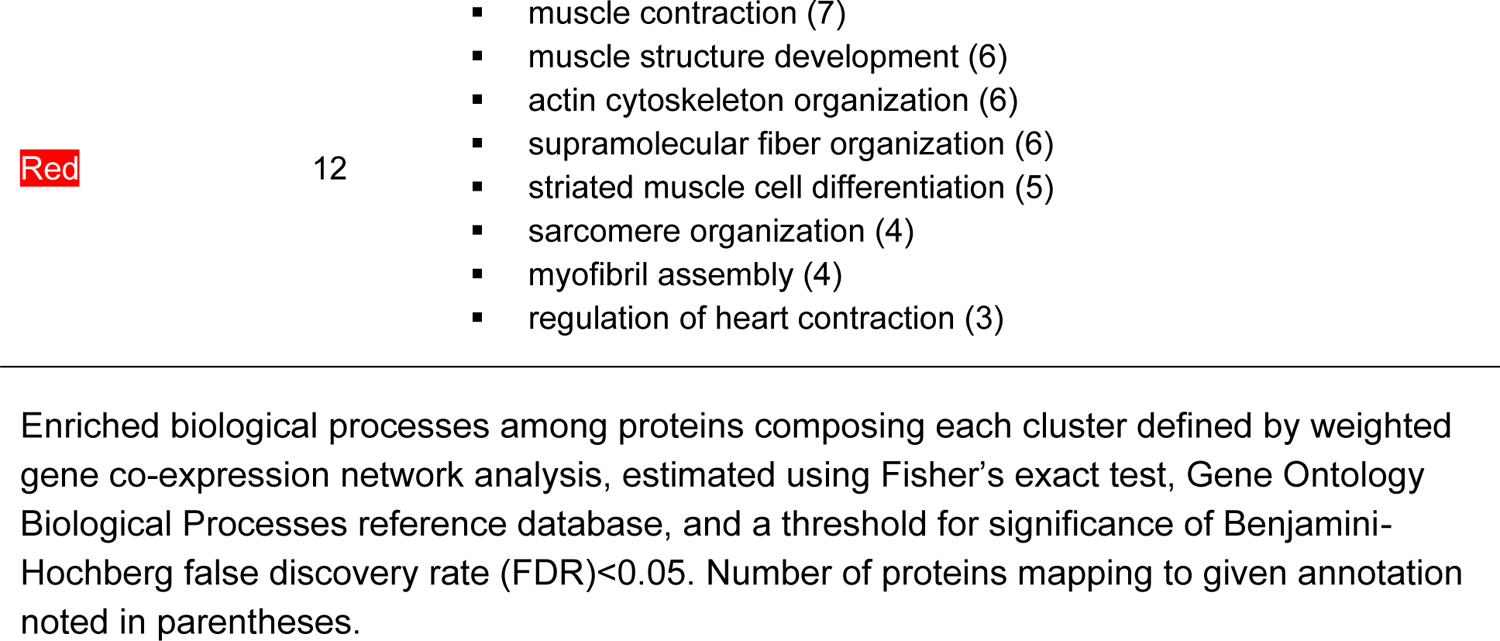
Enriched biological processes and pathways within protein clusters agnostically defined using weighted gene co-expression network analysis.

**SUPPLEMENTAL TABLE S7A.**
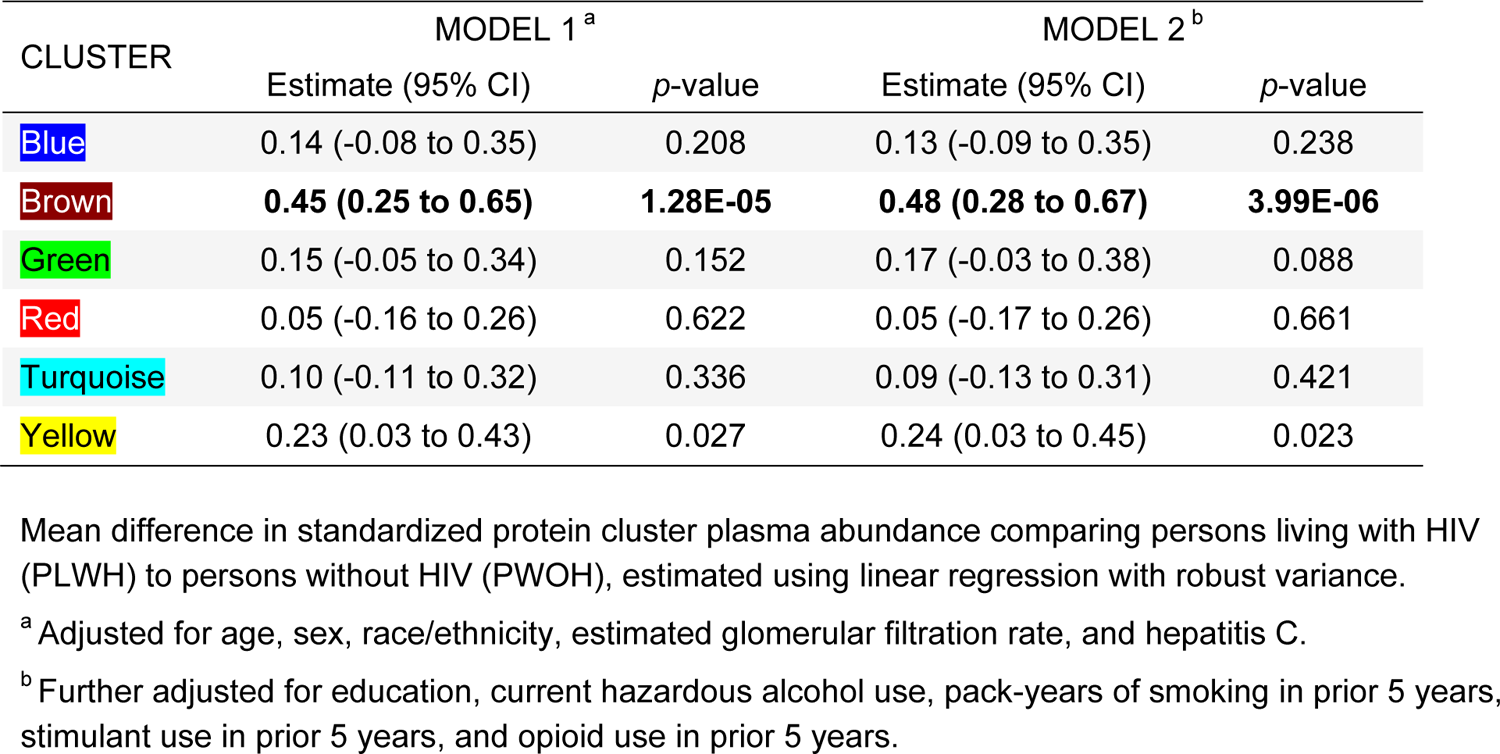
Cross-sectional associations between agnostically defined clusters of proteins and <u>HIV serostatus</u> among PLWH and PWOH in the United States (*n*=352).

**SUPPLEMENTAL TABLE S7B.**
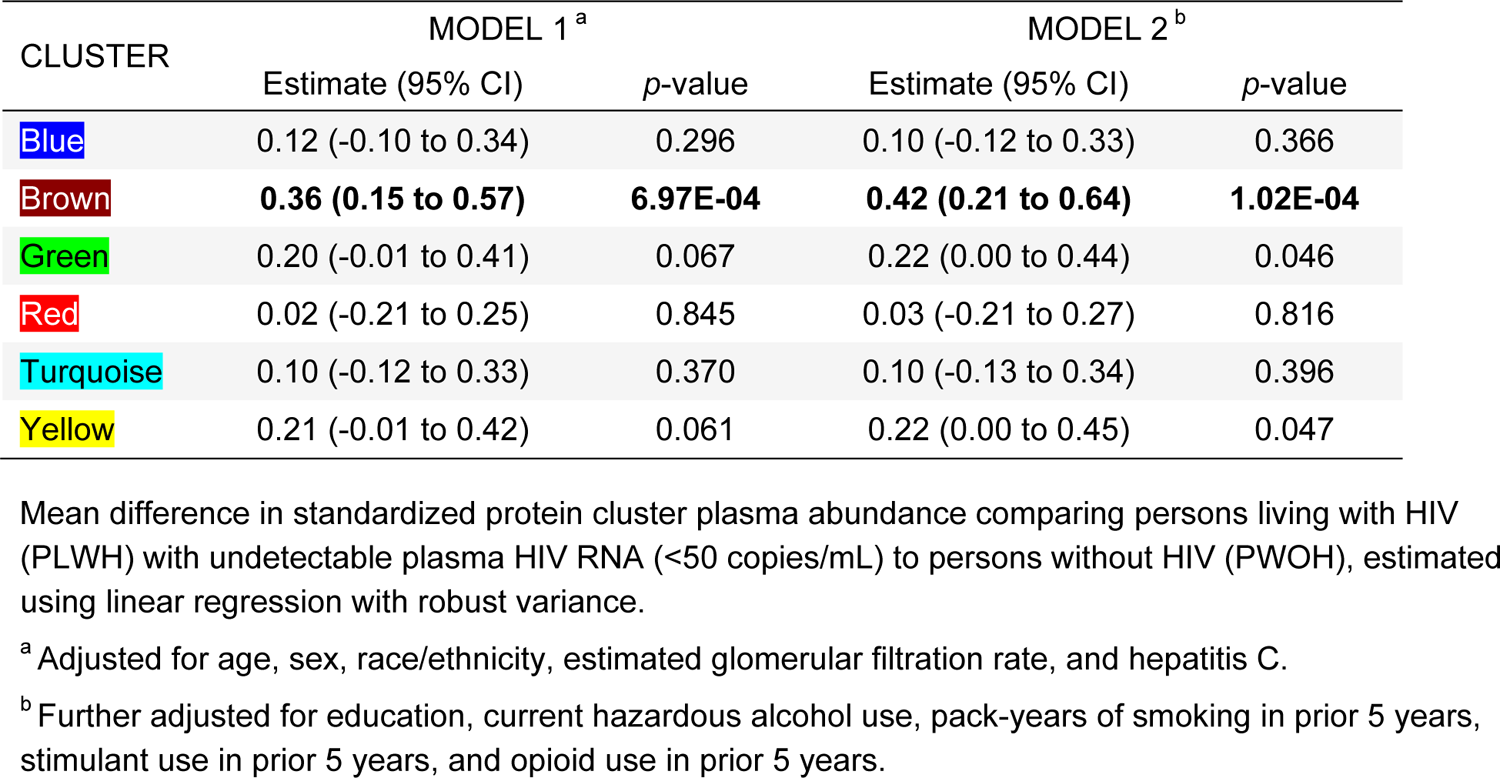
Cross-sectional associations between agnostically defined clusters of proteins and <u>HIV serostatus</u> among PLWH with <u>undetectable plasma HIV RNA</u> and PWOH in the United States (*n*=297).

**SUPPLEMENTAL TABLE S8.** Proteins in Brown cluster defined using weighted gene co-expression network analysis corresponding to statistically over-represented biological processes.

| Gene Ontology:<br>Biological Process | Enrichment<br>FDR | Protein<br>Count | Proteins |
| --- | --- | --- | --- |
| Cell-cell adhesion | 0.022 | 15 | B2M, BSG, CD46, CD93, DSC2, ESAM, FSTL3, HAVCR2, JAM2, NECTIN2, NECTIN4, TGFBR2, TNFRSF14, TNFRSF21, VSIG4 |
| Cytokine production | 0.044 | 13 | ACVRL1, IFNGR1, IL10RB, LTBR, RELT, TGFBR2, TNFRSF10B, TNFRSF11A, TNFRSF14, TNFRSF19, TNFRSF1A, TNFRSF1B, TNFRSF21 |
| Viral entry into host cell | 0.022 | 7 | BSG, CD46, EPHA2, NECTIN2, NECTIN4, SCARB2, TNFRSF14 |
| T cell proliferation | 0.030 | 7 | CD46, HAVCR2, TGFBR2, TNFRSF14, TNFRSF1B, TNFRSF21, VSIG4 |
| TNF signaling | 0.021 | 5 | TNFRSF11A, TNFRSF14, TNFRSF19, TNFRSF1A, TNFRSF1B |
| TNFR2 non-canonical<br>NFkB pathway | 0.024 | 5 | LTBR, TNFRSF11A, TNFRSF14, TNFRSF1A, TNFRSF1B |
| Ephrin signaling | 0.022 | 4 | EFNA4, EPHA2, EPHB4, EPHB6 |
| Lymphangiogenesis | 0.022 | 3 | ACVRL1, CLEC14A, EPHA2 |
| Extracellular matrix<br>organization | 0.022 | 3 | CST3, TNFRSF1A, TNFRSF1B |

<sup>a</sup> Estimated using Fisher's exact test, Gene Ontology Biological Processes reference database, and a threshold for significance of Benjamini-Hochberg false discovery rate (FDR)<0.05.
TNF=tumor necrosis factor.

**SUPPLEMENTAL TABLE S9.** Cross-sectional associations between HIV-associated plasma protein abundances and <u>indexed left atrial volume</u> among PLWH and PWOH in the United States (*n*=352).

See Supplemental Excel file ‘Supplemental Table S9.xlsx’

Mean differences in left atrial volume indexed for body surface area (LAVi) per standard deviation (SD) increment in individual plasma protein abundances, estimated using linear regression with robust variance. Model 1 adjusts for age, sex, and race/ethnicity; Model 2 further adjusts for HIV serostatus, hepatitis C infection, and estimated glomerular filtration rate; and Model 3 further adjusts for education, body mass index, systolic blood pressure, anti-hypertensive medication, dyslipidemia, diabetes, current hazardous alcohol use, pack-years of smoking in prior 5 years, stimulant use in prior 5 years, and opioid use in prior 5 years.

**SUPPLEMENTAL TABLE S10.** Proteins cross-sectionally associated with HIV serostatus and left atrial size corresponding to statistically over-represented biological processes.

| Gene Ontology:<br>Biological Process | Enrichment<br>FDR <sup>a</sup> | Protein<br>Count | Proteins |
| --- | --- | --- | --- |
| Cell-cell adhesion | 1.20E-04 | 26 | CD160, CD27, CD74, CD80, CD83, CDH1, CDH17, CRTAM, CX3CL1, ESAM, HAVCR2, IGFBP2, IL1RL2, ITGB7, JAM2, KLRK1, LAG3, PDGFRA, PODXL2, SCARF2, SIGLEC1, THBS4, TIGIT, TNFRSF14, TNFRSF21, VCAM1 |
| T cell activation | 2.60E-05 | 20 | CD160, CD27, CD48, CD74, CD80, CD83, CRTAM, HAVCR2, IGFBP2, IL1RL2, KLRK1, LAG3, SLAMF7, TIGIT, TNFRSF14, TNFRSF1B, TNFRSF21, TNFRSF4, TNFRSF9, VCAM1 |
| Cytokine production | 0.002 | 20 | AXL, BTN2A1, CD160, CD74, CD80, CD83, COL3A1, CRTAM, CX3CL1, DLL1, EPHA2, HAVCR2, IL1RL2, KLRK1, LAG3, TIGIT, TNFRSF14, TNFRSF1B, TNFRSF21, TNFRSF8 |
| Leukocyte cell-cell adhesion | 7.26E-05 | 18 | CD160, CD27, CD74, CD80, CD83, CRTAM, HAVCR2, IGFBP2, IL1RL2, ITGB7, JAM2, KLRK1, LAG3, PODXL2, TIGIT, TNFRSF14, TNFRSF21, VCAM1 |
| T cell proliferation | 0.007 | 10 | CD80, CRTAM, HAVCR2, IGFBP2, TNFRSF14, TNFRSF1B, TNFRSF21, TNFRSF4, TNFRSF9, VCAM1 |
| TNFR2 non-canonical NFκB pathway | 0.003 | 8 | CD27, TNFRSF1B, TNFRSF4, TNFRSF8, TNFRSF9, TNFRSF13B, TNFRSF14, TNFRSF17 |
| B cell activation | 0.019 | 9 | CD27, CD74, CD79B, CDH17, DLL1, TNFRSF13B, TNFRSF21, TNFRSF4, VCAM1 |
| Cellular response to tumor necrosis factor | 0.022 | 9 | CX3CL1, FAS, IL18BP, TNFRSF1B, TNFRSF4, TNFRSF21, TNFRSF14, TNFRSF17, VCAM1 |
| Viral entry into host cell | 0.029 | 8 | AXL, CD74, CD80, EPHA2, ITGB7, SIGLEC1, TNFRSF4, TNFRSF14 |
| T cell differentiation | 0.045 | 8 | CD27, CD74, CD80, CD83, CRTAM, IL1RL2, LAG3, TNFRSF9 |
| Lymphocyte migration | 0.023 | 7 | CRTAM, CX3CL1, JAM2, CXCL10, TNFRSF14, ITGB7, KLRK1 |
| Natural killer cell mediated cytotoxicity | 0.009 | 6 | CD160, CRTAM, HAVCR2, KLRK1, LAG3, SLAMF7 |
| Platelet derived growth factor signaling | 0.027 | 5 | COL3A1, COL4A1, PDGFRA, THBS2, THBS4 |

<sup>a</sup> Estimated using Fisher's exact test, Gene Ontology Biological Processes reference database, and a threshold for significance of Benjamini-Hochberg false discovery rate (FDR)<0.05.

**SUPPLEMENTAL TABLE S11.**
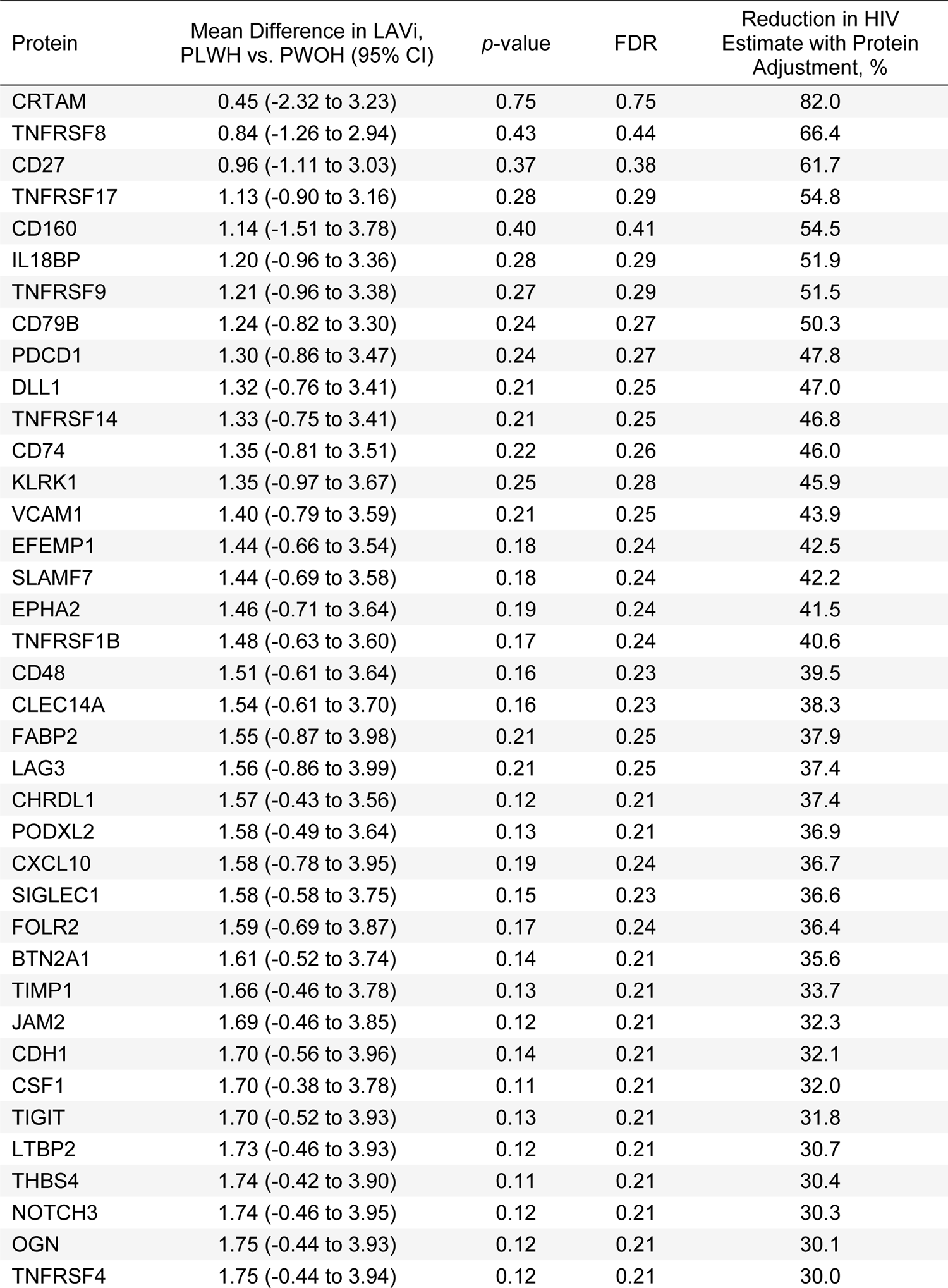
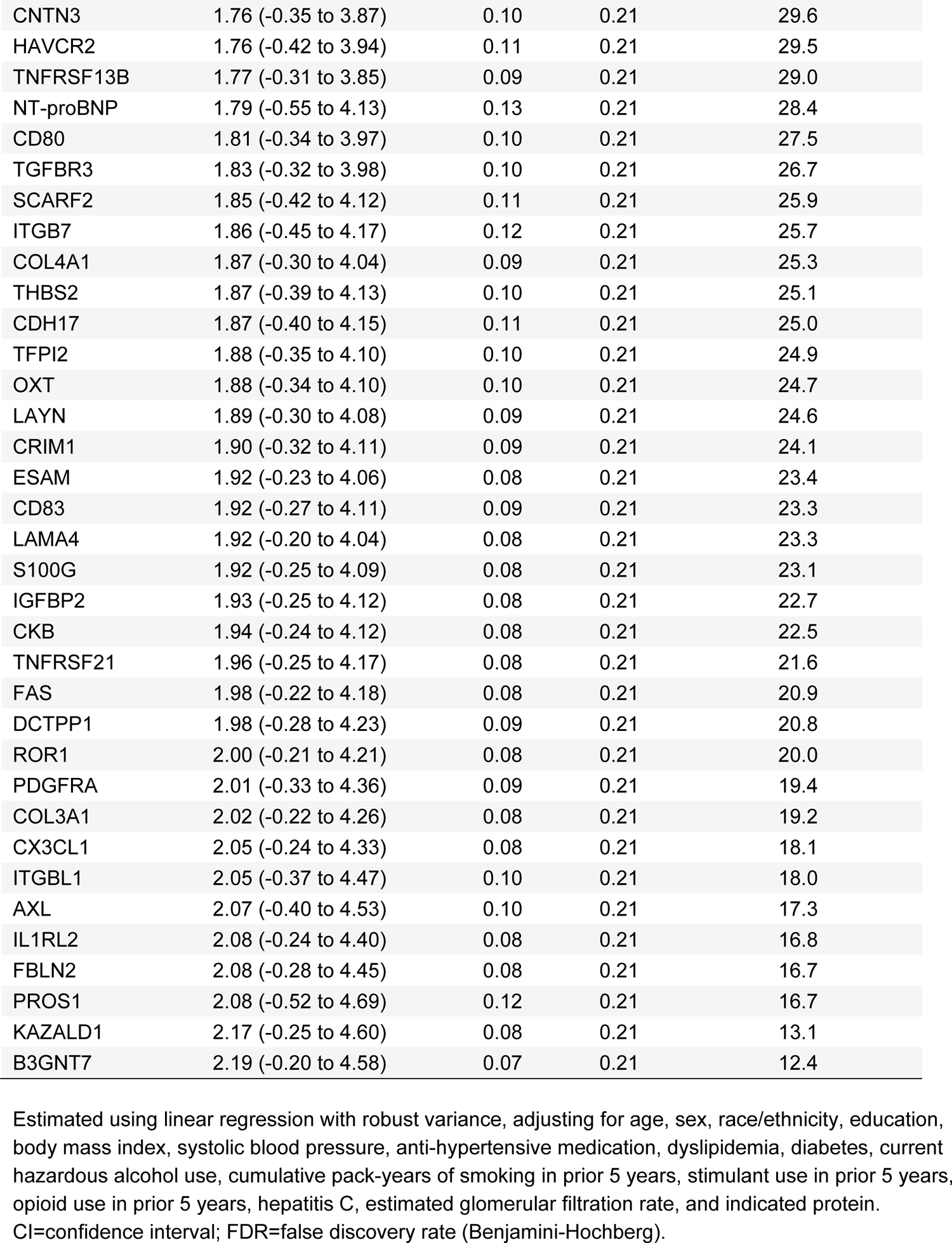
Percent difference in association between HIV serostatus and indexed left atrial volume (LAVi) with adjustment for plasma abundance of 73 individual candidate protein contributors (*n*=352).

**SUPPLEMENTAL TABLE S12.**
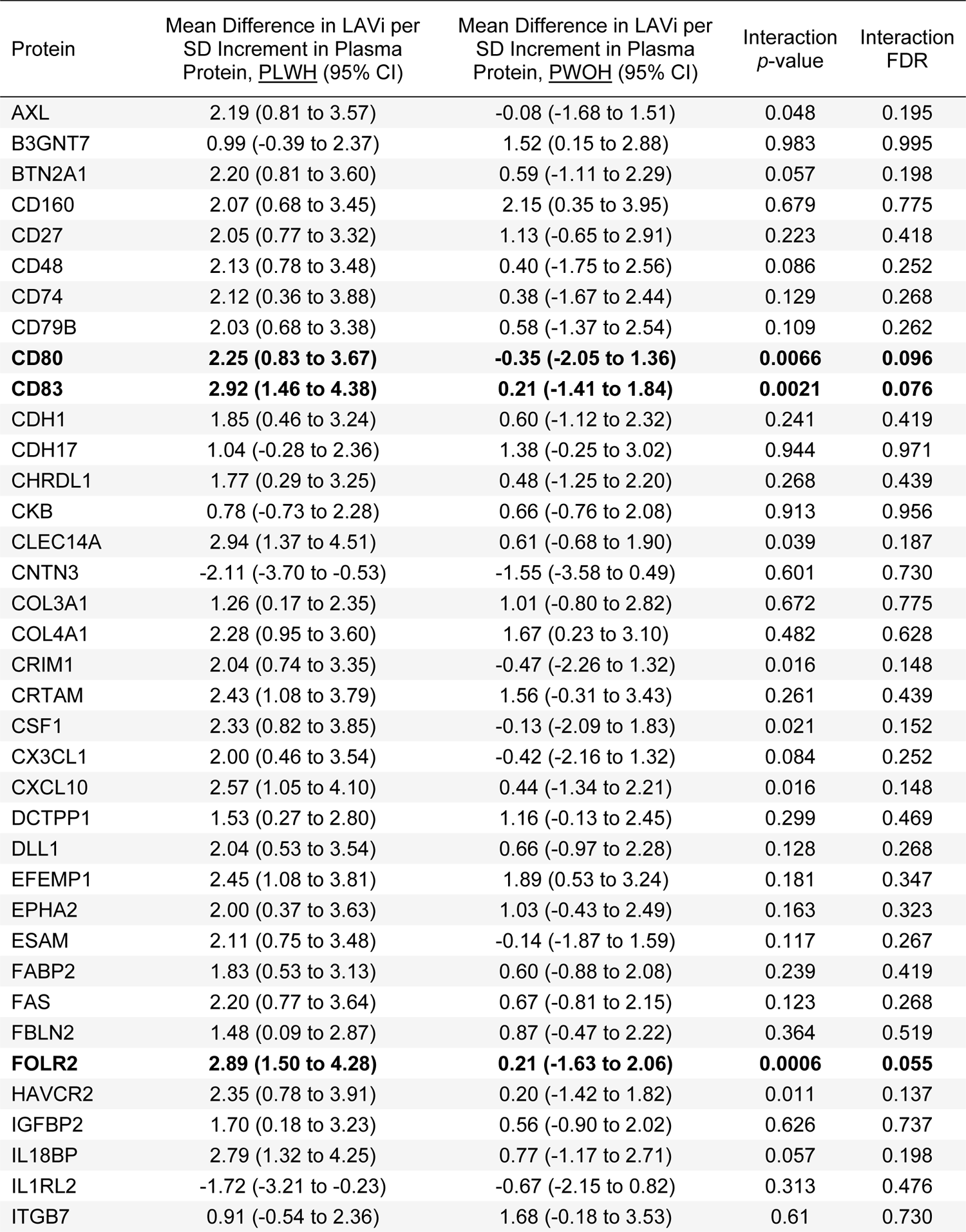
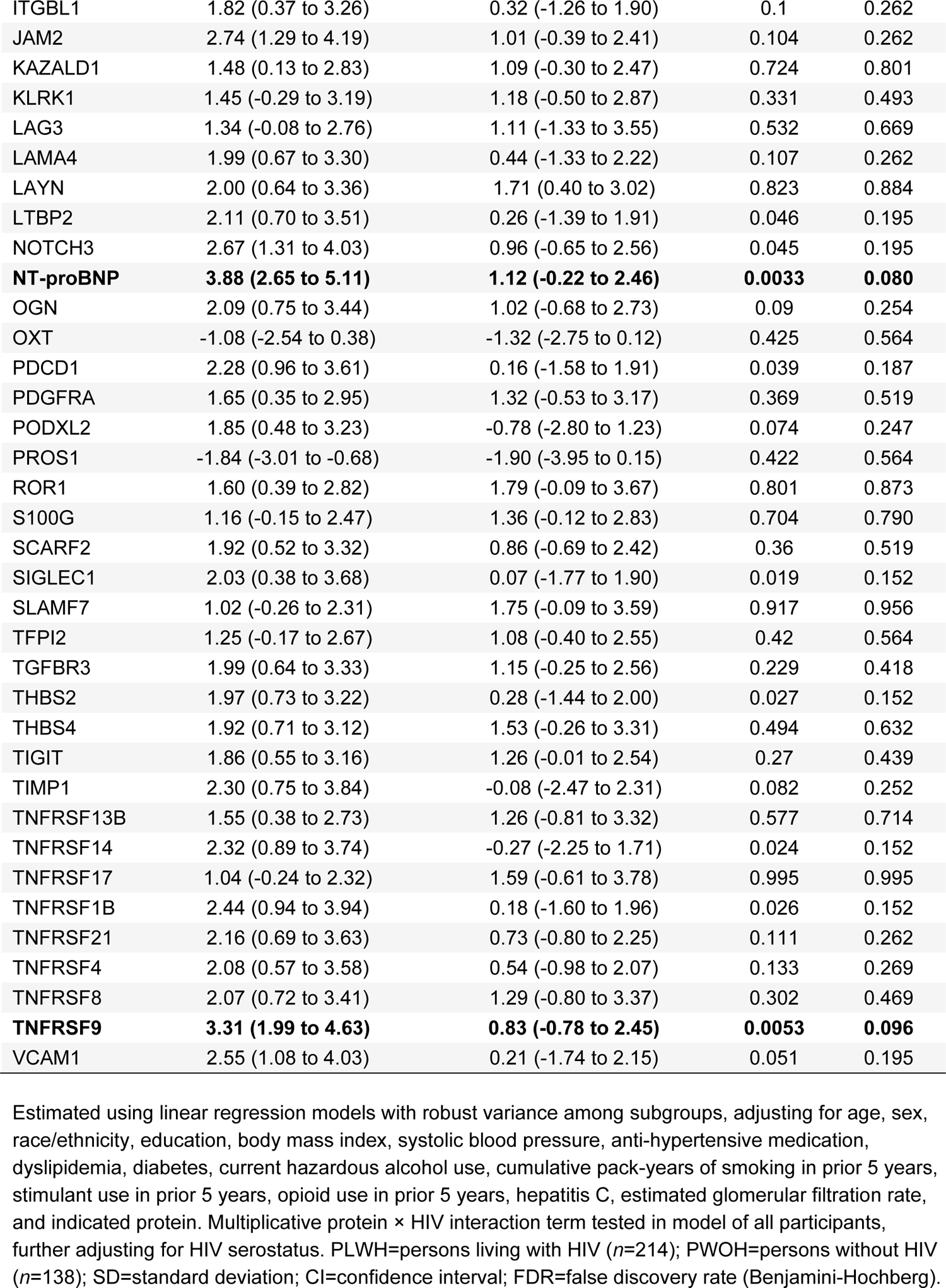
Difference in association between plasma abundance of 73 individual proteins of interest and indexed left atrial volume (LAVi) by HIV serostatus in SMASH (*n*=352).

**SUPPLEMENTAL TABLE S13.** Difference in association between plasma abundance of 73 individual proteins of interest and indexed left atrial volume (LAVi) by age group in SMASH (*n*=352).

| Protein | Mean Difference in LAVi per<br>SD Increment in Plasma<br>Protein, Age $\geq 55$ (95% CI) | Mean Difference in LAVi per<br>SD Increment in Plasma<br>Protein, Age $< 55$ (95% CI) | Interaction<br>$p$-value | Interaction<br>FDR |
| --- | --- | --- | --- | --- |
| AXL | 1.71 (0.17 to 3.25) | 1.07 (-0.26 to 2.40) | 0.703 | 0.994 |
| B3GNT7 | 2.16 (0.84 to 3.49) | 0.42 (-0.80 to 1.64) | 0.277 | 0.994 |
| BTN2A1 | 1.73 (0.26 to 3.20) | 1.26 (-0.26 to 2.78) | 0.935 | 0.994 |
| CD160 | 2.70 (1.04 to 4.36) | 1.62 (0.07 to 3.17) | 0.756 | 0.994 |
| CD27 | 1.89 (0.38 to 3.39) | 1.50 (0.02 to 2.99) | 0.559 | 0.994 |
| CD48 | 1.28 (-0.32 to 2.88) | 2.08 (0.55 to 3.62) | 0.748 | 0.994 |
| CD74 | 0.92 (-0.89 to 2.73) | 2.24 (0.49 to 3.98) | 0.373 | 0.994 |
| CD79B | 1.65 (0.08 to 3.21) | 1.31 (-0.21 to 2.84) | 0.225 | 0.994 |
| CD80 | 1.77 (0.20 to 3.35) | 0.94 (-0.63 to 2.51) | 0.527 | 0.994 |
| CD83 | 1.55 (-0.15 to 3.25) | 1.65 (-0.08 to 3.38) | 0.994 | 0.994 |
| CDH1 | 1.54 (0.18 to 2.90) | 1.27 (-0.31 to 2.85) | 0.536 | 0.994 |
| CDH17 | 1.43 (0.03 to 2.83) | 1.06 (-0.43 to 2.54) | 0.643 | 0.994 |
| CHRD1 | 1.45 (-0.17 to 3.06) | 0.98 (-0.62 to 2.58) | 0.917 | 0.994 |
| CKB | 0.32 (-1.08 to 1.73) | 1.59 (-0.06 to 3.24) | 0.504 | 0.994 |
| CLEC14A | 2.28 (0.89 to 3.67) | 1.37 (-0.19 to 2.93) | 0.914 | 0.994 |
| CNTN3 | -2.45 (-4.24 to -0.67) | -0.91 (-2.54 to 0.72) | 0.304 | 0.994 |
| COL3A1 | 0.90 (-0.57 to 2.37) | 1.48 (0.29 to 2.67) | 0.210 | 0.994 |
| COL4A1 | 2.83 (1.42 to 4.25) | 0.83 (-0.58 to 2.24) | 0.053 | 0.994 |
| CRIM1 | 1.77 (0.35 to 3.19) | 0.77 (-0.75 to 2.28) | 0.249 | 0.994 |
| CRTAM | 2.42 (0.77 to 4.08) | 2.45 (0.82 to 4.09) | 0.627 | 0.994 |
| CSF1 | 1.45 (-0.40 to 3.31) | 1.27 (-0.19 to 2.73) | 0.663 | 0.994 |
| CX3CL1 | 1.18 (-0.36 to 2.72) | 1.33 (-0.49 to 3.16) | 0.771 | 0.994 |
| CXCL10 | 0.80 (-0.97 to 2.56) | 3.15 (1.50 to 4.79) | 0.494 | 0.994 |
| DCTPP1 | 1.29 (-0.13 to 2.72) | 1.35 (0.06 to 2.64) | 0.203 | 0.994 |
| DLL1 | 2.03 (0.45 to 3.60) | 0.94 (-0.59 to 2.47) | 0.530 | 0.994 |
| EFEMP1 | 2.49 (1.14 to 3.84) | 1.72 (0.32 to 3.12) | 0.260 | 0.994 |
| EPHA2 | 1.73 (0.12 to 3.33) | 1.15 (-0.38 to 2.68) | 0.943 | 0.994 |
| ESAM | 1.79 (0.23 to 3.34) | 0.51 (-0.88 to 1.90) | 0.730 | 0.994 |
| FABP2 | 1.79 (0.23 to 3.35) | 1.01 (-0.45 to 2.46) | 0.716 | 0.994 |
| FAS | 1.27 (-0.27 to 2.81) | 1.18 (-0.18 to 2.54) | 0.925 | 0.994 |
| FBLN2 | 1.79 (0.19 to 3.40) | 0.55 (-0.73 to 1.83) | 0.691 | 0.994 |
| FOLR2 | 2.11 (0.62 to 3.61) | 1.75 (0.15 to 3.36) | 0.409 | 0.994 |
| HAVCR2 | 1.78 (0.27 to 3.30) | 0.94 (-0.76 to 2.64) | 0.979 | 0.994 |
| IGFBP2 | 1.27 (-0.25 to 2.78) | 1.66 (0.09 to 3.22) | 0.981 | 0.994 |
| IL18BP | 1.81 (0.22 to 3.40) | 2.01 (0.34 to 3.68) | 0.853 | 0.994 |
| IL1RL2 | 0.17 (-1.54 to 1.88) | -2.44 (-3.70 to -1.18) | 0.105 | 0.994 |
| ITGB7 | 0.90 (-0.98 to 2.78) | 1.28 (-0.23 to 2.78) | 0.431 | 0.994 |
| ITGBL1 | 2.48 (0.76 to 4.21) | 0.33 (-0.97 to 1.64) | 0.985 | 0.994 |
| JAM2 | 2.41 (0.84 to 3.97) | 1.39 (0.09 to 2.69) | 0.797 | 0.994 |
| KAZALD1 | 2.34 (0.88 to 3.80) | 0.96 (-0.29 to 2.20) | 0.294 | 0.994 |
| KLRK1 | 1.09 (-0.61 to 2.79) | 1.28 (-0.30 to 2.85) | 0.601 | 0.994 |
| LAG3 | 1.47 (-0.38 to 3.32) | 1.78 (0.24 to 3.32) | 0.708 | 0.994 |
| LAMA4 | 1.66 (0.12 to 3.21) | 0.55 (-1.03 to 2.14) | 0.351 | 0.994 |
| LAYN | 2.22 (1.05 to 3.40) | 1.25 (-0.23 to 2.72) | 0.994 | 0.994 |
| LTBP2 | 2.67 (1.14 to 4.19) | -0.07 (-1.47 to 1.33) | 0.202 | 0.994 |
| NOTCH3 | 2.12 (0.73 to 3.51) | 2.18 (0.66 to 3.70) | 0.205 | 0.994 |
| NT-proBNP | 2.79 (1.51 to 4.07) | 3.34 (1.91 to 4.77) | 0.569 | 0.994 |
| OGN | 1.63 (0.18 to 3.07) | 1.51 (-0.13 to 3.15) | 0.537 | 0.994 |
| OXT | -0.89 (-2.46 to 0.68) | -1.52 (-2.81 to -0.22) | 0.226 | 0.994 |
| PDCD1 | 1.73 (0.11 to 3.35) | 1.63 (0.10 to 3.15) | 0.565 | 0.994 |
| PDGFRA | 2.25 (0.80 to 3.71) | 0.83 (-0.65 to 2.31) | 0.238 | 0.994 |
| PODXL2 | 0.93 (-0.71 to 2.57) | 1.46 (-0.12 to 3.04) | 0.811 | 0.994 |
| PROS1 | -1.25 (-2.90 to 0.40) | -2.43 (-3.66 to -1.20) | 0.781 | 0.994 |
| ROR1 | 1.43 (0.01 to 2.86) | 1.78 (0.34 to 3.21) | 0.591 | 0.994 |
| S100G | 1.85 (0.50 to 3.21) | 0.24 (-1.12 to 1.60) | 0.421 | 0.994 |
| SCARF2 | 2.01 (0.62 to 3.41) | 1.12 (-0.31 to 2.55) | 0.914 | 0.994 |
| SIGLEC1 | 1.08 (-0.59 to 2.75) | 1.67 (-0.04 to 3.38) | 0.992 | 0.994 |
| SLAMF7 | 1.15 (-0.31 to 2.62) | 1.49 (-0.01 to 3.00) | 0.260 | 0.994 |
| TFPI2 | 1.70 (0.36 to 3.05) | 0.63 (-0.99 to 2.25) | 0.359 | 0.994 |
| TGFBR3 | 1.79 (0.56 to 3.03) | 1.40 (-0.22 to 3.01) | 0.863 | 0.994 |
| THBS2 | 2.24 (0.59 to 3.88) | 0.95 (-0.25 to 2.15) | 0.923 | 0.994 |
| THBS4 | 1.92 (0.37 to 3.46) | 2.14 (0.70 to 3.58) | 0.706 | 0.994 |
| TIGIT | 2.14 (0.91 to 3.36) | 0.64 (-0.73 to 2.01) | 0.976 | 0.994 |
| TIMP1 | 2.56 (0.58 to 4.55) | 0.29 (-1.45 to 2.03) | 0.446 | 0.994 |
| TNFRSF13B | 1.33 (-0.31 to 2.97) | 1.51 (-0.05 to 3.07) | 0.709 | 0.994 |
| TNFRSF14 | 1.82 (0.24 to 3.39) | 0.68 (-0.98 to 2.35) | 0.974 | 0.994 |
| TNFRSF17 | 0.86 (-0.99 to 2.71) | 1.43 (0.04 to 2.81) | 0.848 | 0.994 |
| TNFRSF1B | 1.96 (0.27 to 3.65) | 0.78 (-0.94 to 2.51) | 0.524 | 0.994 |
| TNFRSF21 | 1.40 (-0.04 to 2.84) | 1.42 (-0.05 to 2.89) | 0.994 | 0.994 |
| TNFRSF4 | 0.90 (-0.63 to 2.43) | 1.96 (0.49 to 3.44) | 0.199 | 0.994 |
| TNFRSF8 | 1.78 (0.20 to 3.37) | 1.69 (0.15 to 3.24) | 0.794 | 0.994 |
| TNFRSF9 | 2.26 (0.78 to 3.74) | 2.35 (0.82 to 3.88) | 0.824 | 0.994 |
| VCAM1 | 2.58 (0.86 to 4.29) | 1.49 (-0.01 to 2.99) | 0.285 | 0.994 |

Estimated using linear regression models with robust variance among subgroups, adjusting for age, sex, race/ethnicity, education, body mass index, systolic blood pressure, anti-hypertensive medication, dyslipidemia, diabetes, current hazardous alcohol use, cumulative pack-years of smoking in prior 5 years, stimulant use in prior 5 years, opioid use in prior 5 years, HIV, hepatitis C, estimated glomerular filtration rate, and indicated protein. Multiplicative protein × age interaction term tested in model of all participants. Subgroups defined by dichotomization at median age in SMASH (55 years); SD=standard deviation; CI=confidence interval; FDR=false discovery rate (Benjamini-Hochberg).

**SUPPLEMENTAL TABLE S14.** Cross-sectional associations between proteins of interest and indexed left atrial volume (LAVi) in the Multi-Ethnic Study of Atherosclerosis (median age 67 years, *n*=1242)

| Protein | Model 1 |  |  | Model 2 |  |  |
| --- | --- | --- | --- | --- | --- | --- |
|  | Mean Difference in LAVi per<br>SD Increment in Plasma<br>Protein (95% CI) | *p*-value | FDR | Mean Difference in LAVi per<br>SD Increment in Plasma<br>Protein (95% CI) | *p*-value | FDR |
| AXL | 0.71 (0.11 to 1.31) | 0.02 | 0.08 | 0.48 (-0.12 to 1.09) | 0.11 | 0.31 |
| B3GNT7 | 0.54 (-0.06 to 1.14) | 0.08 | 0.19 | 0.59 (-0.01 to 1.19) | 0.06 | 0.19 |
| BTN2A1 | 0.41 (-0.31 to 1.13) | 0.26 | 0.37 | 0.30 (-0.41 to 1.01) | 0.40 | 0.52 |
| CD160 | 0.62 (-0.06 to 1.30) | 0.07 | 0.19 | 0.49 (-0.18 to 1.16) | 0.15 | 0.32 |
| CD27 | 0.36 (-0.31 to 1.03) | 0.29 | 0.39 | 0.32 (-0.35 to 0.98) | 0.35 | 0.52 |
| CD48 | 0.66 (0.05 to 1.27) | 0.04 | 0.12 | 0.46 (-0.15 to 1.08) | 0.14 | 0.32 |
| CD74 | 0.52 (-0.18 to 1.23) | 0.15 | 0.26 | 0.51 (-0.21 to 1.23) | 0.16 | 0.32 |
| CD79B | 0.46 (-0.18 to 1.10) | 0.16 | 0.27 | 0.42 (-0.22 to 1.06) | 0.20 | 0.37 |
| CD80 | 0.29 (-0.30 to 0.87) | 0.34 | 0.44 | 0.19 (-0.39 to 0.77) | 0.52 | 0.61 |
| CD83 | 0.29 (-0.37 to 0.94) | 0.39 | 0.46 | 0.29 (-0.38 to 0.95) | 0.40 | 0.52 |
| CDH1 | -0.49 (-1.07 to 0.10) | 0.10 | 0.21 | -0.49 (-1.09 to 0.10) | 0.10 | 0.29 |
| CDH17 | 0.06 (-0.54 to 0.67) | 0.83 | 0.88 | 0.11 (-0.49 to 0.70) | 0.73 | 0.79 |
| CHRD1 | -0.01 (-0.65 to 0.63) | 0.98 | 0.98 | 0.01 (-0.62 to 0.64) | 0.97 | 0.97 |
| **CKB** | 0.91 (0.24 to 1.58) | 0.008 | 0.06 | **1.15 (0.45 to 1.84)** | **0.001** | **0.02** |
| **CLEC14A** | **1.45 (0.73 to 2.18)** | **8.2E-05** | **0.003** | **1.36 (0.65 to 2.07)** | **1.8E-04** | **0.01** |
| **CNTN3** | -0.79 (-1.43 to -0.16) | 0.01 | 0.07 | **-1.05 (-1.73 to -0.38)** | **0.002** | **0.03** |
| COL3A1 | 0.77 (0.13 to 1.40) | 0.02 | 0.07 | 0.71 (0.09 to 1.34) | 0.03 | 0.12 |
| **COL4A1** | **1.21 (0.57 to 1.86)** | **2.3E-04** | **0.006** | **1.20 (0.56 to 1.85)** | **2.5E-04** | **0.01** |
| CRIM1 | 0.91 (0.25 to 1.57) | 0.007 | 0.06 | 0.83 (0.19 to 1.48) | 0.01 | 0.09 |
| CRTAM | 0.6 (-0.04 to 1.24) | 0.07 | 0.18 | 0.51 (-0.13 to 1.15) | 0.12 | 0.31 |
| CSF1 | 0.6 (-0.07 to 1.27) | 0.08 | 0.19 | 0.39 (-0.29 to 1.06) | 0.26 | 0.42 |
| CX3CL1 | 0.88 (0.25 to 1.50) | 0.006 | 0.06 | 0.82 (0.20 to 1.44) | 0.01 | 0.08 |
| CXCL10 | 0.52 (-0.09 to 1.13) | 0.10 | 0.21 | 0.35 (-0.28 to 0.98) | 0.27 | 0.43 |
| DCTPP1 | 0.73 (0.11 to 1.36) | 0.02 | 0.08 | 0.64 (0.02 to 1.27) | 0.04 | 0.17 |
| DLL1 | 0.31 (-0.40 to 1.03) | 0.39 | 0.46 | 0.21 (-0.51 to 0.92) | 0.57 | 0.66 |
| EFEMP1 | 0.32 (-0.36 to 0.99) | 0.36 | 0.45 | 0.26 (-0.41 to 0.92) | 0.45 | 0.56 |
| EPHA2 | 0.85 (0.16 to 1.54) | 0.02 | 0.07 | 0.76 (0.08 to 1.45) | 0.03 | 0.13 |
| ESAM | 0.69 (-0.02 to 1.39) | 0.06 | 0.16 | 0.54 (-0.15 to 1.23) | 0.13 | 0.31 |
| FABP2 | -0.06 (-0.71 to 0.58) | 0.84 | 0.88 | 0.10 (-0.54 to 0.73) | 0.77 | 0.83 |
| FAS | -0.01 (-0.62 to 0.60) | 0.97 | 0.98 | -0.07 (-0.67 to 0.53) | 0.82 | 0.86 |
| FBLN2 | 0.42 (-0.22 to 1.06) | 0.20 | 0.29 | 0.28 (-0.35 to 0.91) | 0.39 | 0.52 |
| FOLR2 | 0.55 (-0.10 to 1.21) | 0.10 | 0.21 | 0.39 (-0.26 to 1.05) | 0.24 | 0.40 |
| HAVCR2 | 0.48 (-0.24 to 1.20) | 0.19 | 0.29 | 0.34 (-0.39 to 1.06) | 0.36 | 0.52 |
| **IGFBP2** | **0.99 (0.33 to 1.65)** | **0.003** | **0.04** | **1.26 (0.56 to 1.97)** | **4.4E-04** | **0.008** |
| IL18BP | 0.46 (-0.22 to 1.13) | 0.18 | 0.29 | 0.35 (-0.32 to 1.02) | 0.31 | 0.47 |
| IL1RL2 | -0.16 (-0.78 to 0.46) | 0.62 | 0.67 | -0.14 (-0.75 to 0.48) | 0.66 | 0.74 |
| ITGB7 | 0.40 (-0.20 to 1.00) | 0.19 | 0.29 | 0.35 (-0.24 to 0.94) | 0.24 | 0.40 |
| ITGBL1 | 0.56 (-0.10 to 1.22) | 0.09 | 0.21 | 0.49 (-0.16 to 1.15) | 0.14 | 0.32 |
| JAM2 | 0.60 (-0.13 to 1.33) | 0.11 | 0.21 | 0.52 (-0.20 to 1.24) | 0.16 | 0.32 |
| KAZALD1 | 0.66 (0.03 to 1.29) | 0.04 | 0.13 | 0.53 (-0.09 to 1.16) | 0.09 | 0.29 |
| KLRK1 | 0.44 (-0.15 to 1.03) | 0.15 | 0.26 | 0.38 (-0.20 to 0.96) | 0.20 | 0.37 |
| LAG3 | 0.04 (-0.60 to 0.68) | 0.90 | 0.93 | 0.01 (-0.64 to 0.66) | 0.97 | 0.97 |
| LAMA4 | 0.88 (0.20 to 1.55) | 0.01 | 0.06 | 0.79 (0.13 to 1.45) | 0.02 | 0.11 |
| LAYN | 0.56 (-0.18 to 1.29) | 0.14 | 0.25 | 0.56 (-0.16 to 1.29) | 0.13 | 0.31 |
| LTBP2 | 0.76 (0.10 to 1.43) | 0.03 | 0.09 | 0.70 (0.04 to 1.36) | 0.04 | 0.16 |
| **NOTCH3** | **1.05 (0.39 to 1.72)** | **0.002** | **0.03** | **0.95 (0.30 to 1.61)** | **0.004** | **0.04** |
| **NT-proBNP** | **3.63 (2.93 to 4.33)** | **<1.0E-20** | **<1.0E-20** | **3.41 (2.70 to 4.11)** | **<1.0E-20** | **<1.0E-20** |
| OGN | 0.26 (-0.54 to 1.07) | 0.52 | 0.58 | 0.17 (-0.63 to 0.98) | 0.67 | 0.74 |
| OXT | -0.44 (-1.03 to 0.14) | 0.14 | 0.25 | -0.60 (-1.20 to 0.00) | 0.05 | 0.18 |
| PDCD1 | 0.42 (-0.21 to 1.04) | 0.19 | 0.29 | 0.44 (-0.17 to 1.06) | 0.16 | 0.32 |
| **PDGFRA** | **1.07 (0.43 to 1.71)** | **0.001** | **0.02** | **0.94 (0.32 to 1.57)** | **0.003** | **0.03** |
| PODXL2 | 0.37 (-0.29 to 1.02) | 0.27 | 0.38 | 0.30 (-0.36 to 0.96) | 0.38 | 0.52 |
| PROS1 | -0.78 (-1.40 to -0.16) | 0.01 | 0.07 | -0.72 (-1.33 to -0.10) | 0.02 | 0.12 |
| ROR1 | 0.77 (0.08 to 1.46) | 0.03 | 0.10 | 0.65 (-0.03 to 1.34) | 0.06 | 0.20 |
| S100G | -0.29 (-0.94 to 0.36) | 0.38 | 0.46 | -0.24 (-0.87 to 0.40) | 0.46 | 0.56 |
| SCARF2 | 0.95 (0.23 to 1.67) | 0.01 | 0.06 | 0.88 (0.18 to 1.59) | 0.01 | 0.09 |
| SIGLEC1 | 0.21 (-0.40 to 0.83) | 0.50 | 0.56 | -0.05 (-0.69 to 0.58) | 0.87 | 0.90 |
| SLAMF7 | 0.28 (-0.34 to 0.90) | 0.38 | 0.46 | 0.14 (-0.46 to 0.74) | 0.66 | 0.74 |
| TFPI2 | 0.56 (-0.12 to 1.24) | 0.11 | 0.21 | 0.45 (-0.23 to 1.13) | 0.20 | 0.37 |
| TGFBR3 | 0.53 (-0.06 to 1.12) | 0.08 | 0.19 | 0.49 (-0.10 to 1.07) | 0.10 | 0.29 |
| THBS2 | 0.63 (-0.08 to 1.34) | 0.08 | 0.19 | 0.46 (-0.26 to 1.18) | 0.21 | 0.37 |
| **THBS4** | **0.95 (0.33 to 1.58)** | **0.003** | **0.03** | 0.76 (0.11 to 1.42) | 0.02 | 0.12 |
| TIGIT | -0.07 (-0.71 to 0.57) | 0.82 | 0.88 | -0.08 (-0.71 to 0.55) | 0.80 | 0.85 |
| TIMP1 | 0.84 (0.15 to 1.52) | 0.02 | 0.07 | 0.70 (0.01 to 1.38) | 0.05 | 0.17 |
| TNFRSF13B | 0.32 (-0.31 to 0.94) | 0.32 | 0.43 | 0.36 (-0.27 to 0.99) | 0.27 | 0.42 |
| TNFRSF14 | 0.39 (-0.32 to 1.10) | 0.28 | 0.39 | 0.25 (-0.45 to 0.96) | 0.48 | 0.58 |
| TNFRSF17 | 0.27 (-0.38 to 0.92) | 0.41 | 0.48 | 0.28 (-0.37 to 0.93) | 0.40 | 0.52 |
| TNFRSF1B | 0.45 (-0.26 to 1.16) | 0.21 | 0.31 | 0.32 (-0.39 to 1.03) | 0.38 | 0.52 |
| TNFRSF21 | 0.33 (-0.35 to 1.01) | 0.34 | 0.44 | 0.25 (-0.42 to 0.92) | 0.46 | 0.56 |
| TNFRSF4 | 0.48 (-0.23 to 1.19) | 0.18 | 0.29 | 0.44 (-0.26 to 1.13) | 0.22 | 0.38 |
| TNFRSF8 | 0.27 (-0.39 to 0.94) | 0.42 | 0.48 | 0.30 (-0.37 to 0.97) | 0.38 | 0.52 |
| TNFRSF9 | 0.67 (-0.01 to 1.34) | 0.05 | 0.16 | 0.71 (0.04 to 1.39) | 0.04 | 0.16 |
| VCAM1 | 0.62 (-0.05 to 1.28) | 0.07 | 0.19 | 0.49 (-0.17 to 1.15) | 0.14 | 0.32 |

Estimated using linear regression with robust variance. Model 1 adjusts for study site, age, sex, race/ethnicity, and estimated glomerular filtration rate. Model 2 further adjusts for education, body mass index, systolic blood pressure, anti-hypertensive medication, hyperlipidemia, diabetes, and smoking. SD=standard deviation; CI=confidence interval; FDR=false discovery rate (Benjamini-Hochberg).

**SUPPLEMENTAL TABLE S15.** Cross-sectional associations between proteins of interest and indexed left atrial volume (LAVi) by age group in the Multi-Ethnic Study of Atherosclerosis (median age 67 years, *n*=1242)

| Protein | Mean Difference in LAVi per SD Increment in Plasma Protein, <u>Age $\geq 67$</u> (95% CI) | Mean Difference in LAVi per SD Increment in Plasma Protein, <u>Age $< 67$</u> (95% CI) | Interaction $p$-value | Interaction FDR |
| --- | --- | --- | --- | --- |
| AXL | 1.00 (0.05 to 1.95) | 0.01 (-0.72 to 0.75) | 0.07 | 0.12 |
| **B3GNT7** | **1.38 (0.46 to 2.29)** | **-0.16 (-0.92 to 0.61)** | **0.001** | **0.01** |
| **BTN2A1** | **1.03 (-0.01 to 2.07)** | **-0.58 (-1.49 to 0.34)** | **0.001** | **0.01** |
| CD160 | 0.49 (-0.55 to 1.53) | 0.45 (-0.38 to 1.28) | 0.41 | 0.50 |
| **CD27** | **0.69 (-0.32 to 1.69)** | **-0.17 (-1.04 to 0.71)** | **0.02** | **0.05** |
| CD48 | 0.77 (-0.19 to 1.73) | 0.18 (-0.56 to 0.92) | 0.35 | 0.43 |
| **CD74** | **1.10 (-0.04 to 2.25)** | **-0.25 (-1.12 to 0.62)** | **0.006** | **0.03** |
| CD79B | 0.37 (-0.65 to 1.40) | 0.45 (-0.34 to 1.24) | 0.53 | 0.61 |
| CD80 | 0.29 (-0.69 to 1.28) | -0.01 (-0.70 to 0.68) | 0.14 | 0.19 |
| **CD83** | **0.83 (-0.20 to 1.87)** | **-0.30 (-1.11 to 0.51)** | **0.04** | **0.08** |
| CDH1 | -0.26 (-1.23 to 0.70) | -0.56 (-1.29 to 0.17) | 0.24 | 0.30 |
| CDH17 | -0.21 (-1.11 to 0.70) | 0.55 (-0.19 to 1.28) | 0.77 | 0.81 |
| CHRD1 | 0.41 (-0.56 to 1.38) | -0.28 (-1.09 to 0.53) | 0.20 | 0.26 |
| CKB | 1.18 (0.11 to 2.26) | 0.91 (0.05 to 1.77) | 0.17 | 0.22 |
| **CLEC14A** | **2.28 (1.24 to 3.32)** | **0.38 (-0.55 to 1.31)** | **4.3E-04** | **0.01** |
| CNTN3 | -0.58 (-1.60 to 0.44) | -1.43 (-2.33 to -0.52) | 0.09 | 0.14 |
| **COL3A1** | **0.93 (-0.01 to 1.88)** | **0.35 (-0.42 to 1.11)** | **0.01** | **0.03** |
| **COL4A1** | **2.10 (1.12 to 3.08)** | **0.30 (-0.48 to 1.07)** | **0.001** | **0.01** |
| **CRIM1** | **1.47 (0.52 to 2.42)** | **0.16 (-0.66 to 0.99)** | **0.006** | **0.02** |
| CRTAM | 1.06 (0.02 to 2.09) | 0.04 (-0.72 to 0.81) | 0.09 | 0.13 |
| CSF1 | 0.56 (-0.49 to 1.60) | 0.03 (-0.77 to 0.83) | 0.07 | 0.12 |
| **CX3CL1** | **1.62 (0.66 to 2.59)** | **-0.06 (-0.84 to 0.71)** | **0.006** | **0.03** |
| **CXCL10** | **0.67 (-0.33 to 1.67)** | **-0.04 (-0.82 to 0.74)** | **0.01** | **0.04** |
| **DCTPP1** | **1.71 (0.78 to 2.64)** | **-0.25 (-1.04 to 0.53)** | **0.001** | **0.01** |
| **DLL1** | **1.08 (0.05 to 2.10)** | **-0.67 (-1.60 to 0.26)** | **0.005** | **0.02** |
| **EFEMP1** | **0.80 (-0.19 to 1.79)** | **-0.31 (-1.16 to 0.55)** | **0.04** | **0.08** |
| **EPHA2** | **1.42 (0.40 to 2.44)** | **-0.11 (-0.98 to 0.76)** | **0.002** | **0.01** |
| **ESAM** | **0.94 (-0.05 to 1.93)** | **-0.01 (-0.96 to 0.94)** | **0.04** | **0.07** |
| **FABP2** | **0.93 (-0.07 to 1.93)** | **-0.69 (-1.51 to 0.13)** | **0.02** | **0.05** |
| FAS | 0.16 (-0.73 to 1.05) | -0.22 (-0.98 to 0.54) | 0.09 | 0.13 |
| **FBLN2** | **0.65 (-0.36 to 1.66)** | **-0.24 (-0.98 to 0.51)** | **0.007** | **0.03** |
| FOLR2 | 0.49 (-0.44 to 1.41) | 0.16 (-0.76 to 1.08) | 0.44 | 0.52 |
| **HAVCR2** | **0.63 (-0.50 to 1.75)** | **-0.04 (-1.00 to 0.92)** | **0.01** | **0.04** |
| IGFBP2 | 1.40 (0.28 to 2.52) | 0.95 (0.09 to 1.80) | 0.15 | 0.21 |
| **IL18BP** | **1.03 (-0.01 to 2.08)** | **-0.45 (-1.24 to 0.35)** | **0.006** | **0.02** |
| IL1RL2 | -0.23 (-1.18 to 0.73) | 0.08 (-0.70 to 0.87) | 0.42 | 0.51 |
| ITGB7 | 0.83 (-0.03 to 1.70) | -0.07 (-0.85 to 0.71) | 0.10 | 0.15 |
| ITGBL1 | 1.01 (-0.06 to 2.07) | -0.04 (-0.80 to 0.71) | 0.09 | 0.14 |
| **JAM2** | **1.23 (0.22 to 2.25)** | **-0.24 (-1.25 to 0.77)** | **0.03** | **0.06** |
| **KAZALD1** | **1.12 (0.12 to 2.12)** | **-0.08 (-0.83 to 0.68)** | **0.008** | **0.03** |
| KLRK1 | 0.39 (-0.52 to 1.29) | 0.34 (-0.43 to 1.11) | 0.07 | 0.11 |
| LAG3 | 0.00 (-1.03 to 1.02) | 0.06 (-0.70 to 0.82) | 0.53 | 0.61 |
| **LAMA4** | **1.25 (0.24 to 2.26)** | **0.31 (-0.52 to 1.13)** | **0.04** | **0.08** |
| **LAYN** | **1.33 (0.26 to 2.40)** | **-0.38 (-1.31 to 0.54)** | **0.005** | **0.02** |
| **LTBP2** | **1.53 (0.55 to 2.52)** | **-0.12 (-0.94 to 0.70)** | **0.02** | **0.04** |
| **NOTCH3** | **1.56 (0.61 to 2.52)** | **0.29 (-0.57 to 1.14)** | **0.02** | **0.05** |
| **NT-proBNP** | **4.17 (3.08 to 5.26)** | **2.58 (1.68 to 3.47)** | **0.01** | **0.04** |
| **OGN** | **0.85 (-0.34 to 2.04)** | **-0.49 (-1.56 to 0.58)** | **0.02** | **0.05** |
| OXT | -0.46 (-1.43 to 0.51) | -0.68 (-1.41 to 0.05) | 0.90 | 0.92 |
| PDCD1 | 0.67 (-0.19 to 1.52) | 0.11 (-0.73 to 0.95) | 0.15 | 0.21 |
| **PDGFRA** | **1.36 (0.36 to 2.36)** | **0.39 (-0.38 to 1.16)** | **0.02** | **0.05** |
| PODXL2 | 0.70 (-0.26 to 1.67) | -0.14 (-1.01 to 0.73) | 0.07 | 0.12 |
| PROS1 | -0.49 (-1.46 to 0.47) | -0.79 (-1.57 to -0.01) | 0.62 | 0.67 |
| **ROR1** | **1.09 (0.07 to 2.11)** | **0.23 (-0.68 to 1.13)** | **0.02** | **0.05** |
| **S100G** | **0.33 (-0.67 to 1.33)** | **-0.82 (-1.59 to -0.05)** | **0.03** | **0.05** |
| **SCARF2** | **1.61 (0.57 to 2.64)** | **0.21 (-0.74 to 1.15)** | **0.02** | **0.04** |
| SIGLEC1 | 0.00 (-1.10 to 1.10) | -0.05 (-0.76 to 0.65) | 0.54 | 0.61 |
| SLAMF7 | -0.09 (-1.02 to 0.84) | 0.48 (-0.26 to 1.21) | 0.79 | 0.82 |
| TFPI2 | 0.54 (-0.49 to 1.57) | 0.30 (-0.58 to 1.18) | 0.13 | 0.19 |
| TGFBR3 | 0.55 (-0.34 to 1.45) | 0.48 (-0.28 to 1.25) | 0.20 | 0.26 |
| **THBS2** | **1.49 (0.37 to 2.60)** | **-0.55 (-1.37 to 0.27)** | **0.002** | **0.01** |
| **THBS4** | **1.81 (0.82 to 2.81)** | **-0.13 (-0.95 to 0.69)** | **0.003** | **0.02** |
| TIGIT | -0.27 (-1.26 to 0.72) | 0.11 (-0.72 to 0.93) | 0.70 | 0.75 |
| **TIMP1** | **1.49 (0.44 to 2.55)** | **-0.13 (-0.99 to 0.74)** | **0.003** | **0.02** |
| TNFRSF13B | 0.35 (-0.62 to 1.32) | 0.44 (-0.36 to 1.24) | 0.92 | 0.92 |
| **TNFRSF14** | **1.00 (-0.03 to 2.03)** | **-0.64 (-1.59 to 0.32)** | **0.005** | **0.02** |
| TNFRSF17 | 0.25 (-0.80 to 1.29) | 0.39 (-0.40 to 1.17) | 0.89 | 0.92 |
| **TNFRSF1B** | **1.03 (0.01 to 2.06)** | **-0.59 (-1.55 to 0.37)** | **0.001** | **0.01** |
| **TNFRSF21** | **0.78 (-0.22 to 1.79)** | **-0.34 (-1.21 to 0.53)** | **0.007** | **0.03** |
| **TNFRSF4** | **0.98 (0.03 to 1.94)** | **-0.33 (-1.24 to 0.58)** | **0.002** | **0.01** |
| TNFRSF8 | 0.25 (-0.72 to 1.22) | 0.18 (-0.74 to 1.10) | 0.63 | 0.69 |
| **TNFRSF9** | **1.26 (0.26 to 2.25)** | **-0.03 (-0.88 to 0.83)** | **0.02** | **0.05** |
| **VCAM1** | **1.36 (0.31 to 2.40)** | **-0.39 (-1.14 to 0.35)** | **0.001** | **0.01** |

Estimated using linear regression with robust variance, adjusting for study site, age, sex, race/ethnicity, education, body mass index, systolic blood pressure, anti-hypertensive medication, hyperlipidemia, diabetes, smoking, and estimated glomerular filtration rate. Subgroups defined by dichotomization at median age in MESA cross-sectional analysis sample (67 years).
SD=standard deviation; CI=confidence interval; FDR=false discovery rate (Benjamini-Hochberg).

**SUPPLEMENTAL TABLE S16.**
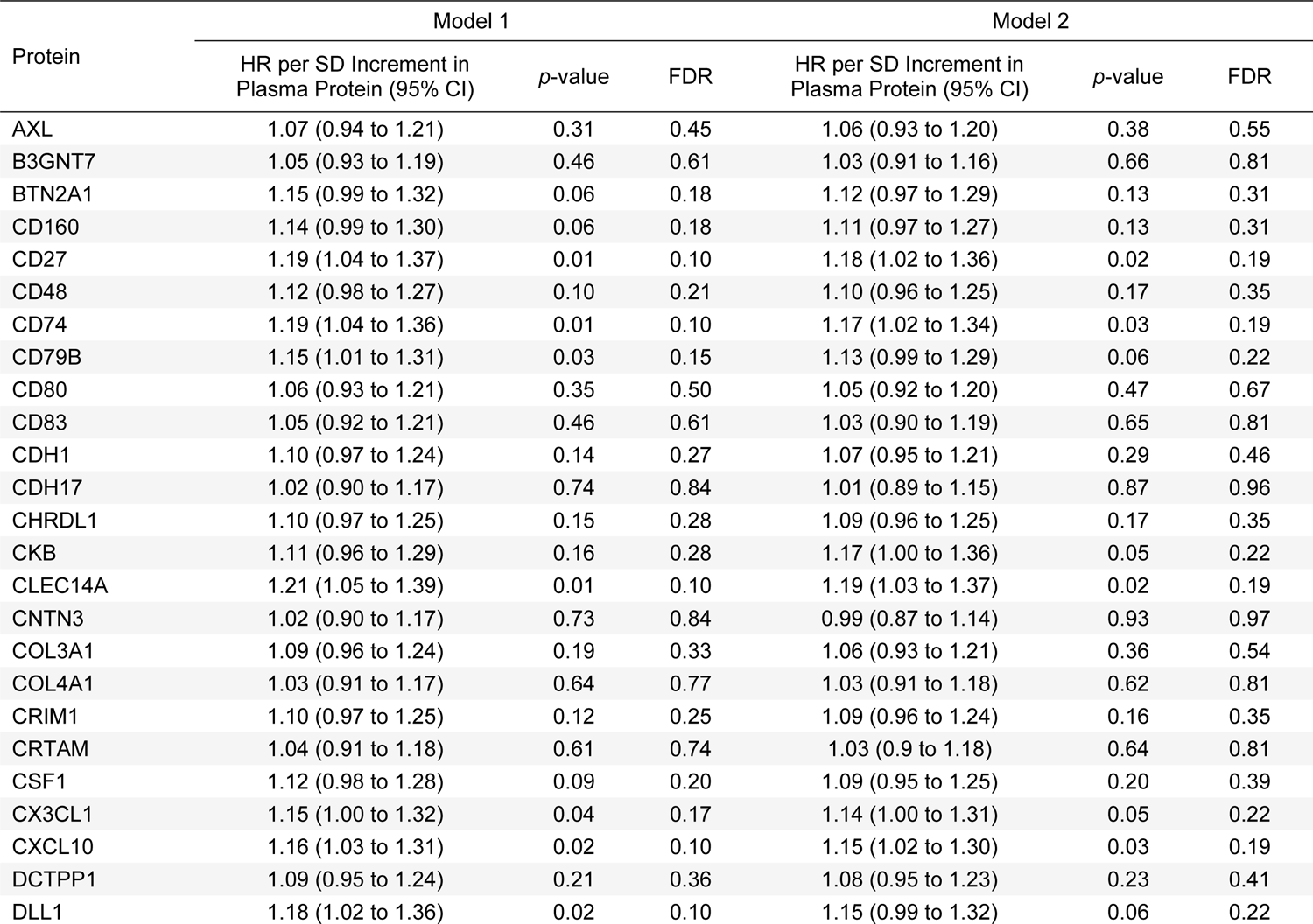
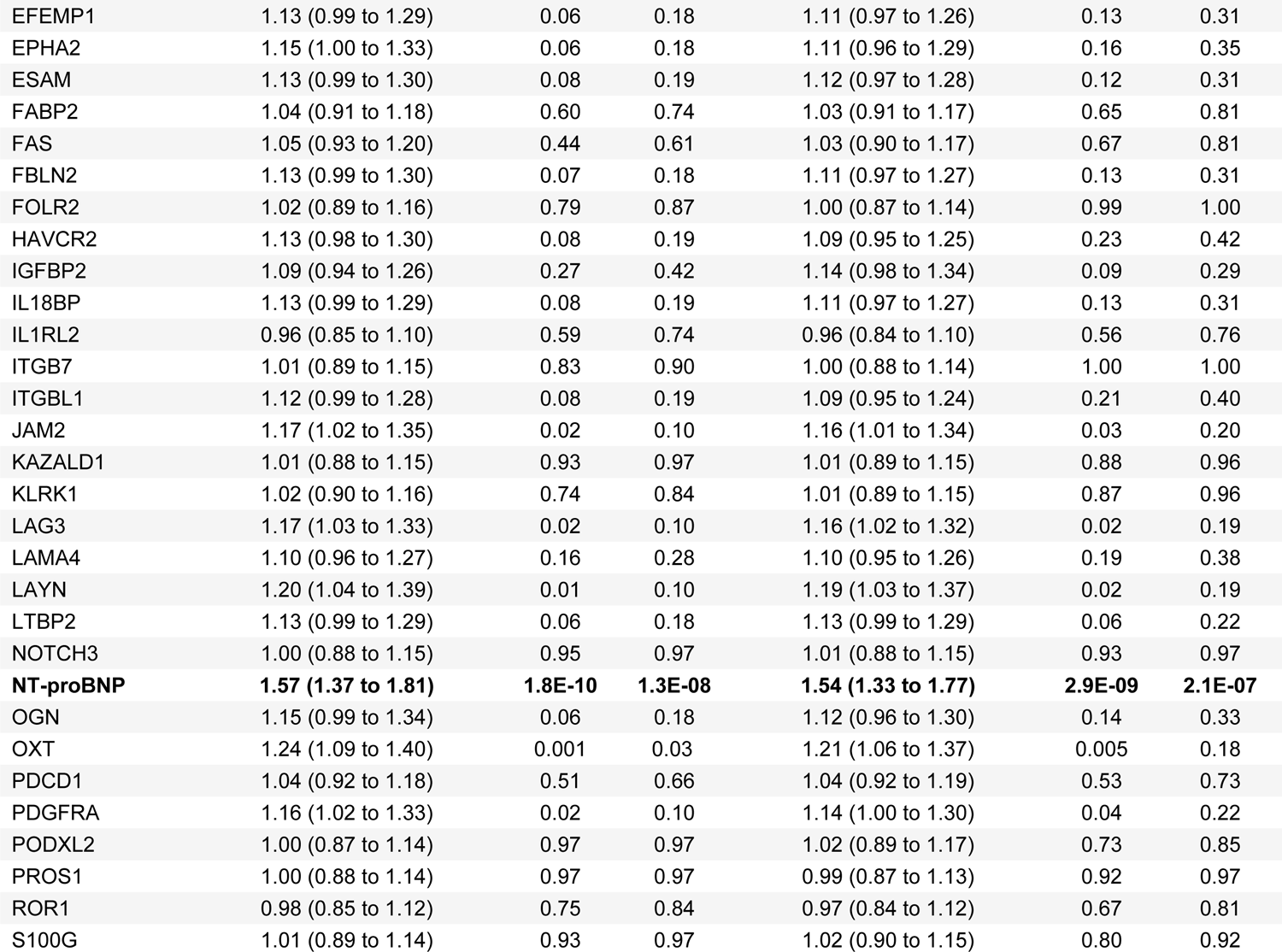
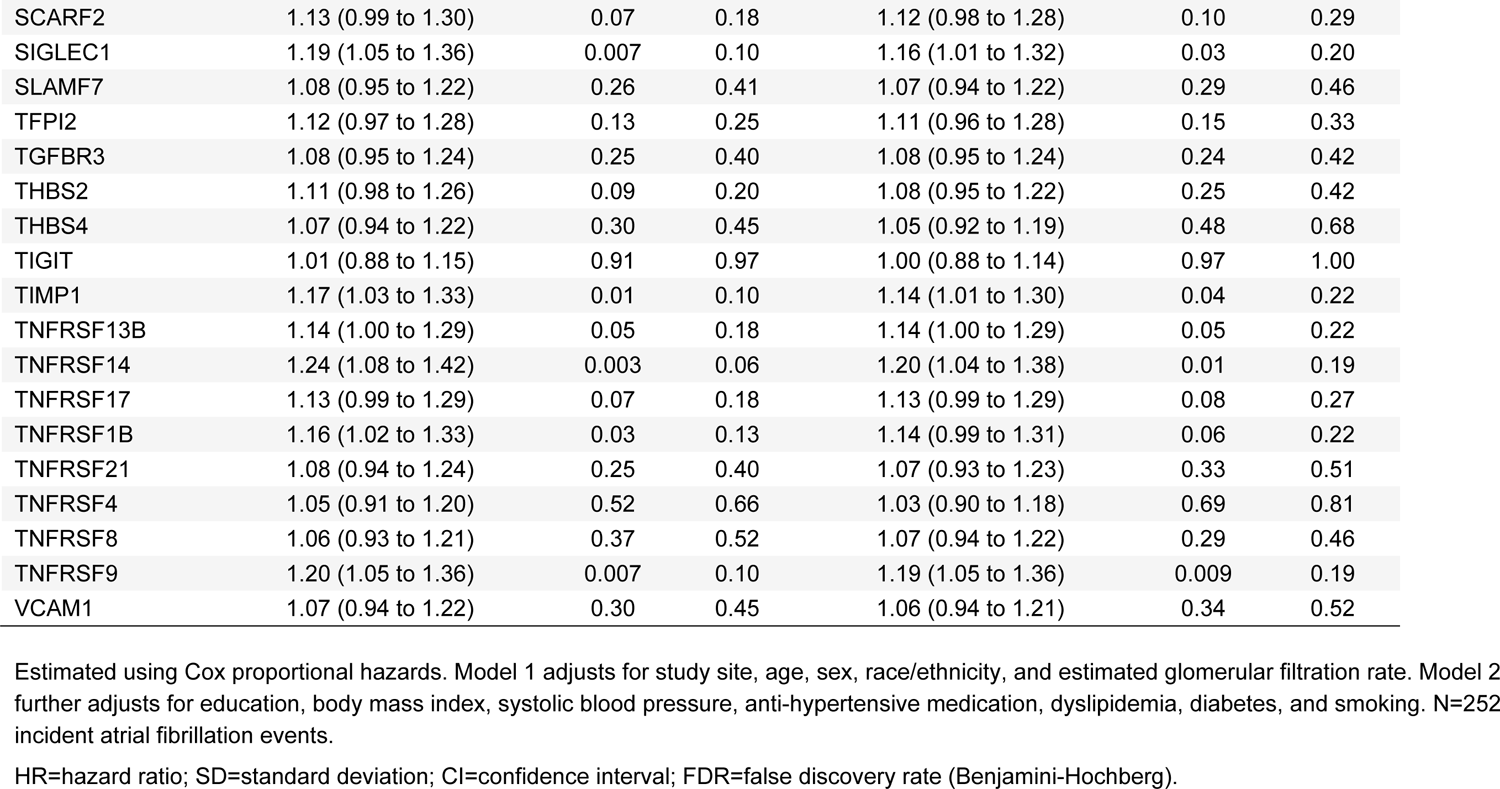
Association between proteins of interest and incident atrial fibrillation in the Multi-Ethnic Study of Atherosclerosis (median age 67 years at the beginning of follow-up, *n*=2185)

**SUPPLEMENTAL TABLE S17.**
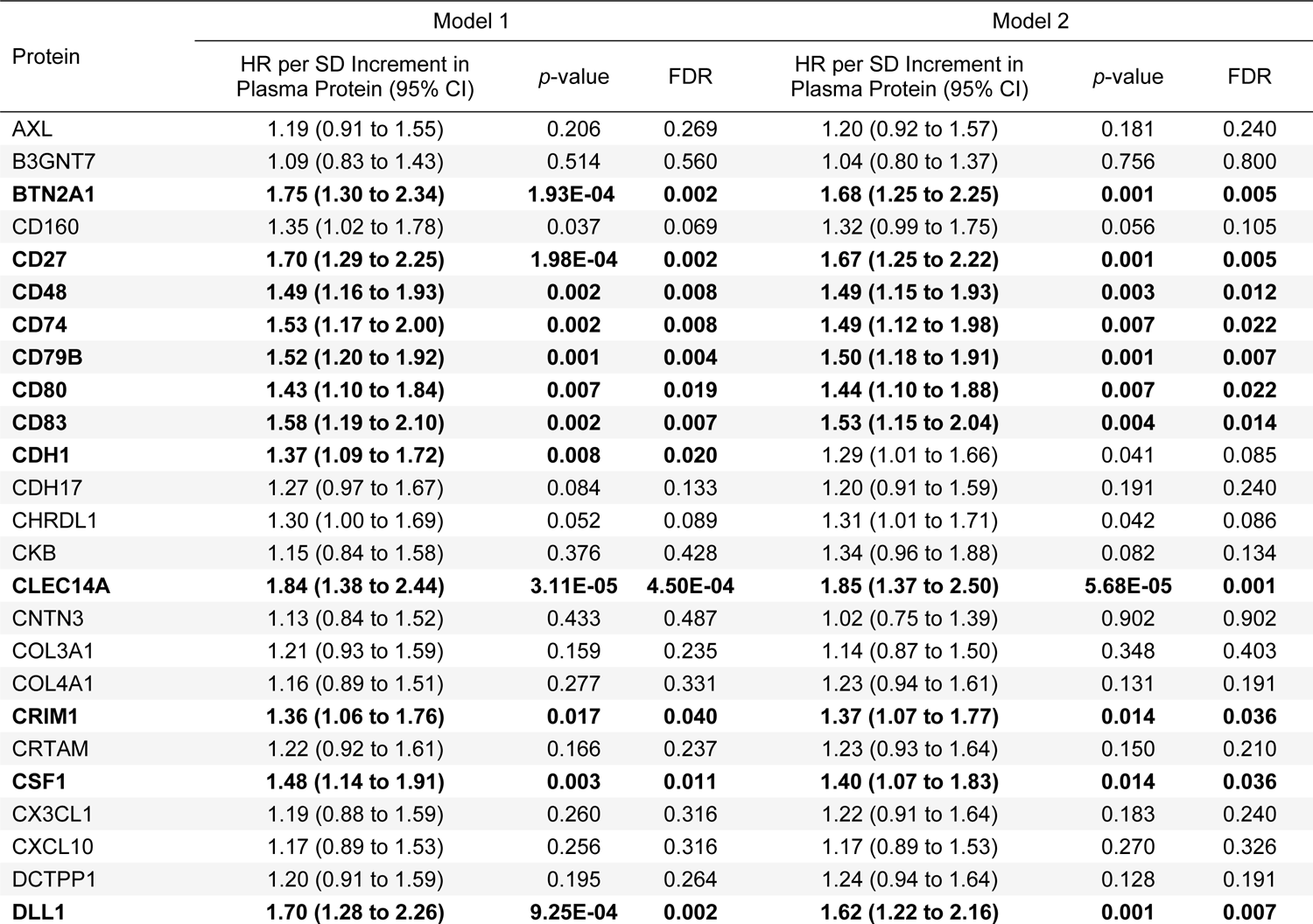
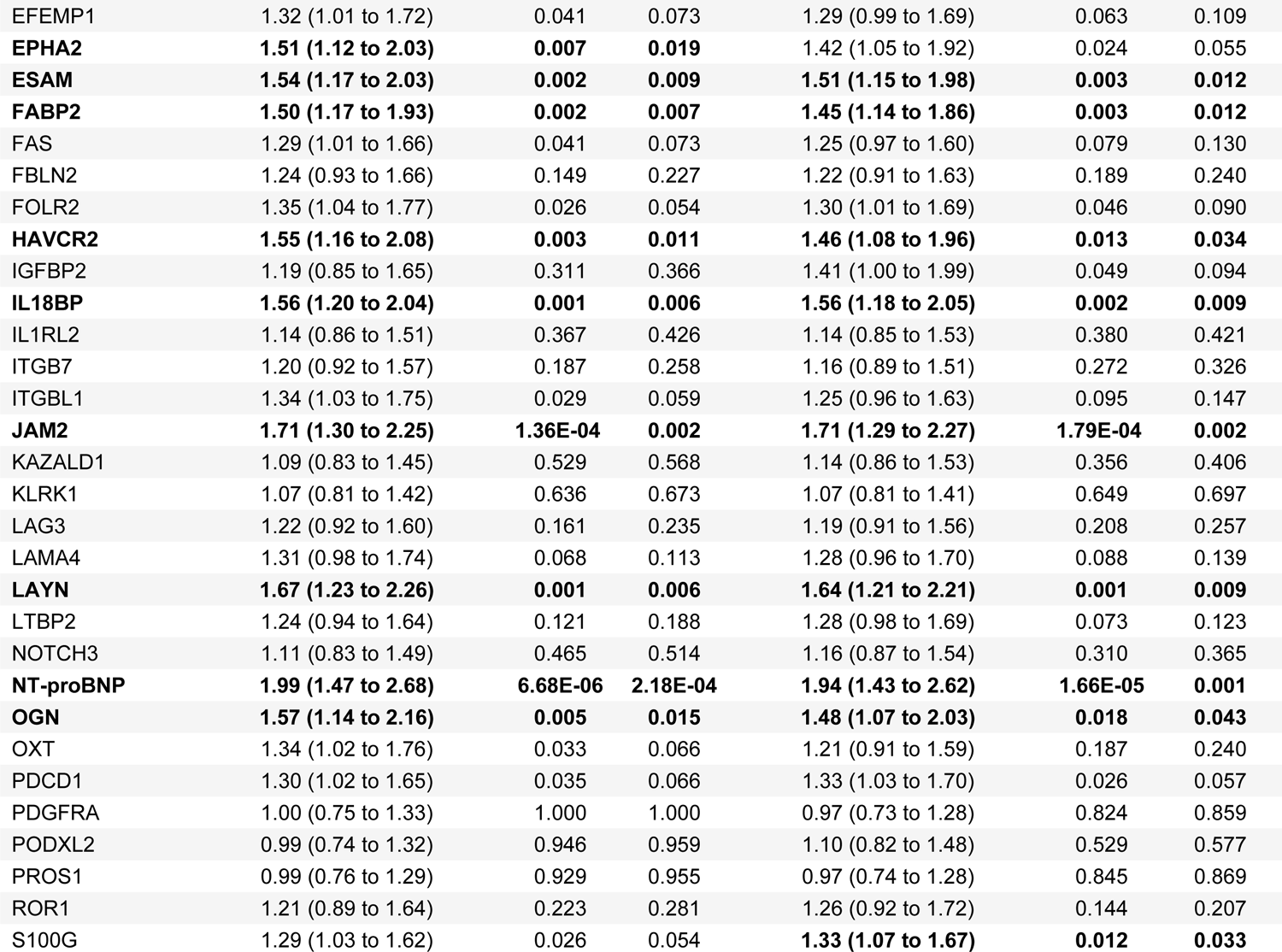
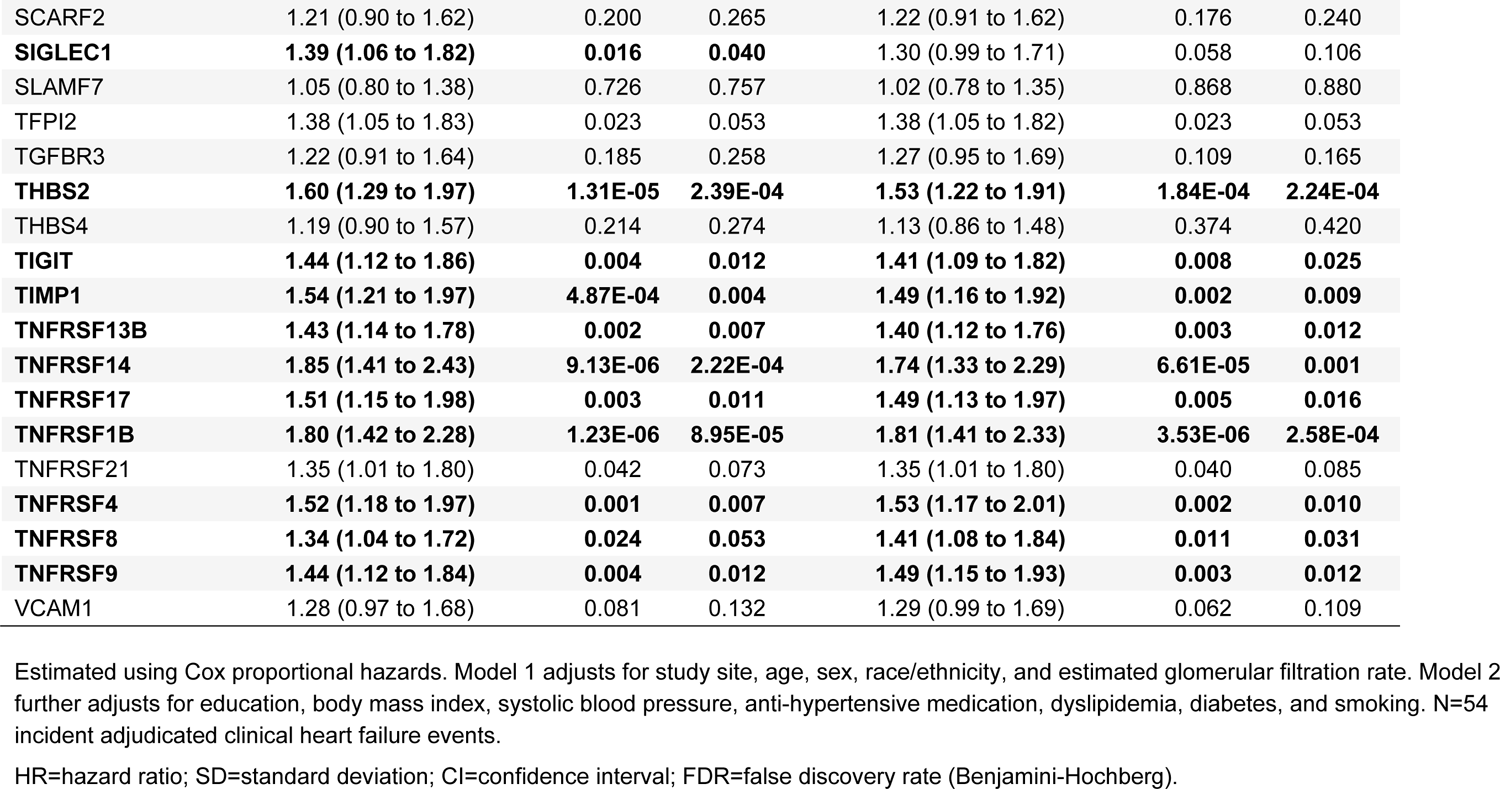
Association between proteins of interest and incident clinical heart failure in the Multi-Ethnic Study of Atherosclerosis (median age 67 years at the beginning of follow-up, *n*=2273)

| Protein | Model 1 |  |  | Model 2 |  |  |
| --- | --- | --- | --- | --- | --- | --- |
|  | HR per SD Increment in Plasma Protein (95% CI) | *p*-value | FDR | HR per SD Increment in Plasma Protein (95% CI) | *p*-value | FDR |
| AXL | 1.19 (0.91 to 1.55) | 0.206 | 0.269 | 1.20 (0.92 to 1.57) | 0.181 | 0.240 |
| B3GNT7 | 1.09 (0.83 to 1.43) | 0.514 | 0.560 | 1.04 (0.80 to 1.37) | 0.756 | 0.800 |
| **BTN2A1** | **1.75 (1.30 to 2.34)** | **1.93E-04** | **0.002** | **1.68 (1.25 to 2.25)** | **0.001** | **0.005** |
| CD160 | 1.35 (1.02 to 1.78) | 0.037 | 0.069 | 1.32 (0.99 to 1.75) | 0.056 | 0.105 |
| **CD27** | **1.70 (1.29 to 2.25)** | **1.98E-04** | **0.002** | **1.67 (1.25 to 2.22)** | **0.001** | **0.005** |
| **CD48** | **1.49 (1.16 to 1.93)** | **0.002** | **0.008** | **1.49 (1.15 to 1.93)** | **0.003** | **0.012** |
| **CD74** | **1.53 (1.17 to 2.00)** | **0.002** | **0.008** | **1.49 (1.12 to 1.98)** | **0.007** | **0.022** |
| **CD79B** | **1.52 (1.20 to 1.92)** | **0.001** | **0.004** | **1.50 (1.18 to 1.91)** | **0.001** | **0.007** |
| **CD80** | **1.43 (1.10 to 1.84)** | **0.007** | **0.019** | **1.44 (1.10 to 1.88)** | **0.007** | **0.022** |
| **CD83** | **1.58 (1.19 to 2.10)** | **0.002** | **0.007** | **1.53 (1.15 to 2.04)** | **0.004** | **0.014** |
| **CDH1** | **1.37 (1.09 to 1.72)** | **0.008** | **0.020** | 1.29 (1.01 to 1.66) | 0.041 | 0.085 |
| CDH17 | 1.27 (0.97 to 1.67) | 0.084 | 0.133 | 1.20 (0.91 to 1.59) | 0.191 | 0.240 |
| CHRD1 | 1.30 (1.00 to 1.69) | 0.052 | 0.089 | 1.31 (1.01 to 1.71) | 0.042 | 0.086 |
| CKB | 1.15 (0.84 to 1.58) | 0.376 | 0.428 | 1.34 (0.96 to 1.88) | 0.082 | 0.134 |
| **CLEC14A** | **1.84 (1.38 to 2.44)** | **3.11E-05** | **4.50E-04** | **1.85 (1.37 to 2.50)** | **5.68E-05** | **0.001** |
| CNTN3 | 1.13 (0.84 to 1.52) | 0.433 | 0.487 | 1.02 (0.75 to 1.39) | 0.902 | 0.902 |
| COL3A1 | 1.21 (0.93 to 1.59) | 0.159 | 0.235 | 1.14 (0.87 to 1.50) | 0.348 | 0.403 |
| COL4A1 | 1.16 (0.89 to 1.51) | 0.277 | 0.331 | 1.23 (0.94 to 1.61) | 0.131 | 0.191 |
| **CRIM1** | **1.36 (1.06 to 1.76)** | **0.017** | **0.040** | **1.37 (1.07 to 1.77)** | **0.014** | **0.036** |
| CRTAM | 1.22 (0.92 to 1.61) | 0.166 | 0.237 | 1.23 (0.93 to 1.64) | 0.150 | 0.210 |
| **CSF1** | **1.48 (1.14 to 1.91)** | **0.003** | **0.011** | **1.40 (1.07 to 1.83)** | **0.014** | **0.036** |
| CX3CL1 | 1.19 (0.88 to 1.59) | 0.260 | 0.316 | 1.22 (0.91 to 1.64) | 0.183 | 0.240 |
| CXCL10 | 1.17 (0.89 to 1.53) | 0.256 | 0.316 | 1.17 (0.89 to 1.53) | 0.270 | 0.326 |
| DCTPP1 | 1.20 (0.91 to 1.59) | 0.195 | 0.264 | 1.24 (0.94 to 1.64) | 0.128 | 0.191 |
| **DLL1** | **1.70 (1.28 to 2.26)** | **9.25E-04** | **0.002** | **1.62 (1.22 to 2.16)** | **0.001** | **0.007** |
| EFEMP1 | 1.32 (1.01 to 1.72) | 0.041 | 0.073 | 1.29 (0.99 to 1.69) | 0.063 | 0.109 |
| **EPHA2** | **1.51 (1.12 to 2.03)** | **0.007** | **0.019** | 1.42 (1.05 to 1.92) | 0.024 | 0.055 |
| **ESAM** | **1.54 (1.17 to 2.03)** | **0.002** | **0.009** | **1.51 (1.15 to 1.98)** | **0.003** | **0.012** |
| **FABP2** | **1.50 (1.17 to 1.93)** | **0.002** | **0.007** | **1.45 (1.14 to 1.86)** | **0.003** | **0.012** |
| FAS | 1.29 (1.01 to 1.66) | 0.041 | 0.073 | 1.25 (0.97 to 1.60) | 0.079 | 0.130 |
| FBLN2 | 1.24 (0.93 to 1.66) | 0.149 | 0.227 | 1.22 (0.91 to 1.63) | 0.189 | 0.240 |
| FOLR2 | 1.35 (1.04 to 1.77) | 0.026 | 0.054 | 1.30 (1.01 to 1.69) | 0.046 | 0.090 |
| **HAVCR2** | **1.55 (1.16 to 2.08)** | **0.003** | **0.011** | **1.46 (1.08 to 1.96)** | **0.013** | **0.034** |
| IGFBP2 | 1.19 (0.85 to 1.65) | 0.311 | 0.366 | 1.41 (1.00 to 1.99) | 0.049 | 0.094 |
| **IL18BP** | **1.56 (1.20 to 2.04)** | **0.001** | **0.006** | **1.56 (1.18 to 2.05)** | **0.002** | **0.009** |
| IL1RL2 | 1.14 (0.86 to 1.51) | 0.367 | 0.426 | 1.14 (0.85 to 1.53) | 0.380 | 0.421 |
| ITGB7 | 1.20 (0.92 to 1.57) | 0.187 | 0.258 | 1.16 (0.89 to 1.51) | 0.272 | 0.326 |
| ITGBL1 | 1.34 (1.03 to 1.75) | 0.029 | 0.059 | 1.25 (0.96 to 1.63) | 0.095 | 0.147 |
| **JAM2** | **1.71 (1.30 to 2.25)** | **1.36E-04** | **0.002** | **1.71 (1.29 to 2.27)** | **1.79E-04** | **0.002** |
| KAZALD1 | 1.09 (0.83 to 1.45) | 0.529 | 0.568 | 1.14 (0.86 to 1.53) | 0.356 | 0.406 |
| KLRK1 | 1.07 (0.81 to 1.42) | 0.636 | 0.673 | 1.07 (0.81 to 1.41) | 0.649 | 0.697 |
| LAG3 | 1.22 (0.92 to 1.60) | 0.161 | 0.235 | 1.19 (0.91 to 1.56) | 0.208 | 0.257 |
| LAMA4 | 1.31 (0.98 to 1.74) | 0.068 | 0.113 | 1.28 (0.96 to 1.70) | 0.088 | 0.139 |
| **LAYN** | **1.67 (1.23 to 2.26)** | **0.001** | **0.006** | **1.64 (1.21 to 2.21)** | **0.001** | **0.009** |
| LTBP2 | 1.24 (0.94 to 1.64) | 0.121 | 0.188 | 1.28 (0.98 to 1.69) | 0.073 | 0.123 |
| NOTCH3 | 1.11 (0.83 to 1.49) | 0.465 | 0.514 | 1.16 (0.87 to 1.54) | 0.310 | 0.365 |
| **NT-proBNP** | **1.99 (1.47 to 2.68)** | **6.68E-06** | **2.18E-04** | **1.94 (1.43 to 2.62)** | **1.66E-05** | **0.001** |
| **OGN** | **1.57 (1.14 to 2.16)** | **0.005** | **0.015** | **1.48 (1.07 to 2.03)** | **0.018** | **0.043** |
| OXT | 1.34 (1.02 to 1.76) | 0.033 | 0.066 | 1.21 (0.91 to 1.59) | 0.187 | 0.240 |
| PDCD1 | 1.30 (1.02 to 1.65) | 0.035 | 0.066 | 1.33 (1.03 to 1.70) | 0.026 | 0.057 |
| PDGFRA | 1.00 (0.75 to 1.33) | 1.000 | 1.000 | 0.97 (0.73 to 1.28) | 0.824 | 0.859 |
| PODXL2 | 0.99 (0.74 to 1.32) | 0.946 | 0.959 | 1.10 (0.82 to 1.48) | 0.529 | 0.577 |
| PROS1 | 0.99 (0.76 to 1.29) | 0.929 | 0.955 | 0.97 (0.74 to 1.28) | 0.845 | 0.869 |
| ROR1 | 1.21 (0.89 to 1.64) | 0.223 | 0.281 | 1.26 (0.92 to 1.72) | 0.144 | 0.207 |
| S100G | 1.29 (1.03 to 1.62) | 0.026 | 0.054 | **1.33 (1.07 to 1.67)** | **0.012** | **0.033** |
| SCARF2 | 1.21 (0.90 to 1.62) | 0.200 | 0.265 | 1.22 (0.91 to 1.62) | 0.176 | 0.240 |
| **SIGLEC1** | **1.39 (1.06 to 1.82)** | **0.016** | **0.040** | 1.30 (0.99 to 1.71) | 0.058 | 0.106 |
| SLAMF7 | 1.05 (0.80 to 1.38) | 0.726 | 0.757 | 1.02 (0.78 to 1.35) | 0.868 | 0.880 |
| TFPI2 | 1.38 (1.05 to 1.83) | 0.023 | 0.053 | 1.38 (1.05 to 1.82) | 0.023 | 0.053 |
| TGFBR3 | 1.22 (0.91 to 1.64) | 0.185 | 0.258 | 1.27 (0.95 to 1.69) | 0.109 | 0.165 |
| **THBS2** | **1.60 (1.29 to 1.97)** | **1.31E-05** | **2.39E-04** | **1.53 (1.22 to 1.91)** | **1.84E-04** | **2.24E-04** |
| THBS4 | 1.19 (0.90 to 1.57) | 0.214 | 0.274 | 1.13 (0.86 to 1.48) | 0.374 | 0.420 |
| **TIGIT** | **1.44 (1.12 to 1.86)** | **0.004** | **0.012** | **1.41 (1.09 to 1.82)** | **0.008** | **0.025** |
| **TIMP1** | **1.54 (1.21 to 1.97)** | **4.87E-04** | **0.004** | **1.49 (1.16 to 1.92)** | **0.002** | **0.009** |
| **TNFRSF13B** | **1.43 (1.14 to 1.78)** | **0.002** | **0.007** | **1.40 (1.12 to 1.76)** | **0.003** | **0.012** |
| **TNFRSF14** | **1.85 (1.41 to 2.43)** | **9.13E-06** | **2.22E-04** | **1.74 (1.33 to 2.29)** | **6.61E-05** | **0.001** |
| **TNFRSF17** | **1.51 (1.15 to 1.98)** | **0.003** | **0.011** | **1.49 (1.13 to 1.97)** | **0.005** | **0.016** |
| **TNFRSF1B** | **1.80 (1.42 to 2.28)** | **1.23E-06** | **8.95E-05** | **1.81 (1.41 to 2.33)** | **3.53E-06** | **2.58E-04** |
| TNFRSF21 | 1.35 (1.01 to 1.80) | 0.042 | 0.073 | 1.35 (1.01 to 1.80) | 0.040 | 0.085 |
| **TNFRSF4** | **1.52 (1.18 to 1.97)** | **0.001** | **0.007** | **1.53 (1.17 to 2.01)** | **0.002** | **0.010** |
| **TNFRSF8** | **1.34 (1.04 to 1.72)** | **0.024** | **0.053** | **1.41 (1.08 to 1.84)** | **0.011** | **0.031** |
| **TNFRSF9** | **1.44 (1.12 to 1.84)** | **0.004** | **0.012** | **1.49 (1.15 to 1.93)** | **0.003** | **0.012** |
| VCAM1 | 1.28 (0.97 to 1.68) | 0.081 | 0.132 | 1.29 (0.99 to 1.69) | 0.062 | 0.109 |

Estimated using Cox proportional hazards. Model 1 adjusts for study site, age, sex, race/ethnicity, and estimated glomerular filtration rate. Model 2 further adjusts for education, body mass index, systolic blood pressure, anti-hypertensive medication, dyslipidemia, diabetes, and smoking. N=54 incident adjudicated clinical heart failure events.
HR=hazard ratio; SD=standard deviation; CI=confidence interval; FDR=false discovery rate (Benjamini-Hochberg).

**SUPPLEMENTAL FIGURE S1A.**
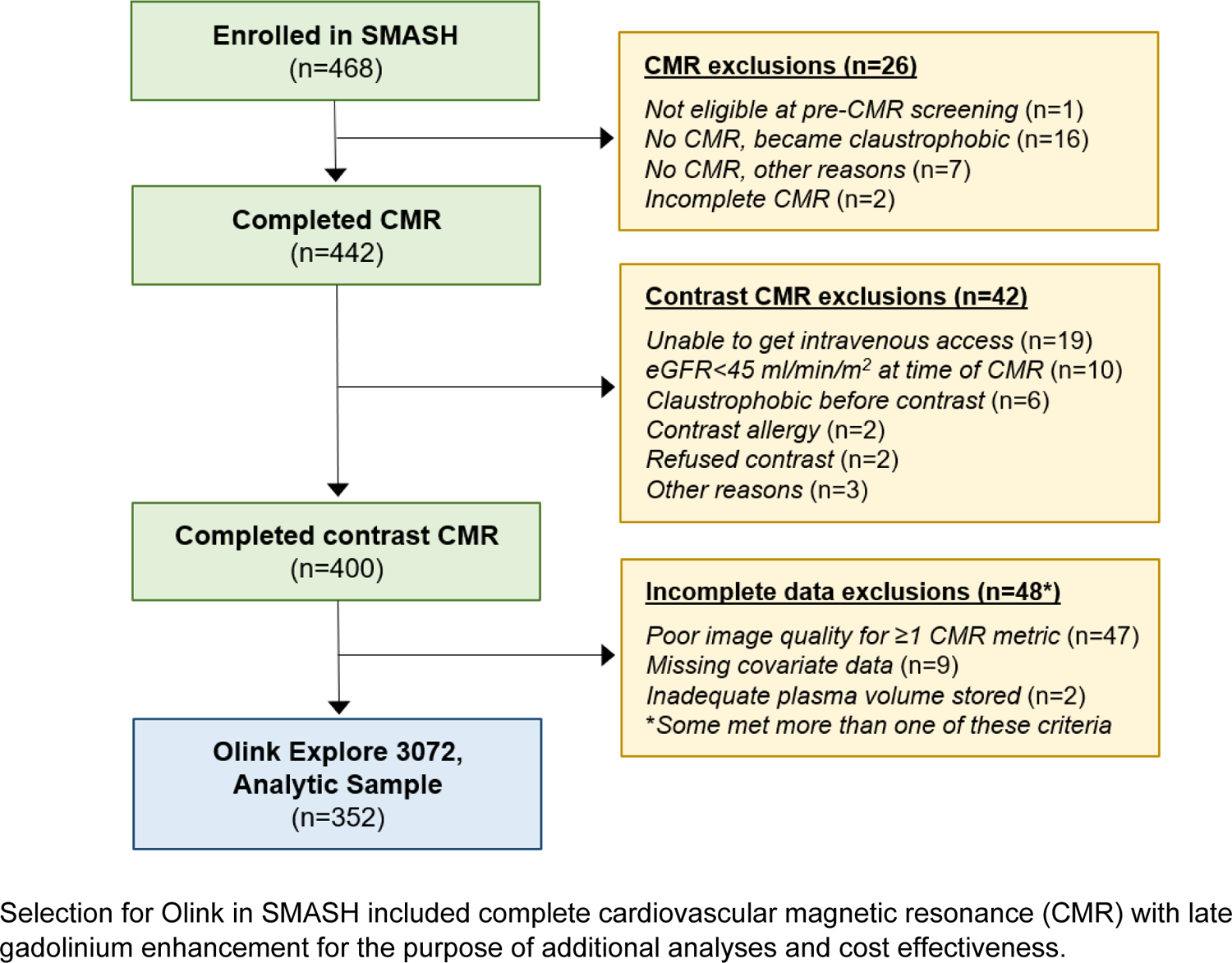
Flow diagram of SMASH study participants included in analysis sample.

**SUPPLEMENTAL FIGURE S1B.**
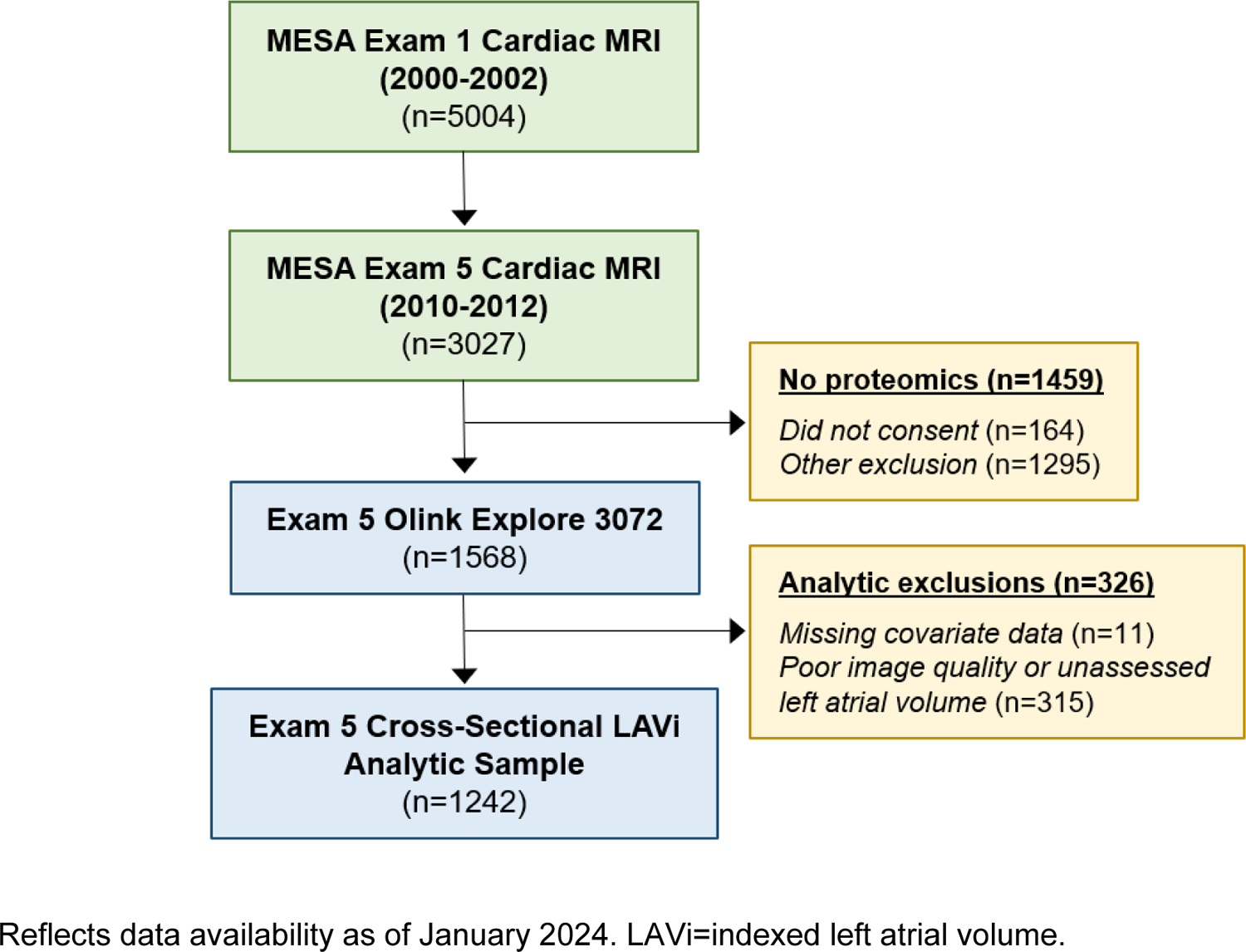
Flow diagram of MESA study participants included in cross-sectional analysis samples.

**SUPPLEMENTAL FIGURE S1C.**
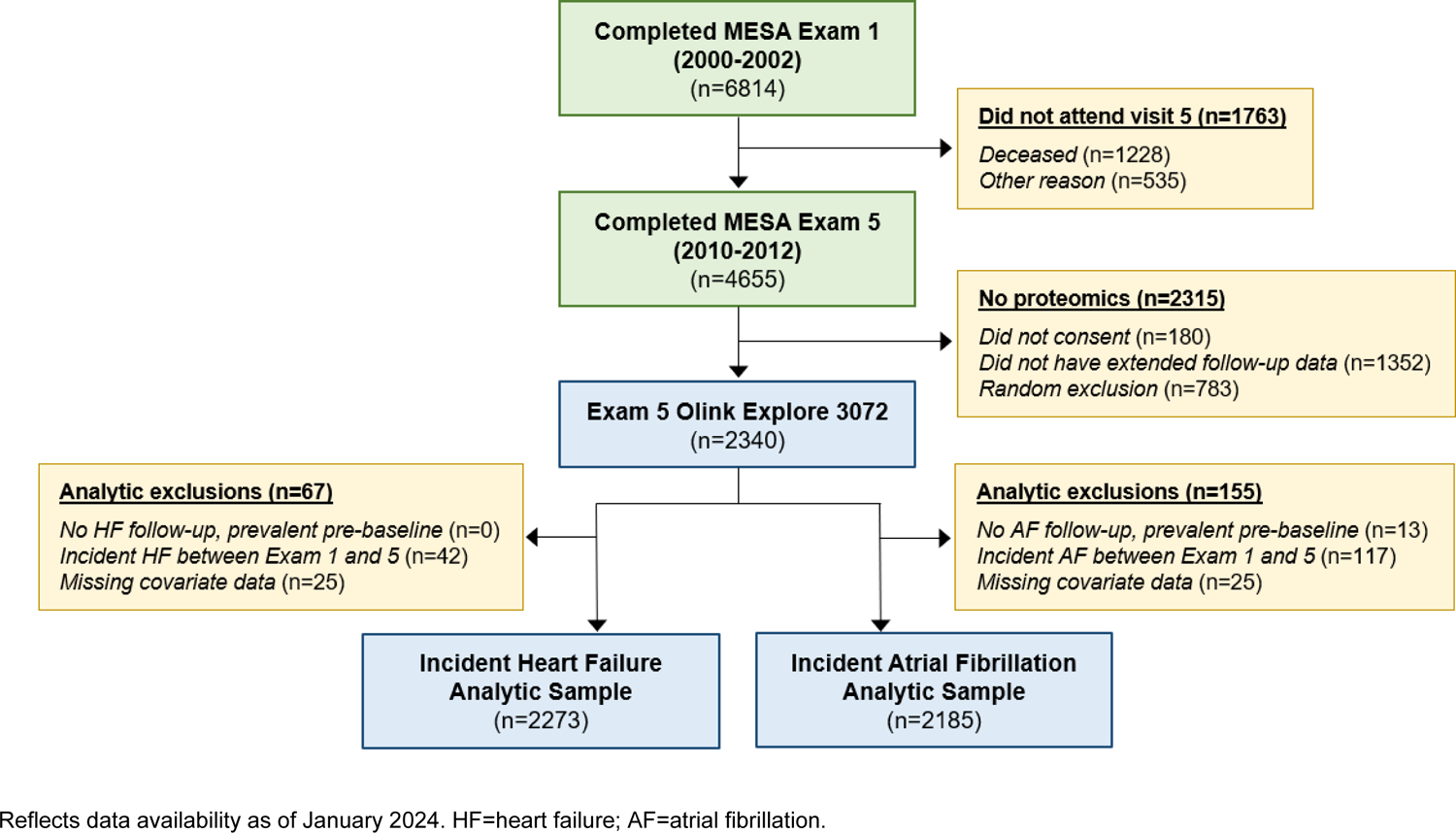
Flow diagram of MESA study participants included in longitudinal analysis samples.

**SUPPLEMENTAL FIGURE S2.**
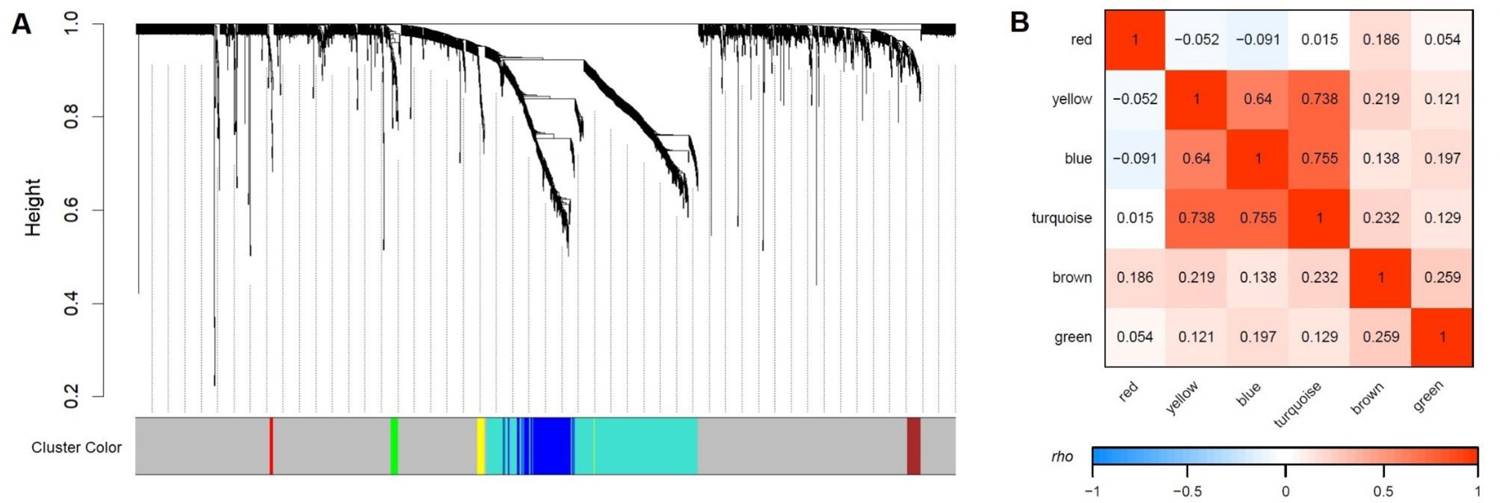
Clusters of proteins agnostically defined using weighted gene co-expression network analysis. (A) Dendrogram of protein clusters; gray indicates unclustered proteins; (B) Spearman’s correlations between cluster eigenprotein values, i.e., first principal component of cluster-specific proteins.

**SUPPLEMENTAL FIGURE S3.**
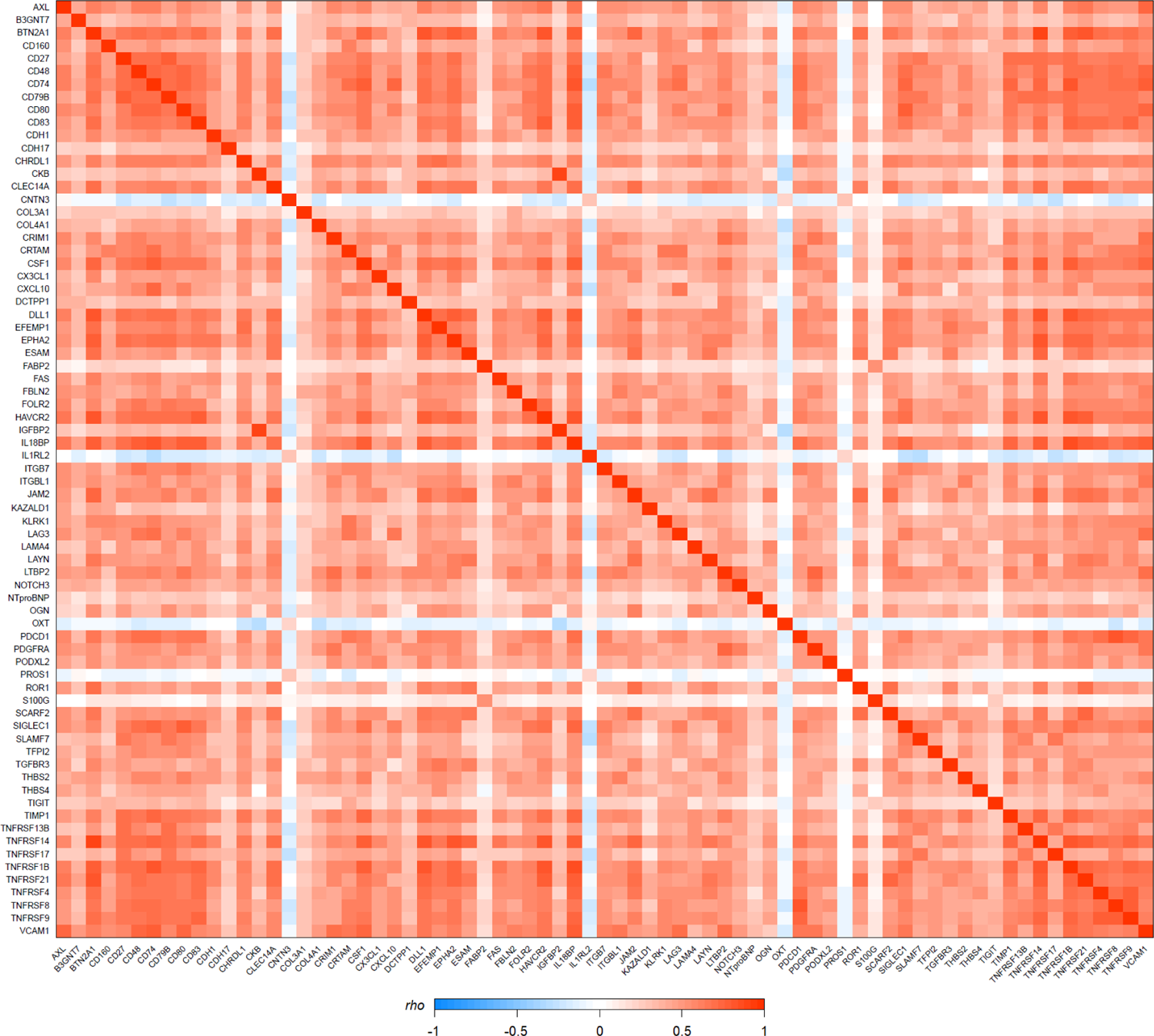
Spearman’s correlation between plasma abundances of 73 proteins independently associated with both HIV seropositivity and incremental indexed left atrial volume, same directionality, among PLWH and PWOH in the United States (*n*=352).

**SUPPLEMENTAL FIGURE S4.**
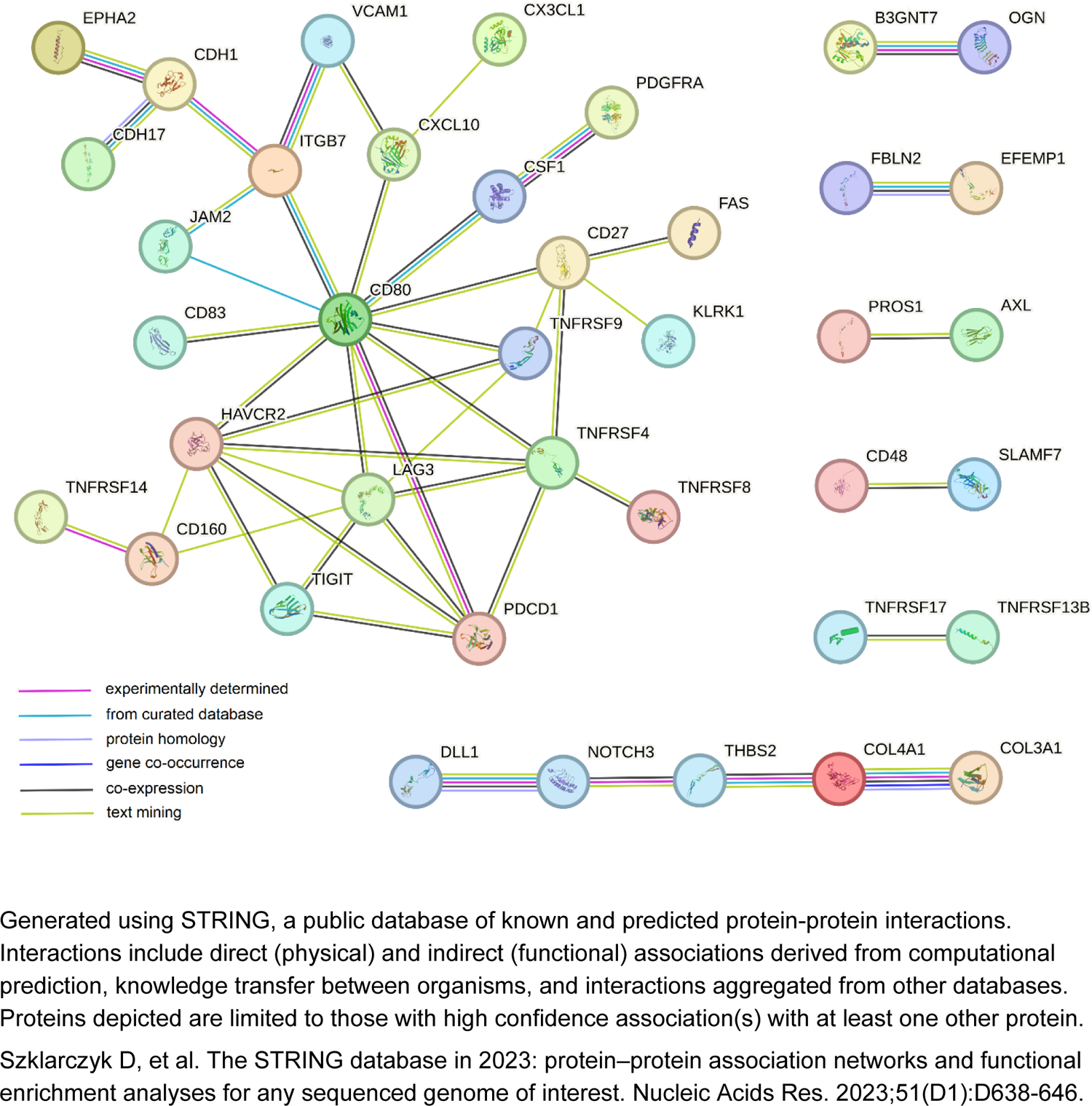
Interaction network of 73 proteins independently associated with both HIV seropositivity and incremental indexed left atrial volume, same directionality.

**SUPPLEMENTAL FIGURE S5.**
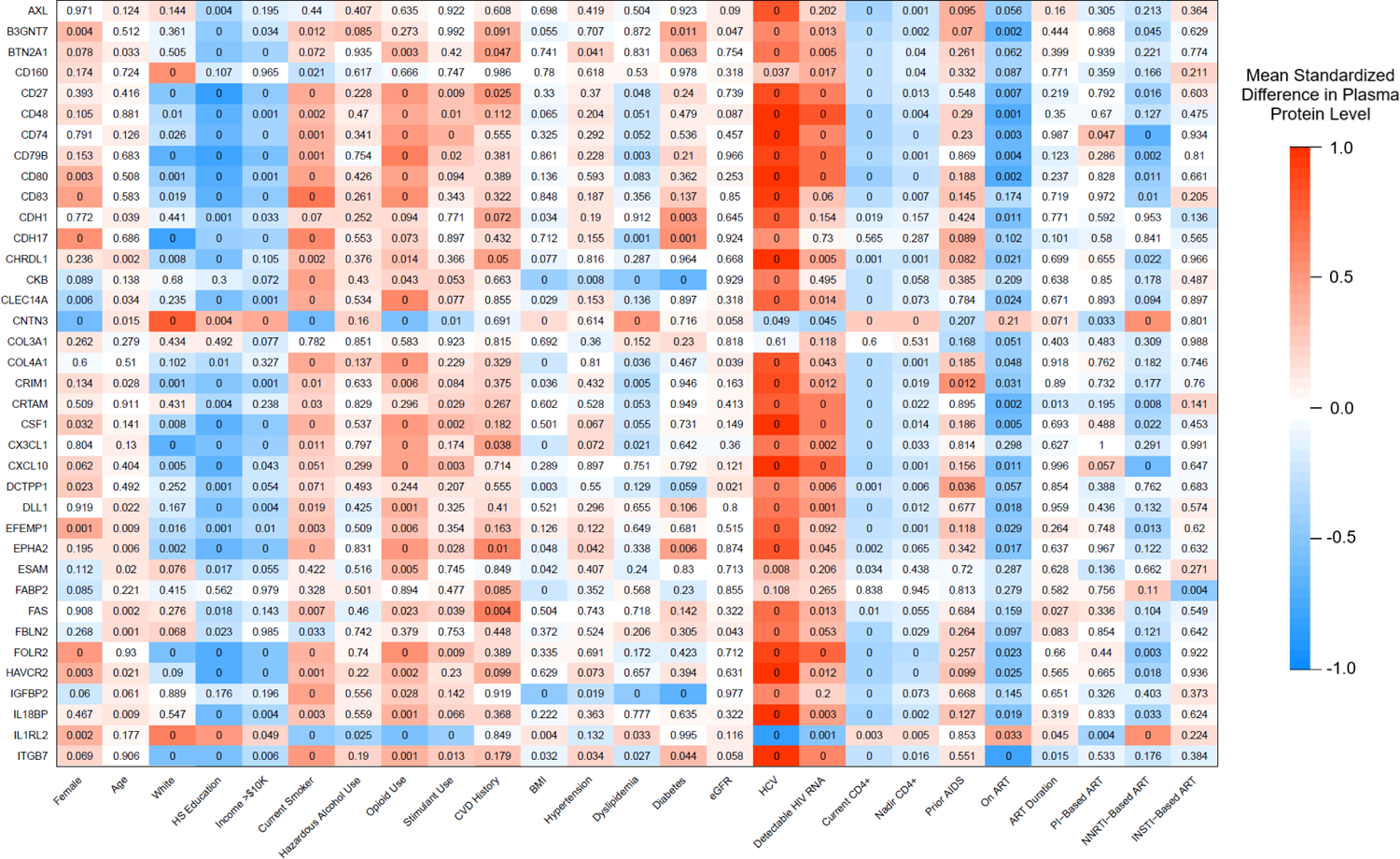
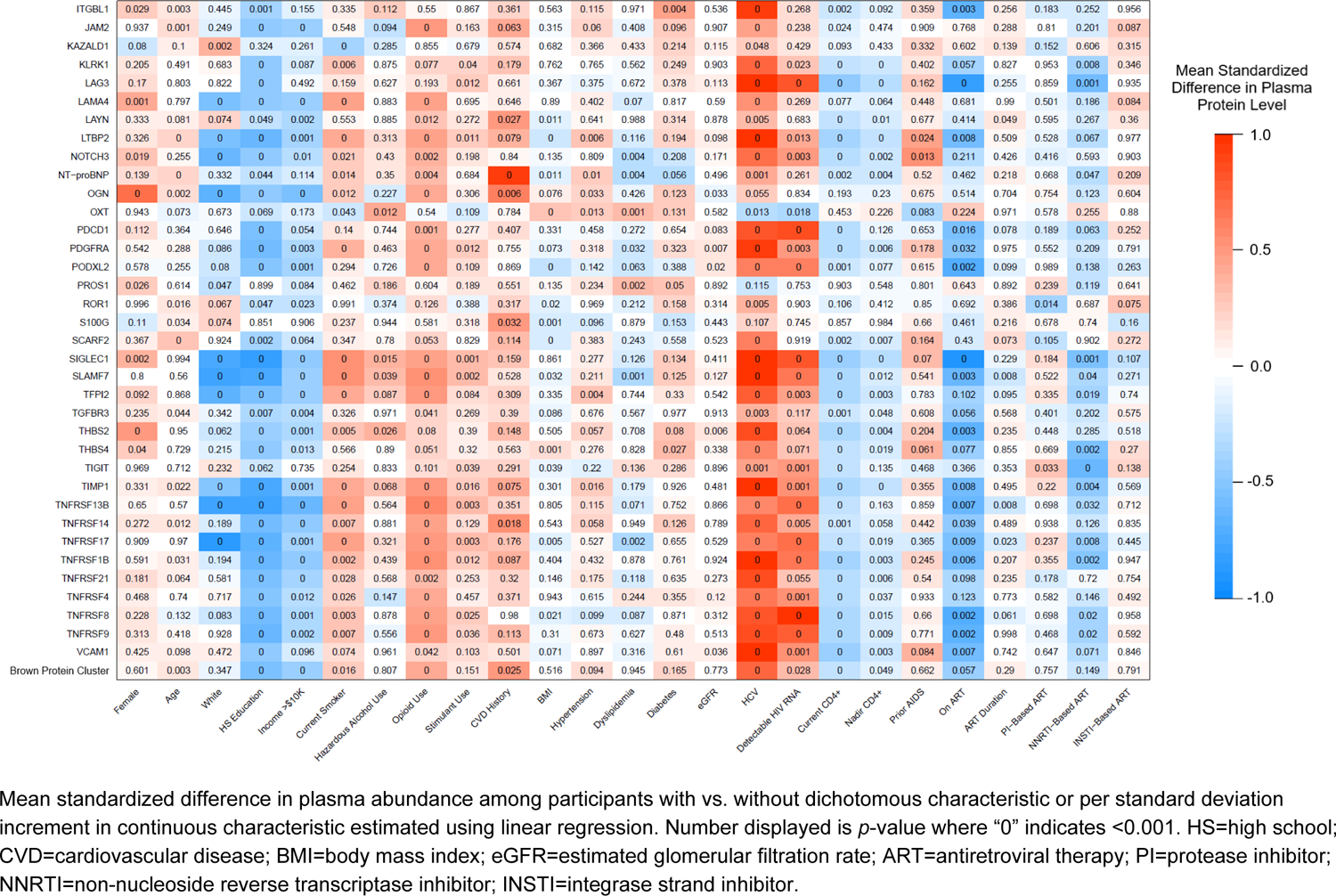
Association between identified HIV-associated proteomic signature of left atrial size and clinical characteristics in SMASH (*n*=352)

**SUPPLEMENTAL FIGURE S6.**
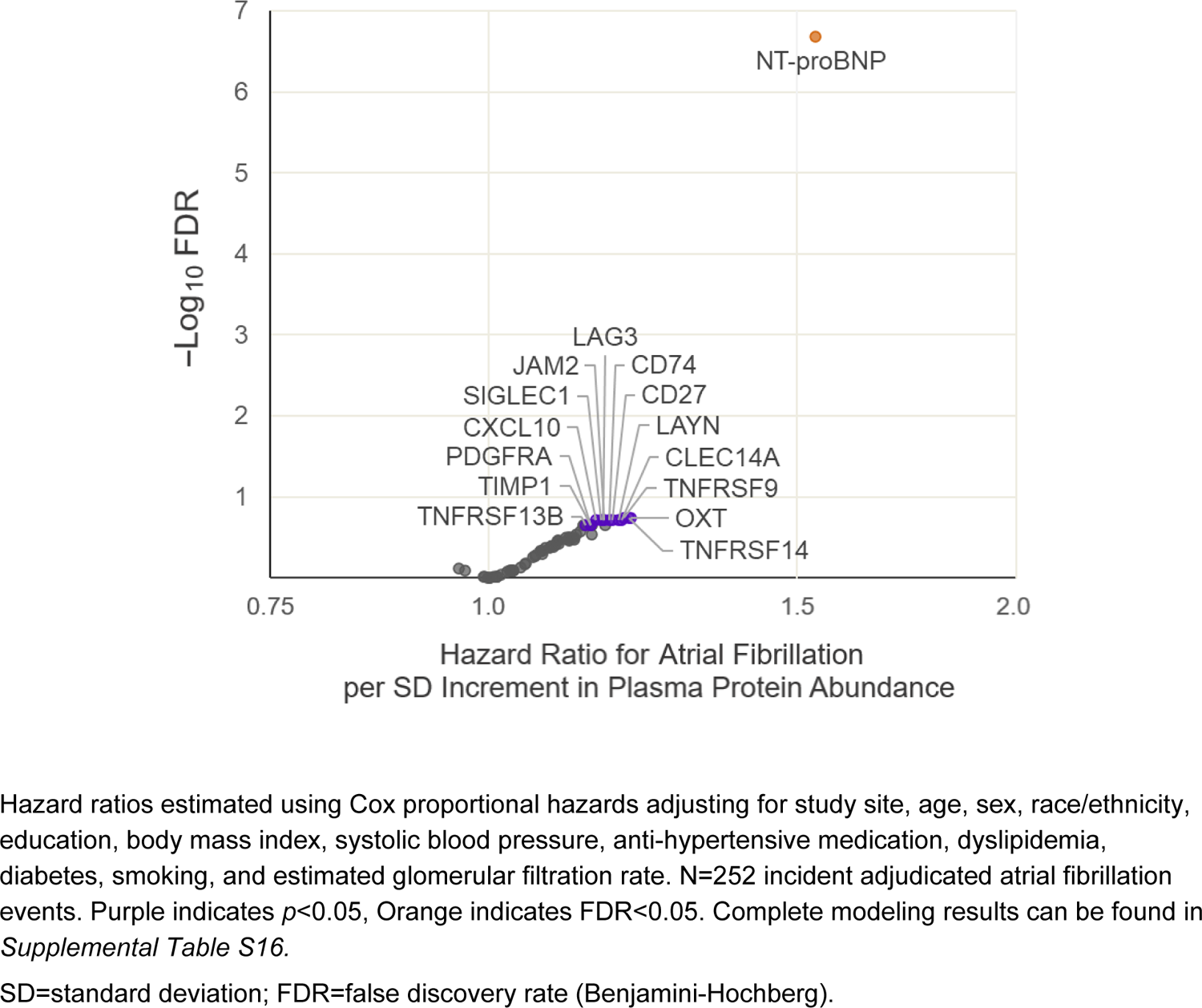
Volcano plot of association between identified HIV-associated proteomic signature of left atrial size and time to incident adjudicated atrial fibrillation in the Multi-Ethnic Study of Atherosclerosis (*n*=2185)

**SUPPLEMENTAL FIGURE S7.**
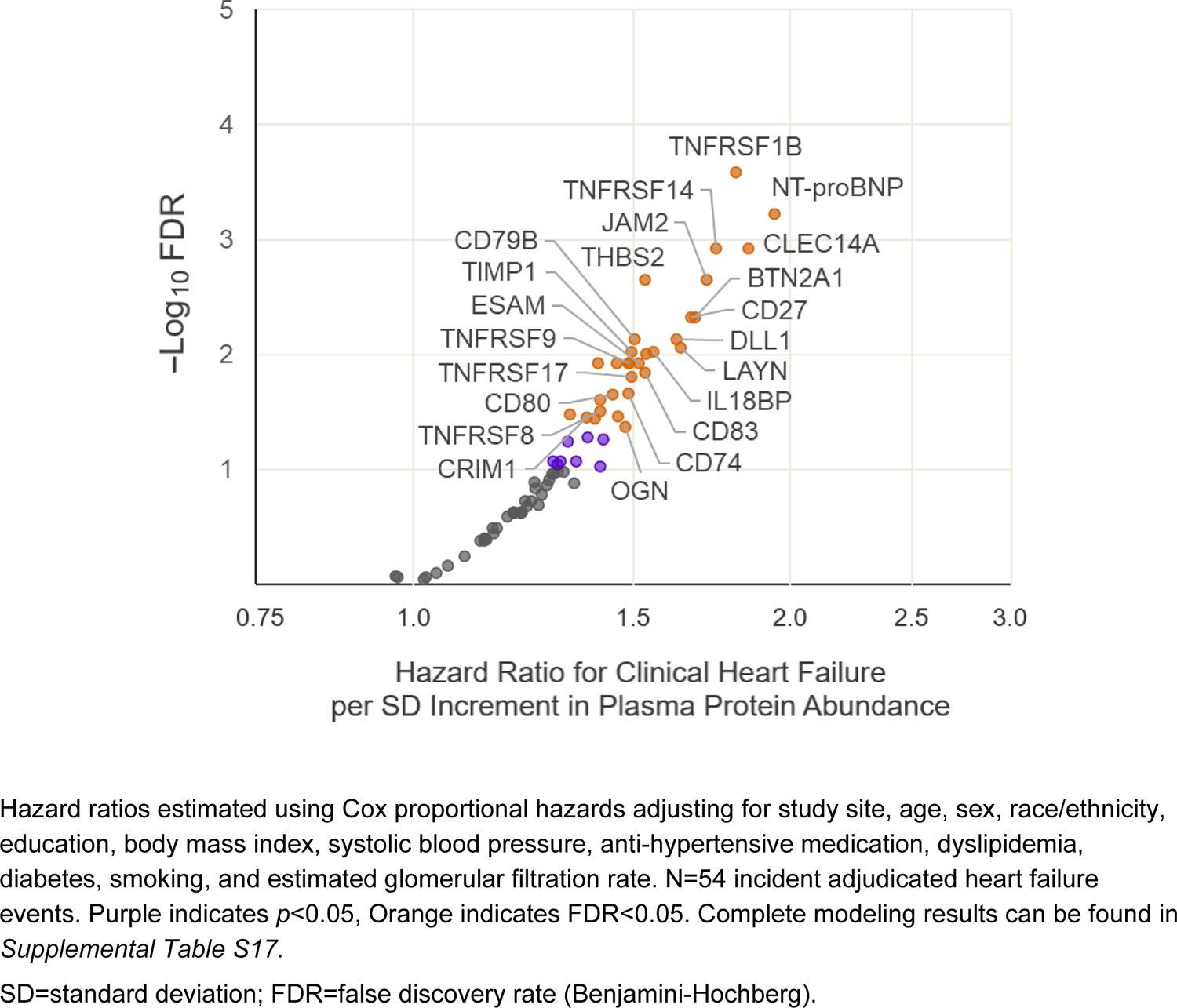
Volcano plot of associations between identified HIV-associated proteomic signature of left atrial size and time to incident adjudicated clinical heart failure in the Multi-Ethnic Study of Atherosclerosis (*n*=2273)

